# Longitudinal evolution of the transdiagnostic prodrome to severe mental disorders: a dynamic temporal network analysis informed by natural language processing and electronic health records

**DOI:** 10.1101/2024.03.08.24303965

**Authors:** Maite Arribas, Joseph M. Barnby, Rashmi Patel, Robert A. McCutcheon, Daisy Kornblum, Hitesh Shetty, Kamil Krakowski, Daniel Stahl, Nikolaos Koutsouleris, Philip McGuire, Paolo Fusar-Poli, Dominic Oliver

## Abstract

**Importance:** Modelling the prodrome to severe mental disorders (SMD), including unipolar mood disorders (UMD), bipolar mood disorders (BMD) and psychotic disorders (PSY), should consider both the evolution and interactions of symptoms and substance use (prodromal features) over time. Temporal network analysis can detect causal dependence between and within prodromal features by representing prodromal features as nodes, with their connections (edges) indicating the likelihood of one feature preceding the other. In SMD, node centrality could reveal insights into important prodromal features and potential intervention targets. Community analysis can identify commonly occurring feature groups to define SMD at-risk states.

**Objective:** To develop a global transdiagnostic SMD network of the temporal relationships between prodromal features, and to examine within-group differences with sub-networks specific to UMD, BMD and PSY

**Design:** Retrospective (2-year), real-world, electronic health records (EHR) cohort study. Validated natural language processing algorithms extracted the occurrence of 61 prodromal features every three months from two years to six months prior to SMD onset. To construct temporal networks of prodromal features, we employed generalized vector autoregression panel analysis, adjusting for covariates.

**Setting:** South London and Maudsley NHS Foundation Trust EHRs.

**Participants:** 6,462 individuals with an SMD diagnosis (UMD:2,066; BMD:740; PSY:3,656).

**Main Outcomes:** Edge weights (partial directed correlation coefficients, *z*) in autocorrelative, unidirectional and bidirectional relationships. Centrality was calculated as the sum of (non-autoregressive) connections leaving (out-centrality, *c_out_*) or entering (in-centrality, *c_in_*) a node. The three sub-networks (UMD, BMD, PSY) were compared using permutation analysis. Community analysis was performed using Spinglass.

**Results:** The SMD network was characterised by strong autocorrelations (0.04 *≤ z ≤* 0.10), predominantly positive connections, and aggression (*c_out_*=.103) and tearfulness (*c_in_*=.134) as the most central features. The UMD sub-network showed few significant differences compared to PSY (3.5%) and BMD (0.8%), and BMD-PSY showed even fewer (0.4%). One positive psychotic (delusional thinking-hallucinations-paranoia) and two behavioural communities (aggression-cannabis use-cocaine use-hostility, aggression-agitation-hostility) were the most common.

**Conclusions and Relevance:** This study represents the most extensive temporal network analysis conducted on the longitudinal interplay of SMD prodromal features. These findings provide further evidence to support transdiagnostic early detection services across SMD, refine assessments to detect individuals at risk and identify central features as potential intervention targets.

## 1. BACKGROUND

Severe mental disorders (SMD) include non-psychotic unipolar mood disorders (UMD), non-psychotic bipolar mood disorders (BMD) and psychotic disorders (PSY), and are characterised by high clinical, societal, familial and personal burden.^1–3^ Electronic health records (EHRs) can provide an opportunity to examine prodromal symptoms contemporaneously, reducing recall bias and enriching our insight into symptom presentation during the prodrome.^4^ This knowledge can help enhance specialised preventive care for people at-risk of emerging SMD.

Temporal network analysis, as an implementation of dynamic systems theory^5^, allows statistical modelling of the relationships between nodes (prodromal features) as edges within a dynamic network (e.g. prodrome) over time.^6^ Weak, sparse networks are more modifiable, while strong, dense networks resist change^7^, needing intensive interventions to alter them^8^ (e.g. preventing SMD onset). Given that edges in temporal networks satisfy the condition that cause precedes effect, they can suggest directed Granger causality between features^9^, potentially enhancing our understanding of SMD development.^10^ Node centrality, representing connection strength in and out of a node,^11^ may highlight the significance of a prodromal feature in the progression of the disorder and its potential as an intervention target due to its influence from/on other prodromal features.^12–15^ Communities are subgroups of nodes which are more densely connected amongst each other than with nodes outside of the subgroup^16^ and could help identify core prodromal connection pathways across SMD.^17^

Firstly, we aimed to develop a global transdiagnostic SMD network to quantify the temporal relationships between prodromal features. Secondly, we aimed to examine within-group differences by computing and comparing sub-networks specific to UMD, BMD and PSY.

## 2. METHODS

### 2.1. Data Source

Data were from the South London and Maudsley National Health Service Foundation Trust (SLaM). SLaM provides secondary mental healthcare across four socioeconomically diverse South London boroughs (eMethods 1). A Clinical Record Interactive Search (CRIS) tool was implemented in the EHR to facilitate research with full but anonymised clinical information.^18^ CRIS has already been extensively validated in previous research studies.^19–21^ CRIS received ethical approval as an anonymised dataset for secondary analyses from Oxfordshire REC C (Ref: 23/SC/0257).

### 2.2. Study Design

Retrospective (2-year), real-world, EHR cohort study (Figure 1). The 2 years were chosen to mirror the typical duration of care in clinical services for primary indicated prevention of SMD (72.4% provide care for 24 months or less).^22^ The index date reflected the date of the first diagnosis within an individual’s SMD group recorded in the EHR (index diagnosis, T-0mo, Figure 1). The antecedent date was defined by a data cut-off at 6 months before the index date (T-6mo), defining the antecedent period, to avoid overlap with the actual onset of SMD. The lookback period (Figure 1) was defined as the 1.5 years before the antecedent date (T-6mo). To minimise violations of the time invariance assumption imposed by network analyses,^23^ we regularised the 1.5-year lookback period into six three-month follow-up intervals.

**Figure 1.**
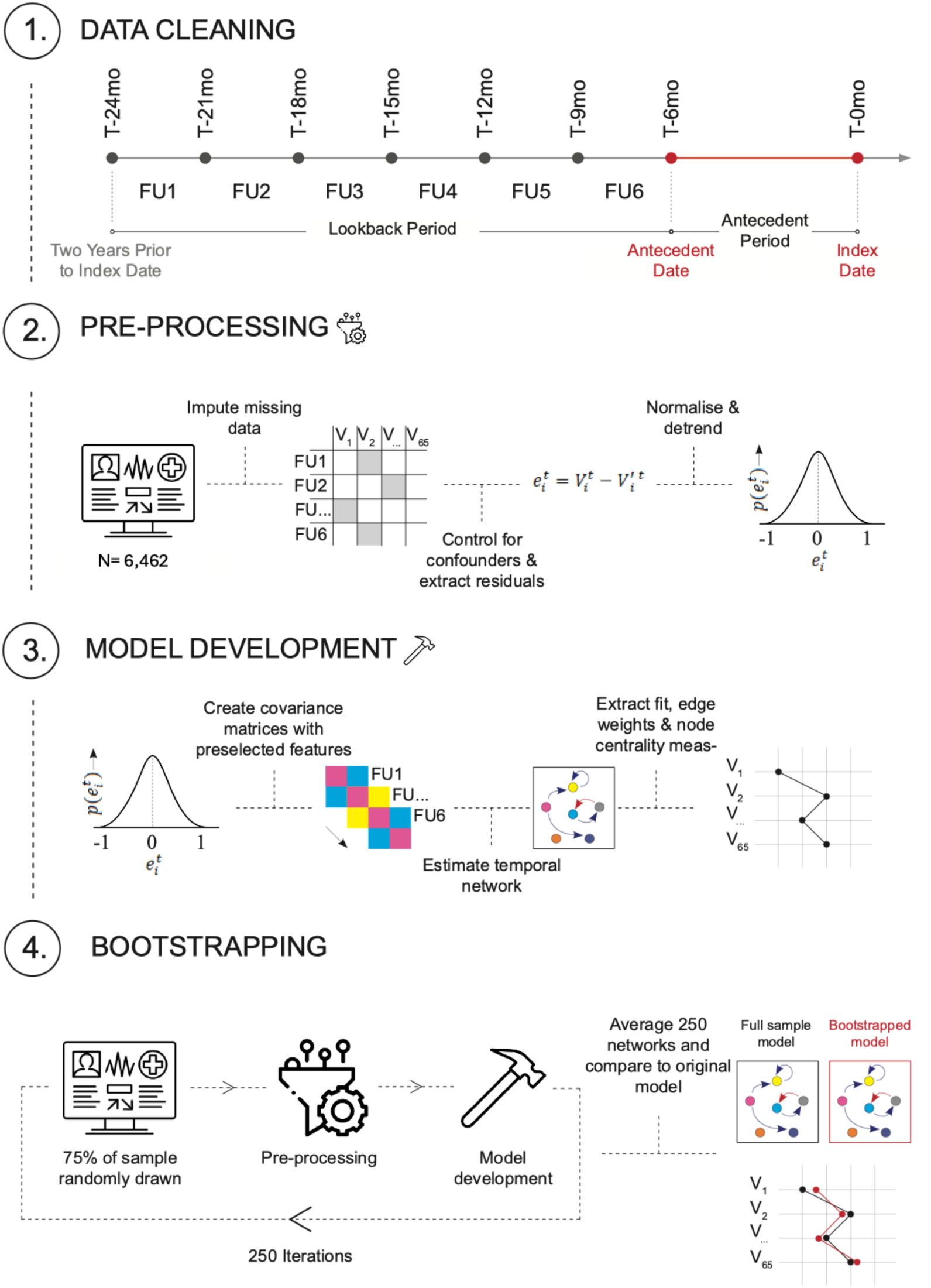
Study design. The look-back period was split into six three-month follow-up intervals (FU 1-6) relative to the index date (T-0mo) of SMD diagnosis. This pipeline (steps 1-4) was followed for both the primary analysis (SMD model) and secondary analysis (sub-networks).

### 2.3. Study Population

All individuals accessing SLaM services between 1^st^ January 2008 and 10^th^ August 2021 and receiving a primary (i.e. not comorbid) ICD-10 index diagnosis of any SMD were eligible. SMD was defined as either UMD, BMD or PSY (operationalised as in eTable 1). Individuals with multiple SMD diagnoses were assigned the diagnosis of greatest severity (i.e. UMD<BMD<PSY).

Individuals with EHR entries (e.g. clinical notes and letters recorded in each month) recorded exclusively after the index date or exclusively in the antecedent period were excluded as they had no detectable prodrome. Individuals who only had empty EHR entries within the lookback period were also excluded, as well as those with EHR entries recorded within four or fewer follow-up intervals within the lookback period, as they did not have sufficient data to contribute to the fitted networks.

### 2.4. Variables

At index date, data were extracted from structured text on age, gender, self-assigned ethnicity (UK Office of National Statistics, eTable 2), ICD-10 diagnoses and prescription of antipsychotics, antidepressants, mood stabilisers and anxiolytics (see eTable 3 for medication classification details).

During the lookback period, data were extracted as binary variables on the occurrence (yes:1/no:0) of 61 natural language processing (NLP)-based prodromal features across each follow-up interval (FU 1-6; Figure 1). These NLP algorithms convert unstructured EHR information (i.e. free text) into structured quantifiable data.^24^ NLP algorithms with precision ≥80% (mean=90%) were included (see eMethods 2 for further details on NLP algorithm development and validation, and eTable 4 for the final list of NLP algorithms employed). Within each follow-up interval, the EHR entry frequency (number of entries) and length (total number of words recorded across all entries) were computed.

### 2.5. Statistical analysis

All analyses were conducted in R (version 4.2.3) on a virtual machine (AMD EPYC 7763 64-Core Processor) in Ubuntu 22.04.1 operating system. All analysis code is publicly available on GitHub: https://github.com/m-arribas/network_analysis.git.

#### 2.5.1. Sociodemographic and Clinical Characteristics

We computed descriptive analyses for sociodemographic variables at index date (age, gender, self-assigned ethnicity) as well as the proportion (N [%]) of individuals with specific ICD-10 diagnoses and prescription of antipsychotics, antidepressants, mood stabilisers and anxiolytics at index in UMD, BMD and PSY. A sensitivity analysis was completed to test for any sampling bias in the final population compared to excluded individuals (eMethods 3).

#### 2.5.2. Network analysis

As a primary analysis, we quantified a set of local network metrics in a transdiagnostic SMD network (hereby called “SMD network”) on the entire study population. In a secondary analysis, we repeated this on each SMD sub-sample separately (UMD, BMD, PSY), to compute three diagnosis-specific sub-networks (hereby called “sub-networks”). For each network (SMD network and three sub-networks), the following steps (pre-processing, network development and stability assessment) were repeated separately in each relevant dataset using a similar step-wise procedure to prior work modelling temporal features in psychopathology.^25^

Pre-processing methods are detailed in eMethods 4. Importantly, during pre-processing, we controlled for a set of demographic, medication and clinical variables through linear regression and used the resultant residuals (on a numeric scale) in our networks to isolate prodromal feature variability over time.

To build such networks (network development), we used Lag-1 Panel Graphical Vector Auto-Regressive (GVAR) analyses,^26^ estimated with the ‘psychonetrics’^27^ package (version 0.10) using the ‘panelgvar’ function. In these network models, nodes represent variables (prodromal feature occurrences) and edges representheir relationship when conditioned on all other nodes in the network in a number of fixed measurement occasions. In the case of contemporaneous and between-individuals networks, these are undirected relationships. In the case of temporal networks, edges are directed (e.g., node A → B) and conditioned upon the current (t) and previous (t-1) state of all nodes (including itself).^26^ This uses a model drawing upon fixed effect lag-k variance-covariance matrices estimated from the data. Edges within temporal networks satisfy the condition that cause precedes effect, which is indicative of Granger causality.^9^ In all networks, we utilised the maximum-likelihood estimator (MLE) in the ‘psychonetrics’ package.

For each network (SMD network and three sub-networks), we extracted the temporal (within-individual lagged outcomes averaged across follow-up intervals), contemporaneous (within-individual relationships between nodes within the same three-month follow-up interval), and between-individuals subject matrices (relationships between nodes averaged across individuals within the same three-month follow-up interval). From each matrix, the edge weights (*z*) were estimated as partial directed correlation coefficients (for temporal networks) and partial correlation coefficients (for contemporaneous and between-individual networks). Temporal edges were categorised as autocorrelative, unidirectional, or bidirectional. Degree centrality measures were extracted from each graph. For temporal networks, centrality was defined as the sum of absolute (directed) edge weights in (in-centrality*, c_in_*) and out (out-centrality*, c_out_*) of a node. Autoregressive edges were excluded from the centrality calculation to isolate the effect of one node on other nodes or vice versa. Detailed definitions of centrality in contemporaneous and between-individual networks are presented in eMethods 5.

To evaluate the robustness of the edge weight estimates and to avoid overfitting in our networks, we computed the stability of edges within each network using bootstrapping procedures: over 250 iterations, 25% of the sample was randomly held out and the full model refitted on the remaining 75% of participants (following standard methods).^28^ Within each iteration, the selected data was pre-processed in the same manner as in the full model to control for errors and variance within the data cleaning and scaling process. The averaged edge weights and 95%CIs over all 250 iterations were retained and reported. All edges with 95%CIs crossing zero were forced to 0.

Model fit was assessed using the following fit statistics: relative fit indices (Tucker Lewis Index [TLI]) and the non-centrality-based indices (Comparative Fit Indices [CLI] and Root Mean Square Error Approximation [RMSEA] with 95% confidence intervals [95%CIs]). Absolute fit indices are reported (Chi-square). However, due to the metric’s sensitivity to sample size, Chi-square estimates were not interpreted. For TLI and CFI, values 0.90–0.95 are considered accepted cut-offs for good fit^29^. RMSEA<0.05 indicate excellent fit; RMSEA<0.10 indicate good fit. For each of the networks, we generated simulated data using the approximated full model structure and refitted the model on the simulated data to estimate model recovery using the same model fit metrics. The residual variance was estimated using Cholesky decomposition.

Each network was visualised with a graph where node placement was determined by Fruchterman-Reingold algorithm^30^ and edge weights were thresholded to display the top 40 (temporal) or 20 (contemporaneous/between-subject). For equal edge weights, non-superiority was assumed and all edge weights were displayed. For visualisation purposes, nodes in the graphs are categorized into six broader clusters: depressive, manic, negative, positive, substance use and other. This categorization, developed by Jackson et al ^24^, is based on previous studies that utilised symptomatology factor analysis ^31,32^ and are aligned with publicly available, validated NLP dictionaries ^33^.

#### 2.5.3. Permutation analysis

To test for statistically significant differences in the temporal, contemporaneous, and between-subject relationships across the three sub-networks (UMD, BMD, PSY) we conducted permutation analyses^34^.

To generate networks with the same topology required for valid comparisons, we re-fitted the three original sub-networks restricted to common features only, after pre-processing the data for each sub-sample.

In each permutation iteration, raw data in each sub-sample population (UMD, BMD, PSY) was randomly re-sampled for each node. The permuted dataset for each sub-sample was then pre-processed in the same manner as in the original sub-networks, and permuted sub-networks were fitted. From each permuted sub-network, temporal, contemporaneous and between-subjects matrix estimates were obtained, as in the main analysis. For each edge, the difference in permuted edge weights was calculated (UMD-BMD, UMD-PSY and BMD-PSY). Each edge weight comparison was visually inspected using histograms to assess the normality of the permuted data.

For each edge weight comparison, the number of iterations where the permuted difference was equal to or greater than the absolute observed difference (from the actual dataset) was divided by the total number of permutations (250) to obtain the p-value. We corrected the resulting p-values for multiple comparisons using the false discovery rate method set at 5% to ensure the robustness of our findings and used p*_permuted_*<0.05 as the threshold for statistical significance. Observed differences in edge weights (from the actual dataset) and corrected p-values (from the permutation analysis) were reported and visualised with heatmaps.

#### 2.5.4. Community analysis

We conducted community analyses using the Spinglass algorithm^35^ within the SMD network and three sub-networks (UMD, BMD, PSY) from the permutation analysis. For each network, a heatmap was produced to display the probability of node-node co-occurrence (covariance) within community structures, and the three most occurring communities were visualised.

To detect community structures, subgraphs of nodes which are more densely connected amongst each other than with nodes outside of the subgraph^16^, we conducted community analyses within the SMD network and the three sub-networks (UMD, BMD, PSY) from the permutation analysis.

For this, the Spinglass algorithm^35^ was run using the edge weight matrices of each network. This algorithm was selected due to its ability to handle weighted edges with directionality.^36^ which other algorithms are unable to. The Spinglass algorithm uses simulations based on the Potts-model from statistical mechanics.^37^ Due to the simulations, the algorithm is non-deterministic. Following previous methods^38^, we ran the algorithm with 1000 iterations to calculate the average probability of 2 nodes co-occurring within the same community. The spin argument was restricted to 12 (SMD network) and 8 (UMD, BMD, PSY), to detect community structures composed of more than 2 nodes.

For each network, a heatmap was produced to display the probability of node-node co-occurrence, with nodes ordered by hierarchical clustering^38^. The three most occurring communities were visualised.

## 3. RESULTS

### 3.1. Study Population

The final study population (n=6,462 Table 1; eTable 5-6) had EHR entries with more than four follow-up intervals (mean number of intervals [SD]=5.67 [0.46]) within the lookback period (Figure 2). Included participants were similar to excluded participants in terms of sociodemographics and clinical characteristics (eResults 1). Seven participants were excluded as the imputation method was unable to converge on stable approximations.

**Figure 2.**
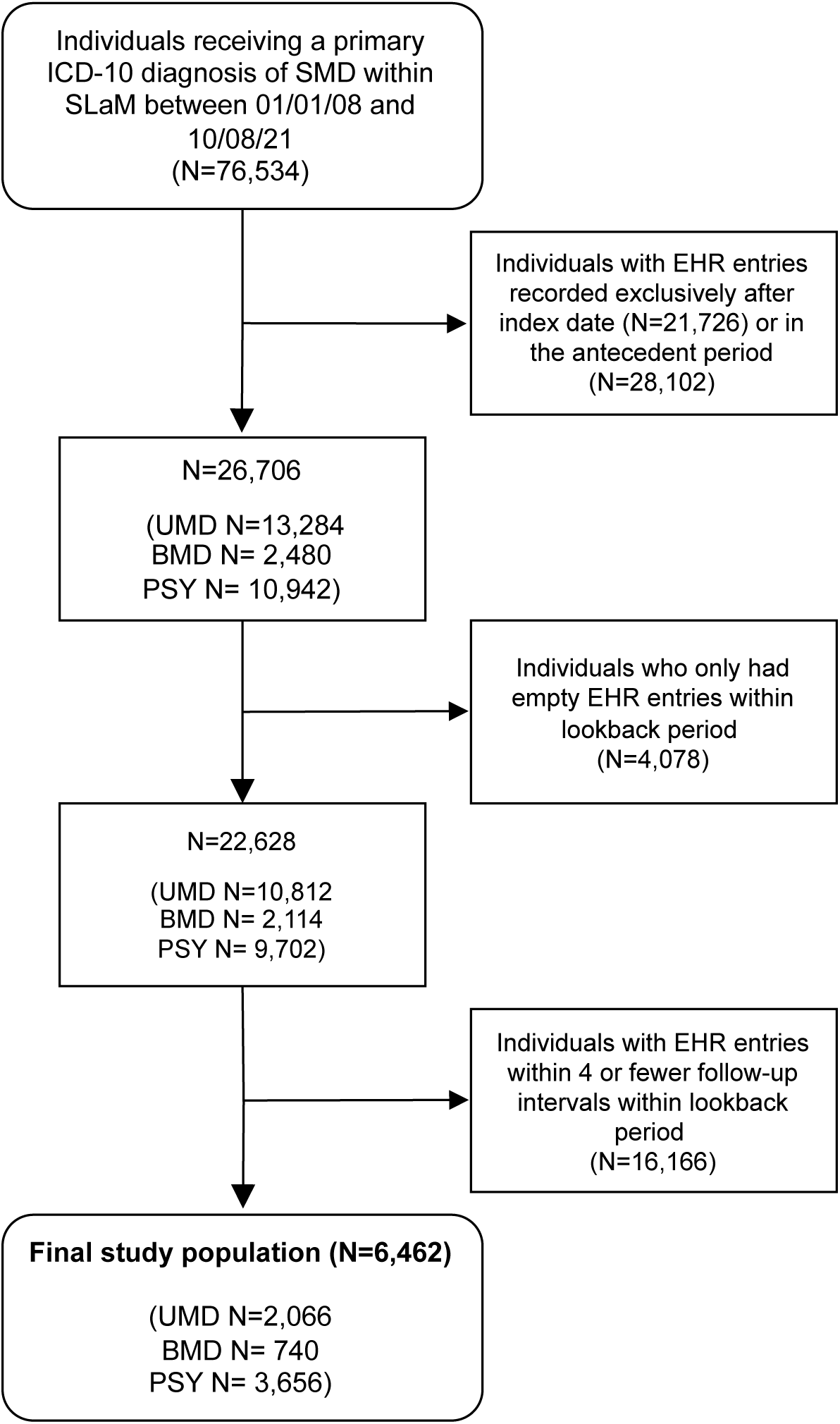
Flow-chart of the study population.

**Table 1.** Demographics and medication variables at index of final study population (N=6,462). Continuous variables are represented by mean (SD), categorical variables are represented by count (frequency).

| **Characteristic** | **Whole sample, N=6,462** | **UMD, N = 2,066** | **BMD, N = 740** | **PSY, N = 3,656** |
| --- | --- | --- | --- | --- |
| **Age** | 43.5 (16.1) | 43.9 (18.5) | 44.0 (15.2) | 43.3 (14.8) |
| **Gender** |  |  |  |  |
| Female | 3,321 (51) | 1,315 (64) | 471 (64) | 1,535 (42) |
| Male | 3,138 (49) | 748 (36) | 269 (36) | 2,121 (58) |
| Other | 3,321 (51) | 3 (0.1) | 0 (0) | 0 (0) |
| missing | 0 (0) | 0 (0) | 0 (0) | 0 (0) |
| **Ethnicity** |  |  |  |  |
| White | 3,536 (55) | 1,371 (66) | 512 (69) | 1,653 (45) |
| Black | 1,906 (29) | 324 (16) | 117 (16) | 1,465 (40) |
| Other | 223 (3.5) | 82 (4.0) | 23 (3.1) | 118 (3.2) |
| Asian | 459 (7.1) | 137 (6.6) | 50 (6.8) | 272 (7.4) |
| Mixed | 209 (3.2) | 78 (3.8) | 24 (3.2) | 107 (2.9) |
| missing | 129 (2.0) | 74 (3.6) | 14 (1.9) | 41 (1.1) |
| **Antidepressants** | 2,533 (39) | 1,019 (49) | 292 (39) | 1,222 (33) |
| **Mood stabilisers** | 1,074 (17) | 141 (6.8) | 384 (52) | 549 (15) |
| **Anxiolytics** | 1,566 (24) | 378 (18) | 239 (32) | 949 (26) |
| **Antipsychotics** | 3,426 (53) | 350 (17) | 399 (54) | 2,677 (73) |

### 3.2. Primary Analysis (SMD network)

Out of the 61 NLP-derived prodromal features, 38 displayed near-zero variance and were excluded, leaving 23 features for the analyses (eFigure 1, eTable 7). A saturated model (a densely connected network with all available edges) was fitted with the 23 features at 6 follow-up intervals (Figure 3A). This network demonstrated excellent fit (RMSEA=.010 [95%CI: .0099, .011]; *X*^2^(8625)=14,455, p<.0001; CFI=.96; TLI=.96) and had better fit than a sparse network (pruned edges) (Δ*X*^2^(737)=2,184, p<.0001). The model showed high recoverability (eResults 2) and robustness (see Figure 3B and eTables 8-9 for actual model and bootstrapped estimates, and eFigure 2 for the un-thresholded SMD network).

**Figure 3.**
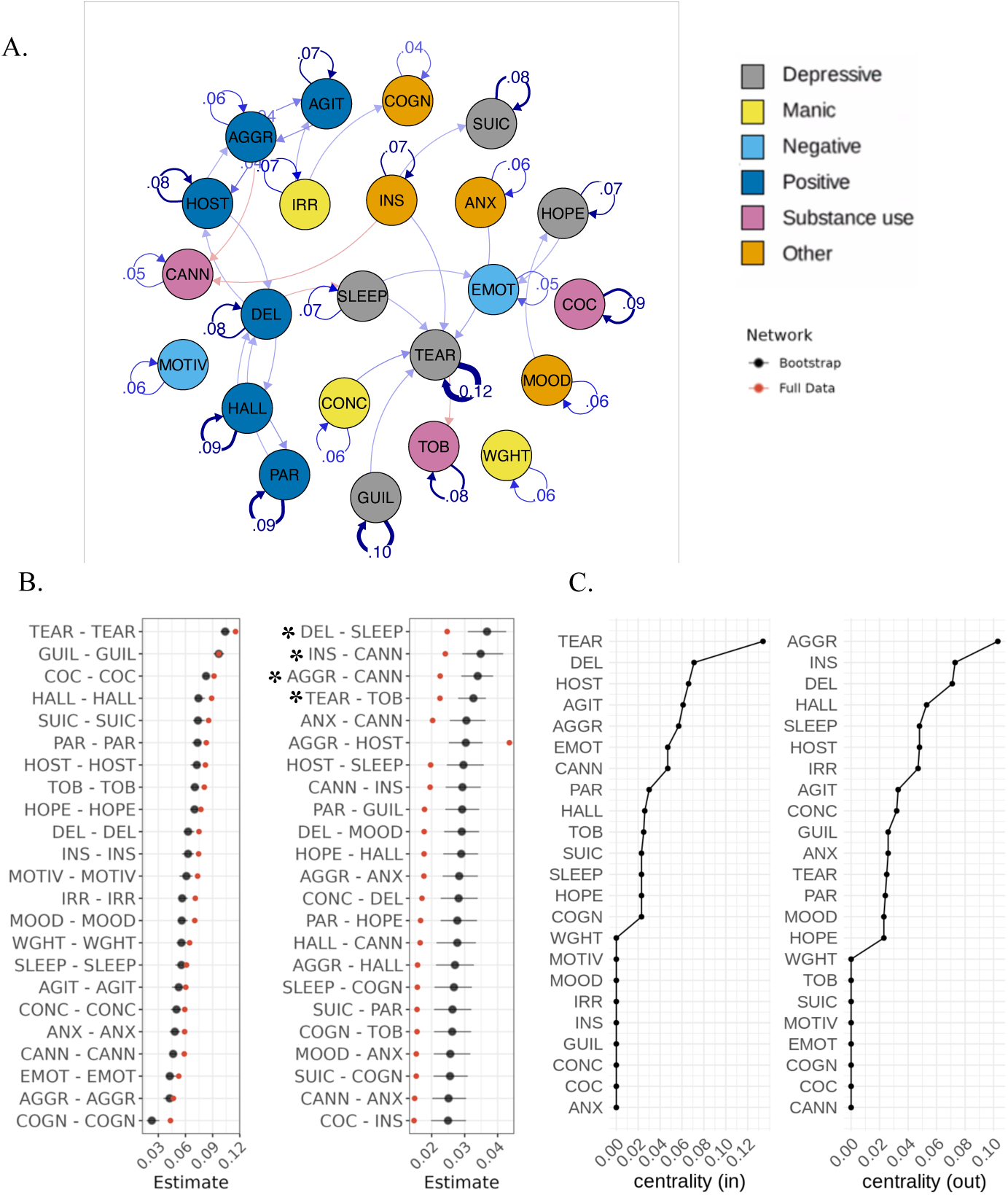
Temporal relationships between nodes in SMD network **A.** Temporal network graph displaying positive (blue) and negative (red) relationships between nodes from actual model estimates. Edges are displayed as lines, with the thickness representing the strength of the edge weight estimate (partial directed correlation coefficient, *z*). Edges are thresholded (|*z*| > .022) and labelled (|*z*| > .03). For visualisation purposes, nodes are clustered into six categories (depressive, manic, negative, positive, substance use and other) according to the type of prodromal feature. **B.** Bootstrapped (250 repetitions; black) vs actual model (n=6,462; red) edge weight estimates (|*z*| > .022). Edges are directed such that “node1 – node2” represent the edge from node1 to edge2. All edges were positive except the one marked with an asterisk (DEL-SLEEP, INS-CANN, AGGR-CANN, TEAR-TOB). **C.** Centrality measures for all nodes AGGR: aggression, AGIT: agitation, ANX: anxiety, CANN: cannabis use, COC: cocaine use, COGN: cognitive impairment, CONC: poor concentration, DEL: delusional thinking, EMOT: emotional withdrawal, GUIL: guilt, HALL: hallucinations (all), HOPE: feeling hopeless, HOST: hostility, INS: poor insight, IRR: irritability, MOOD: mood instability, MOTIV: poor motivation, PAR: paranoia, SLEEP: disturbed sleep, SUIC: suicidality, TEAR: tearfulness, TOB: tobacco use, WGHT: weight loss

The strongest autocorrelation was observed for tearfulness (partial directed correlation coefficient, *z*=.12), with all the other autocorrelations between 0.04-0.10 (Figure 3A). The most prominent unidirectional relationships were negative: delusional thinking-disturbed sleep (*z*_12_=-.02), poor insight-cannabis use (*z*_12_=-.02), aggression-cannabis use (*z*_12_=-.02) and tearfulness-tobacco use (*z*_12_=-.02), with one positive unidirectional relationship between anxiety-cannabis use (*z*_12_=.02). All other unidirectional relationships were | *z*_12_| <.02.

With respect to bidirectional relationships, positively recurring pairs were observed between aggression-hostility (*z*_12_=.04, *z*_21_=.02), delusional thinking-hallucinations (*z*_12_=.03, *z*_21_=.03), aggression-agitation (*z*_12_=.04, *z*_21_=.03) and delusional thinking-hostility (*z*_12_=.02, *z*_21_=.02).

Considering centrality (Figure 3C), aggression (*c_out_*=.103), poor insight (*c_out_*=.073) and delusional thinking (*c_out_*=.071) had the strongest out-centrality, whereas tearfulness (*c_in_*=.134), delusional thinking (*c_in_*=.071) and hostility (*c_in_*=.066) had the strongest in-centrality (eTable 10).

Results and visualisations for the contemporaneous and between-subject relationships of nodes are presented in eResults 3 and eFigure 3. See eTable 9 for actual model and bootstrapped estimates.

### 3.3 Secondary Analysis (sub-networks)

Out of the 61 NLP-derived prodromal features, after applying the relevant exclusions within each sub-sample, 21 features were included for the UMD network, 19 for BMD and 24 for PSY (eMethods 6). A saturated model was fitted with the relevant features at 6 follow-up intervals in each sub-sample (UMD, BMD, PSY) (Figure 4). Similarly to the primary analysis, saturated networks showed excellent fit and better fit than sparse models for the three networks (UMD: Δ*X*^2^(706)=1585, p<.0001; BMD: Δ*X*^2^(602)=1557, p<.0001; PSY: Δ*X*^2^(862)=2717.5, p<.0001). Further model fit results, including recoverability (eResults 4), and bootstrapping estimates (eTable 11, eFigure 4) can be found in the Supplement.

**Figure 4.**
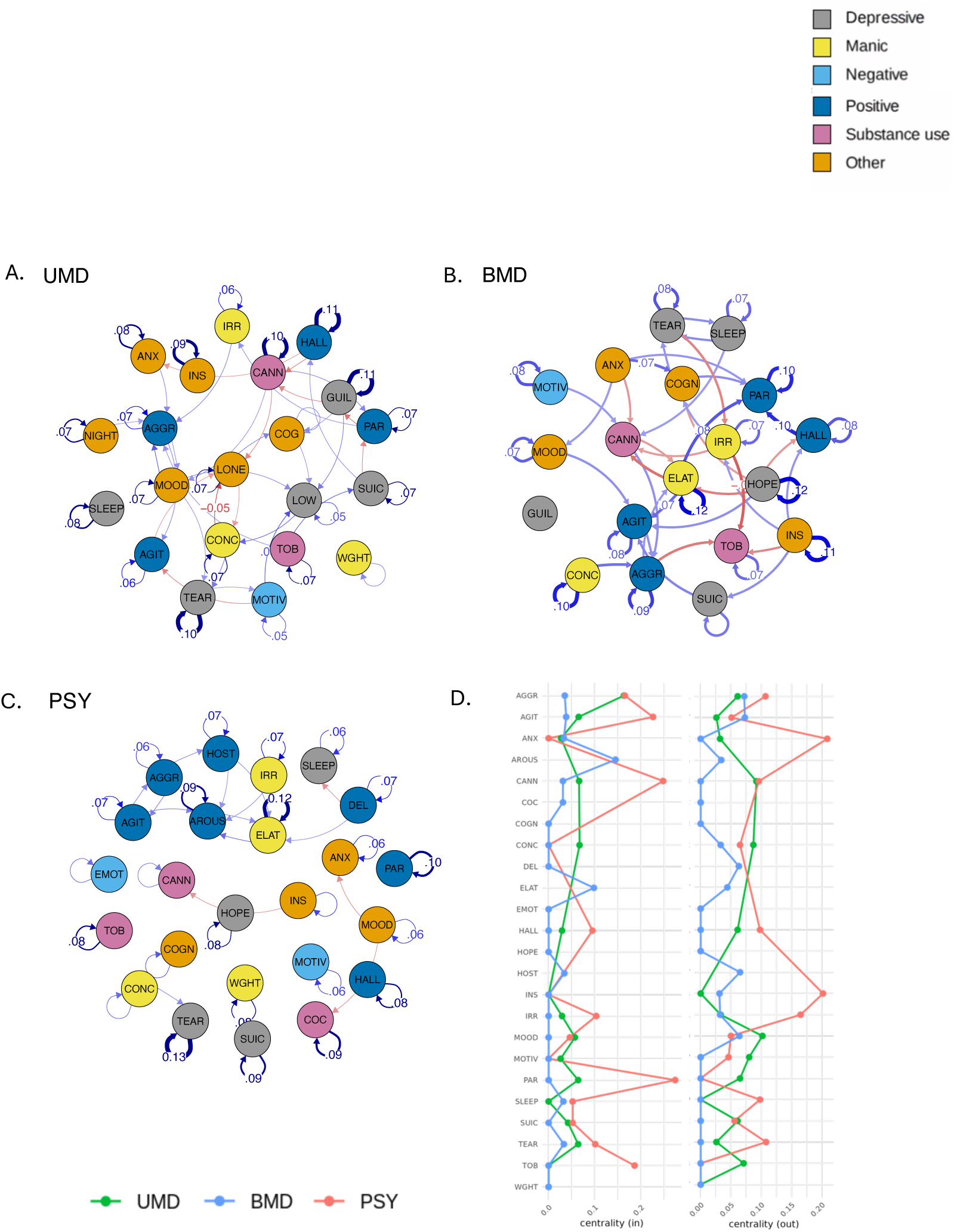
Temporal relationships between nodes in sub-networks Temporal network graphs displaying positive (blue) and negative (red) relationships between nodes from actual model estimates for sub-networks (**A.** UMD, **B.** BMD, **C.** PSY). Edges are displayed as lines, with the thickness representing the strength of the edge weight estimate (partial directed correlation coefficient, *z*). Edges are thresholded (UMD: |*z*| > .026, BMD: |*z*| > .045, PSY: |*z*| > .03) and labelled (UMD: |*z*| > .04, UMD: |*z*| > .06, UMD: |*z*| > .05). For visualisation purposes, nodes are clustered into six categories (depressive, manic, negative, positive, substance use and other) according to the type of prodromal feature. D. Centrality measures for all nodes in sub-networks (green: UMD, blue: BMD, red: PSY) AGGR: aggression, AGIT: agitation, ANX: anxiety, AROUS: arousal, CANN: cannabis use, COC: cocaine use, COGN: cognitive impairment, CONC: poor concentration, DEL: delusional thinking, ELAT: elation, EMOT: emotional withdrawal, GUIL: guilt, HALL: hallucinations (all), HOPE: feeling hopeless, HOST: hostility, INS: poor insight, IRR: irritability, LONE: feeling lonely, LOW: low energy, MOOD: mood instability, MOTIV: poor motivation, NIGHT: nightmares, PAR: paranoia, SLEEP: disturbed sleep, SUIC: suicidality, TEAR: tearfulness, TOB: tobacco use, WGHT: weight loss

Considering centrality, in the UMD sub-network, aggression (*c_in_*=.163), low energy (*c_in_*=.141), and feeling lonely (*c_in_*=.109) had the strongest in-centrality, whereas mood instability (*c_out_*=.102), guilt (*c_out_*=.094) and cannabis use (*c_out_*=.092), had the strongest out-centrality (eTable 13A). In the BMD sub-network, paranoia (*c_in_*=.274), cannabis use (*c_in_*=.248) and agitation (*c_in_*=.226) had the strongest in-centrality, whereas elation (*c_out_*=.209), anxiety (*c_out_*=.209), feeling hopeless (*c_out_*=.202) and poor insight (*c_out_*=.202) had the strongest out-centrality (eTable 13B). In the PSY sub-network, arousal (*c_in_*=.145), elation (*c_in_*=.098) and agitation (*c_in_*=.038) had the strongest in-centrality, whereas agitation (*c_out_*=.073), aggression (*c_out_*=.072) and hallucinations (*c_out_*=.072) had the strongest out-centrality (eTable 13C).

Edge weight results for each sub-network are reported in eResults 5. Moreover, results and visualisations for the contemporaneous and between-subject relationships of nodes for all sub-networks are in eResults 6. See eTable 12 for the actual model and bootstrapped estimates and eFigure 5 for un-thresholded sub-networks graphs.

**Figure 5.**
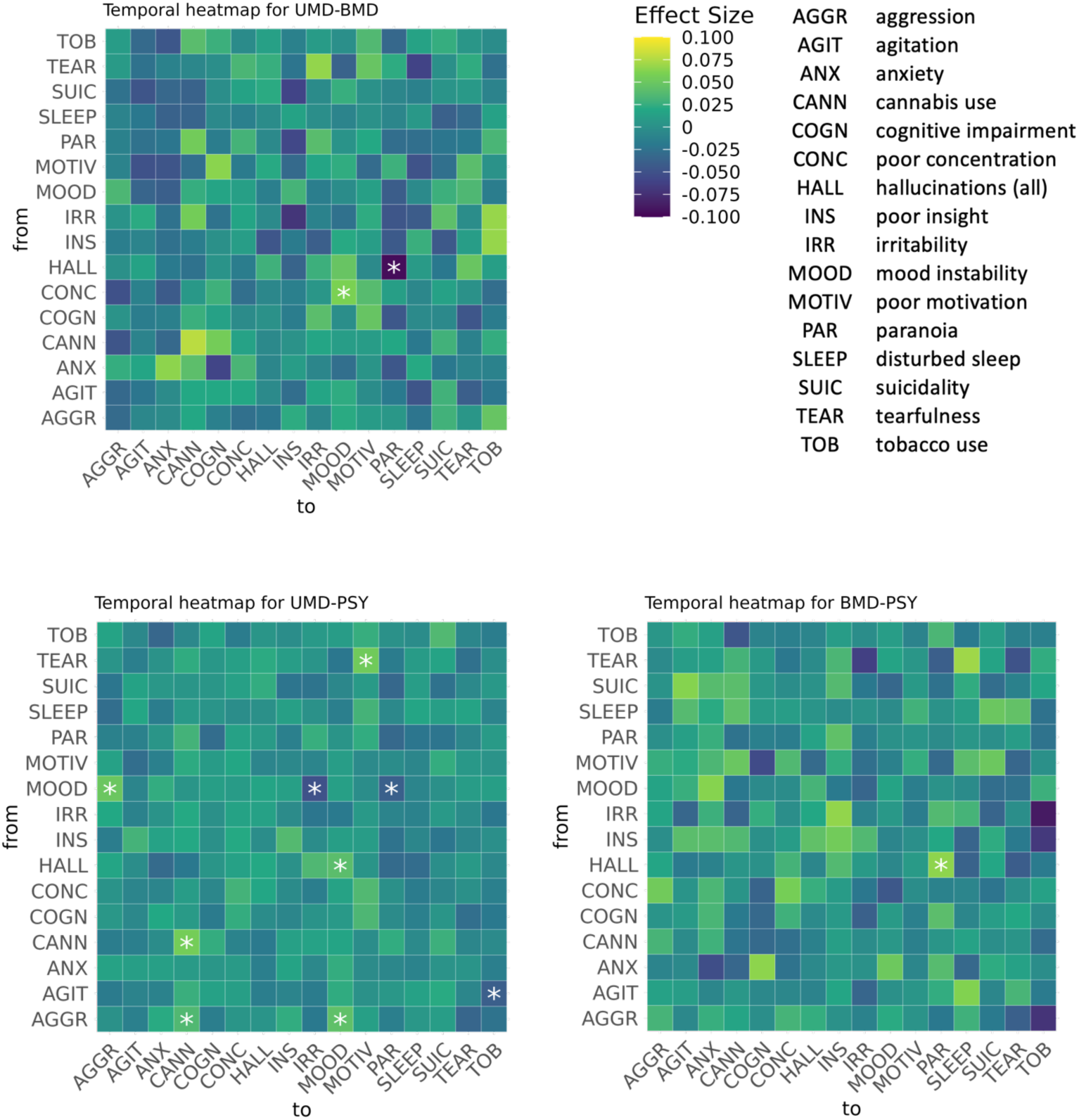
Heat-maps for pairwise edge comparisons (UMD-BMD, BMD-PSY, UMD-PSY) in temporal sub-networks in permutation analysis. Magnitude and direction of effect size is colour-coded such that for the pairwise comparison Group1-Group2, yellow indicates the edge estimate is more positive in Group1>Group2 and blue indicates the opposite Group1<Group2. Significant pairwise comparisons (corrected p<0·05) are marked with an asterisk (*).

### 3.4 Permutation Analysis

The final nodes for permutation analysis are found in eMethods 7, with the actual model estimates in eTable 14. The histograms of permuted edge weights exhibited a normal (bell-shaped), zero-centred curve, indicating that edge weights that lay outside of the null distribution following permutation testing are likely to be a true positive difference between networks. Out of all possible edge weight comparisons, few of them were significantly different: UMD-PSY (3.5%), followed by UMD-BMD (0.8%) and then BMD-PSY (0.4%).

UMD showed a significantly stronger edge weight compared to BMD from poor concentration to mood instability (*z*_UMD-BMD_=.06, p*_permuted_*<.001). In addition, the following edge weights were significantly stronger compared to PSY: aggression-cannabis use (*z*_UMD-PSY_=.036, p*_permuted_*<.001), aggression-mood instability(*z*_UMD-PSY_=.043, p*_permuted_*<.001), cannabis use (autocorrelation) (*z*_UMD-PSY_ =.056, p*_permuted_*<.001), hallucinations-mood instability (*z*_UMD-PSY_=.041, p*_permuted_*<.001), mood instability-aggression (*z*_UMD-PSY_=.050, p*_permuted_*<.001), tearfulness-poor motivation (*z*_UMD-PSY_=.053, p*_permuted_*<.001) (Figure 5).

BMD showed one edge weight that was significantly stronger than UMD from hallucinations to paranoia (*z*_BMD-UMD_ =.095, p*_permuted_*<.001), and another edge weight that was stronger compared to PSY from hallucinations to paranoia (*z*_BMD-PSY_ =.065, p*_permuted_*<.001) (Figure 5).

PSY showed significantly stronger edge weights compared to UMD for the following edges: agitation-tobacco use (*z*_PSY-UMD_=.039, p*_permuted_*<.001), mood instability to irritability (*z*_PSY-UMD_=.049, p*_permuted_*<.001) and mood instability to paranoia (*z*_PSY-UMD_=.039, p*_permuted_*<.001). PSY didn’t show any edge weights to be significantly stronger compared to BMD (Figure 5).

Histograms showing null distributions for significant temporal comparisons are in eFigure 6, with the results from the contemporaneous and between-subject matrices in eFigures 7-8.

### 3.5 Community analysis

Heatmaps of node-node covariance across communities are displayed in Figure 6. The most commonly occurring communities for the SMD network were: delusional thinking-hallucinations-paranoia (occurs in 4.5% of 1000 iterations of the Spinglass algorithm), aggression-cannabis use-cocaine use-hostility (2.8%) and aggression-agitation-hostility (1.9%). For UMD: irritability-hallucinations-paranoia (20.3%), agitation-disturbed sleep-mood instability-poor concentration-poor insight-tearfulness (14.5%) and cannabis use-cognitive impairment-irritability-poor motivation-suicidality-tobacco use (11.7%), aggression-anxiety-mood instability, and cannabis use-poor insight-suicidality-tobacco use (18.6%). For BMD: aggression-hallucinations-irritability-poor insight-poor motivation (44.9%), cognitive impairment-disturbed sleep-poor concentration-paranoia-tearfulness (23.6%) and, agitation-anxiety-cannabis use-mood instability-suicidality-tobacco use (11.3%). For PSY: poor insight-tearfulness-suicidality (19.9%), cannabis use-irritability-mood instability-poor motivation (13.9%), and anxiety-disturbed sleep-hallucinations (12.0%).

**Figure 6.**
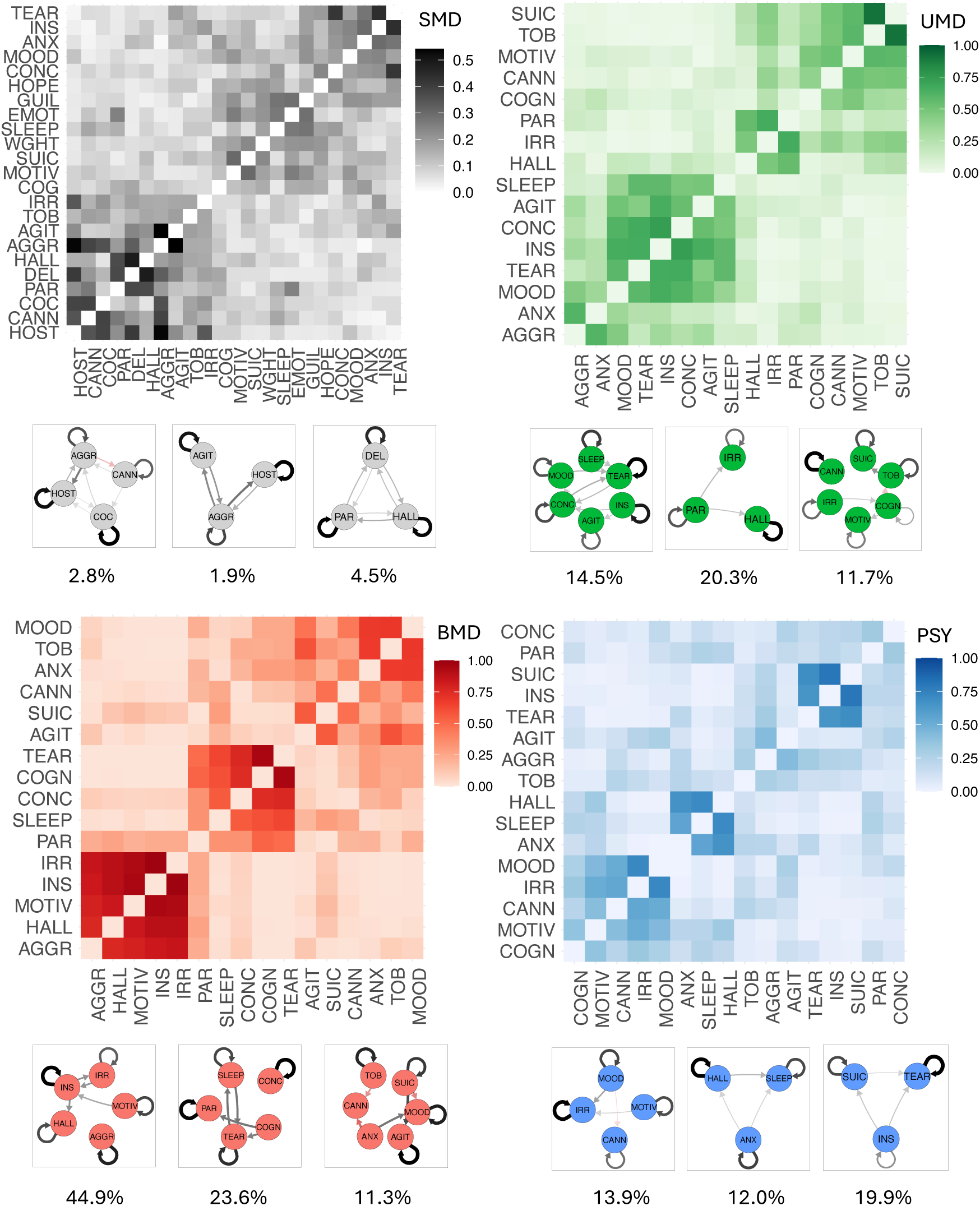
Heatmaps of node-node covariance across communities. The colour scale indicates the probability of 2 nodes co-occurring within the same community across 1000 iterations of Spinglass algorithm (spin restricted to 12 [SMD] and 8 [UMD, BMD, PSY]). The nodes are ordered by hierarchical clustering. The 3 most-occurring communities are displayed under each heatmap, with their % occurrence across 1000 iterations. Edges have been thresholded at |*z*| > 0.01 (SMD, PSY), > 0.02 (UMD) and > 0.035 (BMD), for visualisation purposes. Positive edges are displayed in black, and negative edges in red.

## 4. DISCUSSION

This study represents the most extensive temporal network analysis to date modelling the temporal dynamics between prodromal features in SMD, with respect to both the breadth of features and the large sample.

First, our findings provide evidence for the existence of dynamic relationships between prodromal features which are transdiagnostic across SMD (in the context of secondary mental health care). Understanding these dynamics can be used to identify risk states to prevent the progression to SMD onset.^39^ We identified one positive psychotic community (delusional thinking-hallucinations-paranoia) and two behavioural communities (aggression-cannabis use-cocaine use-hostility and aggression-agitation-hostility) as the most common pathways leading to SMD onset^17^. Furthermore, we found minimal differences among diagnosis-specific sub-networks, highlighting substantial overlap in the dynamic progression of prodromal stages of UMD, BMD and PSY. This finding aligns with previous findings of transdiagnostic overlap in duration, first presentation and frequencies of prodromal features across SMD,^40^ and extends them by showing transdiagnostic temporally causal relationships between these features. Echoing our earlier findings,^40^ BMD and PSY exhibited the most similar pattern of relationships between prodromal features, with UMD being the most dissimilar. These findings support the emergence of transdiagnostic early detection strategies^41^ that have previously been conceptualised in the clinical staging model^42–47^ and the Clinical High At Risk Mental State (CHARMS^48^) criteria, which have started to be implemented clinically.^48–52^

Second, insights from our diagnosis-specific sub-networks might inform the development and refinement of at-risk psychometric tools. For example, CHARMS criteria currently use assessments developed for full-threshold disorders to define UMD and BMD risk states^53,54^. These assessments could be refined to improve their specificity for at-risk populations^36^. To detect individuals at clinical high risk for psychosis (CHR-P), current assessments show low specificity^55^, which may be improved by incorporating additional features from our analyses. The prominence of positive symptoms in PSY affirms the relevance of existing assessments,^56–58^ which primarily focus on these symptoms. However, our findings also support the expansion of these tools for other SMD. In our network models, communities represent commonly occurring dynamic pathways of prodromal features, which gould guide these refinements. For example, hallucinations consistently emerge in the most frequent communities across all sub-networks but co-occur with different symptoms in each case. Specifically, in UMD, hallucinations are associated with irritability, while in BMD, they are linked to irritability along with aggression, poor insight and poor motivation. This finding aligns with the inclusion of hypomanic symptoms as diagnostic risk factors^59^ in psychometric instruments^60–62^ for bipolar at-risk states^63^. In PSY, hallucinations are linked with disturbed sleep and anxiety. Sleep issues have already been reported in in CHR-P individuals^64,65^ and those with psychosis^66,67^. Our findings extend this by providing evidence that, in the PSY prodrome, sleep disturbances may be a consequence of hallucinations, which may inform early detection strategies.

Third, our networks were densely connected with predominantly positive associations between prodromal features; presenting with one feature typically predicts the emergence, rather than absence, of the same or another feature in the future. As a result of this sequential symptom build-up, interventions need to target highly central features to disrupt the SMD prodrome network and reduce the risk of further prodromal features emerging. We have identified features with high centrality that represent potential preventive intervention targets. Typical symptoms of depression including mood instability and guilt were highly central in the UMD prodrome. Cannabis use also showed high centrality, aligning with previous research showing that cannabis consumption is associated with an increased risk of developing depression later in life.^68,69^ However, suicidality was not a central feature, likely highlighting its lower prominence during the prodrome relative to the later stages of the full disorder.^70^ For BMD, manic symptoms (e.g. elation) were highly predictive of other features, but positive psychotic symptoms (i.e. paranoia) were highly predicted by other features, whereas the reverse was observed in PSY. This finding supports the idea that developing interventions which distinctly target these symptoms may halt the progression of these disorders. Aggression and agitation were central features across all diagnosis-specific networks and part of the most prominent behavioural communities across SMD. However, this finding requires careful interpretation as our NLP algorithm for aggression does not distinguish between forms of violence directed to others or oneself, and individuals with PSY are more likely to be victims of violence than the general population.^2,71^

This study, while comprehensive, has several limitations. First, the network features are prodromal as they are detectable in secondary care before these diagnoses. However, despite our extensive range of prodromal, sociodemographic and treatment variables, there may still be unaccounted factors that influence the temporal evolution of SMD prodromes, such as functioning.^72^ While Granger causality in this study provides insights into temporal predictive relationships between prodromal features, it does not account for the underlying mechanisms or confounding factors that may drive SMD progression. Future work should investigate symptom trajectories, and aspire to construct explanatory causal models of their phenomenological and neurobiological alterations, which would enable a deeper mapping of causality.^73–75^ Second, to reduce the missingness in the dataset, we used a relatively short look-back period. However, this two-year period before disorder onset aligns with the typical duration of clinical care for at-risk individuals.^76^ Third, the final population presents a selection bias towards those receiving more frequent secondary care, limiting generalizability. Similarly, specific features, such as disorganised symptoms, may be underrepresented due to the need for consistent clinical visits. However, there were minimal differences between included and excluded individuals, in terms of demographics, clinical variables and presenting symptoms. EHR and NLP-related limitations are discussed in eLimitations 1.

Overall, our study highlights the presence of a detectable transdiagnostic SMD prodrome by modelling the evolution of symptoms and substance use over time. Our findings support the potential for broader transdiagnostic early detection services for SMD that provide preventive care to individuals at-risk and a research platform for investigating putative interventions.

## Data Availability

The data accessed by CRIS remain within an NHS firewall and governance is provided by a patient-led oversight committee. Subject to these conditions, data access is encouraged and those interested should contact Robert Stewart, CRIS academic lead. There is no permission for data sharing. Covariance matrices to estimate networks and all analysis code are available on GitHub: https://github.com/m-arribas/network_analysis.git.

## Data Sharing Statement

The data accessed by CRIS remain within an NHS firewall and governance is provided by a patient-led oversight committee. Subject to these conditions, data access is encouraged and those interested should contact Robert Stewart, CRIS academic lead. Further details regarding the CRIS platform can be found elsewhere^18^. There is no permission for data sharing. Covariance matrices to estimate networks and all analysis code are available on GitHub: https://github.com/m-arribas/network_analysis.git.

## Ethics committee approval

Permissions for the study were granted by the Oxfordshire Research Ethics Committee C; because the data set comprised deidentified data, informed consent was not required^18^.

## Authors’ contribution

MA, JMB and DO conceptualised the study. DO and PFP supervised the study. MA, JMB and DO ran the statistical analyses. All authors drafted, edited, and approved the final version of the manuscript.

