## Supplementary Material for "Longitudinal evolution of the transdiagnostic prodrome to severe mental disorders: a dynamic temporal network analysis informed by natural language processing and electronic health records"

### eMethods 1 SLaM Service Characteristics

With respect to service characteristics, SLaM early intervention teams serve an overall catchment area of 443,050 people aged 16–35 years (2017)[1], and are amongst the largest of their kinds in the UK and worldwide. Incidence of psychosis in SLaM (from 58.3 to 71.9 cases per 100,000 person-years[1]) is greater than the national average, and it is one of the highest worldwide[2]. This may be accounted for by accumulation of several risk factors for psychosis such as immigration, higher numbers of people from ethnic minorities associated with psychosis (e.g., 50% of SLaM population is from African/Afro-Caribbean backgrounds) and illicit substance misuse[3–5].

The trust is digitized and paper-free[6], with each patient having a personal EHR since 2007[7,8]. SLaM healthcare professionals are legally required to update these records[6]. The SLaM register contains all these clinical records which are constantly updated throughout the patient's care, irrespective of any discharges from and/or referrals to other services.

The CRIS data resource received ethical approval as an anonymized dataset for secondary analyses from Oxfordshire REC C (Ref: 18/SC/0372).

eTable 1 Operationalisation of ICD-10 diagnoses employed in the current study (https://icd.who.int/browse10/), stratified by SMD group and excluded diagnoses (including organic disorders, non-SMD and unspecified mental disorders).

| **Diagnostic group** | **Specific ICD-10 code** | **Specific ICD-10 diagnosis, version 2016** |
|---|---|---|
| Unipolar mood disorders (UMD) | F32.x (excluding F32.3) | Non-psychotic depressive episode |
| | F33.x (excluding F33.3) | Non-psychotic recurrent depressive disorder |
| | F34.x (excluding F34.0) | Persistent unipolar mood disorders |
| | F34.8, F34.9, F38, F39, F38.x, F39.x | Unspecified mood disorders |
| Bipolar mood disorders (BMD) | F30.x (excluding *.2) | Non-psychotic manic episode |
| | F31.x (excluding F31.2 and F31.5) | Non-psychotic bipolar affective disorder |
| | F34.0 | Cyclothymia |
| Psychotic disorders (PSY) | [F10-F19] .4, [F10-F19] .5, [F10-F19] .7 | Mental and behavioural disorders due to psychoactive substance use with psychotic symptoms or delirium |
| | F20-29 | Schizophrenia, schizotypal and delusional disorders |
| | F30.2 | Mania with psychotic symptoms |
| | F31.2 | Bipolar affective disorder, current episode manic with psychotic symptoms |
| | F31.5 | Bipolar affective disorder, current episode severe depression with psychotic symptoms |
| | F32.3 | Severe depressive episode with psychotic symptoms |
| | F33.3 | Recurrent depressive disorder, current episode severe with psychotic symptoms |
| | F53.1 | Severe mental and behavioural disorders associated with the puerperium, not elsewhere classified (post-partum psychosis) |
| Excluded | F00-F09 | Organic, including symptomatic, mental disorders |
| | F10-19 (excluding *.4, *.5 and *.7) | Mental and behavioural disorders due to psychoactive substance use |
| | F40-48 | Neurotic, stress-related and somatoform disorders |
| | F50-59 (excluding F53.1) | Behavioural syndromes associated with physiological disturbances and physical factors |
| | F60-69 | Disorders of adult personality and behaviour |
| | F70-79 | Mental retardation |
| | F80-89 | Disorders of psychological development |
| | F90-98 | Behavioural and emotional disorders with onset usually occurring in childhood and adolescence |
| | F99 | Unspecified mental disorder |

eTable 2 Definition of self-assigned ethnicity according to UK Office of National Statistics

| **Ethnic group** | **Self-assigned ethnicity as recorded in EHR** |
|---|---|
| Black | Black or Black British - African<br>Black or Black British - Caribbean<br>Black or Black British - Any other Black background |
| White | White - British<br>White - Irish<br>White - Any other White background |
| Asian | Asian or Asian British - Bangladeshi<br>Asian or Asian British - Indian<br>Asian or Asian British - Pakistani<br>Asian or Asian British - Any other Asian background<br>Other Ethnic Groups - Chinese |
| Mixed | Mixed - White and Asian<br>Mixed - White and Black African<br>Mixed - White and Black Caribbean<br>Mixed - Any other mixed background |
| Other | Other Ethnic Groups - Any other ethnic group |
| Missing | Not Known<br>Not Recorded |

eTable 3 Medication Classification

| Antipsychotics | Antidepressants | Mood Stabilisers | Anxiolytics |
|---|---|---|---|
| Amisulpride (Solian) | Agomelatine (Valdoxan) | Carbamazepine (Carbagen, Tegretol) | Alprazolam |
| Aripiprazole (Abilify) | Amitriptyline (Triptafen) | Desitrend | Bio-Melatonin |
| Asenapine (Sycrest) | Amitriptyline hydrochloride | Lamotrigine (Lamictal) | Buspirone |
| Benperidol | Buproprion (Wellbutrin, Zyban) | Levetiracetam (Keppra) | Buspirone hydrochloride |
| Cariprazine (Reagila) | Citalopram (Cipramil) | Lithium Carbonate (Camcolit, Liskonum, Priadel) | Chloral Hydrate |
| Chlorpromazine (Thorazine, Largactil) | Citalopram Hydrobromide | Lithium Citrate (Priadel) | Chlordiazepoxide |
| Clozapine (Clozaril) | Citalopram Hydrochloride | Pregabalin (Alzain, Lecaent, Lyrica, Rewisca) | Chlorhexidine |
| Denzapine | Clomipramine (Anafranil) | Sodium Valproate (Convulex, Epilim, Epilim Chronosphere, Episenta, Epivalk, Orlept) | Clomethiazole |
| Droperidol | Dosulepin (Dothiepin, Prothiaden) | Topiramate (Topamax) | Clonazepam |
| Flupentixol Decanoate (Psytixol) | Doxepin | Valproic Acid (Valproate) | Diazepam |
| Flupentixol Hydrochloride (Depixol) | Duloxetine (Cymbalta, Duciltia) | | Diazepam Rectube |
| Fluphenazine | Escitalopram (Cipralex) | | Flurazepam |
| Fluphenazine Decanoate (Modecate) | Fluoxetine (Olena, Oxactin, Prozac, Prozep) | | Loprazolam |
| Fluphenazine Hydrochloride | Flupentixol (Fluanxol) | | Lorazepam |
| Haloperidol (Haldol, Seranace) | Flupentixol Hydrochloride (Depixol) | | Lormetazepam |
| Haloperidol Decanoate | Fluvoxamine (Faverin) | | Melatonin |
| Levomepromazine (Nozinan) | Imipramine | | Nitrazepam |
| Levomepromazine Hydrochloride | Isocarboxazid | | Oxazepam |
| Levomepromazine Maleate (Levinan) | Lofepramine | | Rivotril |
| Lurasidone (Latuda) | Maprotiline | | Temazepam |
| Melperone | Mianserin | | |
| Olanzapine (Zalasta, Zypadhera, Zyprexa) | Mirtazapine (Zispin) | | |
| Olanzapine embonate | Moclobemide (Manerix) | | |
| Paliperidone (Invega, Trevicta, Xeplion) | Nefazodone | | |
| Penfluridol | Nortriptyline | | |
| Pericyazine | Opipromal (Insidon) | | |
| Perphenazine (Fentazin) | Paroxetine (Seroxat) | | |
| Pimozide (Orap) | Reboxetine (Edronax) | | |

| | |
|---|---|
| Pipotiazine (Piportil) | Sertraline (Lustral) |
| Pipotiazine palmitate | Tianeptine (Coaxil, Stablon) |
| Promazine | Tranylcypromine |
| Quetiapine (Atrolak, Biquelle, Brancico, Ebesque, Mintreleq, Neotiapim, Psyquet, Seroquel, Sondate, Zaluron) | Trazodone (Molipaxin) |
| Risperidone (Risperdal) | Trimipramine (Surmontil) |
| Sulpiride (Dolmatil) | Tryptophan (Optimax) |
| Thioridazine (Melleril) | Venlafaxine (Alventa, Amphero, Depefex, Efexor, Politid, Rodomel, Sunveniz, Tonpular, Venax, Vencarm, Venlablue, Venladex, Venlalic, Vensir, ViePax, Winfex) |
| Trifluoperazine (Stelazine) | Vortioxetine (Brintellix) |
| Zaponex | |
| Ziprasidone (Geodon) | |
| Zuclopenthixol (Clopixol, Clopixol Acuphase) | |
| Zuclopenthixol Acetate | |
| Zuclopenthixol Decanoate | |
| Zuclopenthixol Hydrochloride | |

### eMethods 2 NLP algorithm development and validation

The CRIS symptom algorithms (e.g. 'guilt') have been developed using machine learning approaches against gold standard training sets manually annotated for positive, negative and unknown (irrelevant) mentions. As such, they are able to exclude language features such as negation (e.g. 'patient denies guilt', 'patient has no guilt') and irrelevant mentions (e.g. 'his mother felt guilty'). Patterns of failure driving false positives are identified through manual testing of algorithm output (e.g. 'ZZZZZ was found guilty of stealing'); the machine learning classifier is then trained on these false positives to ignore these and similar statements in an iterative process of testing and redeveloping until acceptable precision is achieved. Patterns of failure identified through testing can be found in the CRIS service's comprehensive online NLP algorithm library provided at https://www.maudsleybrc.nihr.ac.uk/facilities/clinical-record-interactive-search-cris/cris-natural-language-processing/.

The performance of each NLP algorithm was measured with precision (proportion of true positive instances of total NLP-labelled positive instances) and recall (proportion of true positive instances of all positive instances in the text). As EHRs provide multiple opportunities for term detection, we favour precision over recall, using only NLP algorithms with at least 80% precision (see eTable 4 for a final list of NLP algorithms employed). Precision was defined as the ratio of the number of relevant (true positive) instances retrieved out of the total NLP-labelled positive instances (including irrelevant [false positive] and relevant [true positive] instances) in human-annotated EHR.

These algorithms were manually validated by an independent researcher at the SLaM Biomedical Research Centre Nucleus prior to the current research project. The programme for algorithms validation was responsive to the specific needs of scheduled CRIS research activities and therefore the approach was not standardised. For example, depression symptom algorithms have been validated against records for SLaM individuals who had ever had a depression diagnosis; other algorithms have been validated against records for all individuals on the SLaM register.

eTable 4 Type and precision for 61 NLP algorithms

Precision values are taken from the CRIS Natural Language Processing Library (2021) [9], and were obtained by randomly selecting *n* positive annotations from each algorithm for a specified cohort, limited to one annotation per patient ID. Precision was then calculated as the ratio of the number of relevant (true positive) instances retrieved out of the total NLP-labelled positive instances (including irrelevant [false positive] and relevant [true positive] instances) for each NLP algorithm.

| NLP algorithms | Cohort | Annotations validated (n) | Precision (%) |
|---|---|---|---|
| Aggression | Random sample | 100 | 90 |
| Agitation | Random sample | 100 | 85 |
| Anergia | Random sample | 100 | 84 |
| Anhedonia | Random sample | 100 | 94 |
| Anxiety | Random sample | 100 | 94 |
| Apathy | Random sample | 100 | 94 |
| Arousal | Random sample | 100 | 89 |
| Bad dreams | CAMHS events | 100 | 92 |
| Blunted affect | Random sample | 100 | 98 |
| Cannabis use | All patients | 100 | 88 |
| Circumstantiality | Random sample | 100 | 97 |
| Cocaine use | Random sample | 30 | 97 |
| Cognitive impairment | Patients with F20 | 100 | 84 |
| Concrete thinking | Random sample | 146 | 91 |
| Delusional thinking | Random sample | 100 | 90 |
| Derailment | Random sample | 100 | 87 |
| Disturbed sleep | Random sample | 100 | 89 |
| Diurnal mood | Random sample | 100 | 86 |
| Early morning wakening | Random sample | 100 | 96 |
| Echolalia | Random sample | 100 | 96 |
| Elation | Random sample | 100 | 95 |
| Emotional withdrawal | Random sample | 100 | 87 |
| Feeling helpless | Random sample | 100 | 92 |
| Feeling hopeless | Random sample | 100 | 88 |
| Feeling lonely | Random sample | 100 | 87 |
| Feeling worthless | Random sample | 100 | 91 |
| Flight of ideas | Random sample | 100 | 89 |
| Formal thought disorder | Random sample | 100 | 85 |
| Grandiosity | Random sample | 100 | 89 |
| Guilt | Random sample | 100 | 84 |
| Hallucinations (all) | Random sample | 100 | 90 |
| Hostility | Random sample | 100 | 86 |
| Insomnia | Random sample | 100 | 97 |
| Irritability | Random sample | 100 | 99 |
| Loss of coherence | Random sample | 158 | 85 |

| | | | |
|---|---|---|---|
| Low energy | CAMHS events | 100 | 89 |
| MDMA use | Random sample | 100 | 94 |
| Mood instability | Random sample | 100 | 91 |
| Mutism | Random sample | 100 | 95 |
| Negative symptoms | Random sample | 100 | 87 |
| Nightmares | Random sample | 100 | 89 |
| Paranoia | Random sample | 100 | 89 |
| Passivity | Random sample | 100 | 88 |
| Poor appetite | Random sample | 100 | 89 |
| Poor concentration | Random sample | 100 | 88 |
| Poor insight | Random sample | 100 | 85 |
| Poor motivation | Random sample | 100 | 95 |
| Poverty of speech | Random sample | 100 | 88 |
| Poverty of thought | Random sample | 100 | 98 |
| Social withdrawal | Random sample | 100 | 98 |
| Stupor | Random sample | 100 | 88 |
| Suicidality | CAMHS events | 100 | 87 |
| Tangential speech | Random sample | 100 | 90 |
| Tearfulness | Random sample | 100 | 94 |
| Thought block | Random sample | 100 | 92 |
| Thought broadcast | Random sample | 100 | 84 |
| Thought insertion | Random sample | 100 | 84 |
| Thought withdrawal | Random sample | 100 | 84 |
| Tobacco use | Random sample | 118 | 90 |
| Waxy flexibility | Random sample | 100 | 81 |
| Weight loss | Random sample | 100 | 80 |

### eMethods 3 Sensitivity Analysis

In a sensitivity analysis, to test for any sampling bias in the final population, we compared excluded individuals (with four or fewer follow-up intervals) to those included (with five or more follow-up intervals) on sociodemographic variables (age, gender, self-assigned ethnicity), clinical variables (proportion of individuals belonging to each SMD group and medication prescriptions at index), as well as the severity of presenting features (frequency of prodromal clusters within antecedent period).

eMethods 4 Pre-processing

All pre-processing steps were included in both the main analysis and bootstrap analysis to allow for any error in the pre-processing stage to be considered within stability checks. Moreover, imputation was carried out before and separate from the model fitting process for more transparency. For all numerical missing data (EHR entry length, EHR entry frequency and feature occurrence at each follow-up interval), we imputed values using linear interpolation as this method has shown higher imputation performance compared to non-linear imputation methods.[10] We implemented it using the 'imputeTS' package[11] (version 3.3) with the function 'na_interpolation'.

For all categorical missing data (gender and self-assigned ethnicity), we imputed values using multivariate imputation (m=1) by chained equations, implemented in the 'MICE' package[12] (version 3.15.0), using the method polytomous regression imputation, 'polyreg'. For the latter imputation, we used demographics variables (age, medication variables [prescription of antipsychotics, antidepressants, mood stabilisers or anxiolytics at index]), as well as auxiliary demographics (borough of residence) to improve MICE algorithm stability of the target variables of interest. These were selected due to their theoretical relationship to our variables of interest.

To control for confounders, we regressed out age ($Age$; continuous), self-assigned ethnicity (*Ethnicity;* categorical), sex ($Sex$; categorical) and whether a participant was receiving a prescription of antipsychotics, antidepressants, mood stabilisers or anxiolytics at index (*Antipsychotics*, *Antidepressants*, *Mood stabilisers*, *Anxiolytics;* binary). To control for the frequency and intensity of clinical contacts within each follow-up interval, we also regressed out the frequency (*EHR entry frequency* $^t$: number of entries within each follow-up interval; continuous) and length of EHR entries (*EHR entry length* $^t$*:* total number of words recorded across all EHR entries within each follow-up interval; continuous) (See Eq. 1):

(Eq. 1)

$$\bar{V}_i^t = \alpha + (\beta_1 * Age^t) + (\beta_2 * Ethnicity) + (\beta_3 * Sex) + (\beta_4 * Antipsychotics) + (\beta_5 * Antidepressants) + (\beta_6 * Mood\ Stabilisers) + (\beta_7 * Anxiolytics) + (\beta_8 * EHR\ entry\ frequency^t) + (\beta_9 * EHR\ entry\ length^t) + \varepsilon_i^t$$

$$\varepsilon_i^t = V_i^t - \bar{V}_i^t$$

The resulting residuals ($\varepsilon_i^t$) from each equation (observed values [$V_i^t$] minus predicted values [$\bar{V}_i^t$], for each variable [$i$], at each follow-up interval [$t$]) were then retained as nodes for the models. We then applied scaling (using the 'scale' function in R) to $\varepsilon_i^t$ to ensure all nodes and edges were assessed on the same dimension and any mean trends removed. This approach helps to understand how prodromal features vary with time, both individually and in relation to each other, regardless of whether the symptoms are generally getting worse over time. Detrended and normalised between-individual residuals over time were visualised graphically and are presented in eFigure 1.

Variables with near-zero variance were then excluded from the analysis to reduce network instability[13]. To determine the variables to include in each network, the frequency ratio (ratio of the frequency of the most common to the second most common value) was calculated for each feature at each follow-up interval in the imputed datasets (see eTable 7). Features with at least one follow-up interval with a large frequency ratio (greater than 20) were considered to have near-zero variance and were subsequently excluded to reduce network instability.[13]

### eMethods 5 Centrality Measure definitions

From each matrix, the strength of connections between features (edge weights, $z$) was estimated as directed partial correlation coefficients (temporal) or partial correlation coefficients (contemporaneous/between-subject). In temporal networks, edges were categorised into 3 types: autocorrelative (node predicts itself in the next time point), unidirectional (node predicts another, without reciprocation) and bidirectional (mutual prediction between two nodes). Degree centrality were extracted from each graph. For temporal networks, centrality was defined as the sum of absolute (directed) edge weights in (in-centrality, $c_{in}$) and out (out-centrality, $c_{out}$) of a node (excluding autocorrelative edges). For contemporaneous and between-subject networks, centrality was defined as the sum of absolute (undirected) edge weights for a node (autocorrelative edges do not exist).

eTable 5 Index diagnoses across the final sample (N=6,462)

| **Index diagnosis** | **UMD (N=2,066)** | **BMD (N=740)** | **PSY (N=3,656)** |
|---|---|---|---|
| F32.x (excluding F32.3) | 1112 | 0 | 0 |
| F33.x (excluding F33.3) | 750 | 0 | 0 |
| F34.x (excluding F34.0) | 79 | 0 | 0 |
| F34.8, F34.9, F38, F39, F38.x, F39.x | 125 | 0 | 0 |
| F30.x (excluding *.2) | 0 | 21 | 0 |
| F31.x (excluding F31.2 and F31.5) | 0 | 694 | 0 |
| F34.0 | 0 | 25 | 0 |
| [F10-F19].4, [F10-F19].5, [F10-F19].7 | 0 | 0 | 56 |
| F20-29 | 0 | 0 | 3205 |
| F30.2 | 0 | 0 | 14 |
| F31.2 | 0 | 0 | 128 |
| F31.5 | 0 | 0 | 22 |
| F32.3 | 0 | 0 | 154 |
| F33.3 | 0 | 0 | 74 |
| F53.1 | 0 | 0 | 3 |

eTable 6 Raw counts of each NLP feature in total samples (N=6,462)

| Feature | Follow-up interval | | | | | |
|---|---|---|---|---|---|---|
| | **1** | **2** | **3** | **4** | **5** | **6** |
| Aggression | 666 | 750 | 826 | 762 | 705 | 749 |
| Agitation | 770 | 806 | 806 | 853 | 832 | 837 |
| Anergia | 18 | 14 | 24 | 27 | 16 | 16 |
| Anhedonia | 127 | 161 | 165 | 139 | 127 | 106 |
| Anxiety | 2671 | 3049 | 3073 | 3076 | 3024 | 2929 |
| Apathy | 48 | 77 | 59 | 54 | 50 | 58 |
| Arousal | 240 | 247 | 246 | 248 | 237 | 261 |
| Bad dreams | 41 | 44 | 44 | 27 | 37 | 34 |
| Blunted affect | 101 | 148 | 126 | 107 | 112 | 119 |
| Cannabis use | 595 | 692 | 684 | 694 | 664 | 679 |
| Circumstantiality | 60 | 74 | 60 | 50 | 55 | 55 |
| Cocaine use | 328 | 395 | 361 | 377 | 343 | 347 |
| Cognitive impairment | 2378 | 2683 | 2756 | 2692 | 2663 | 2620 |
| Concrete thinking | 15 | 24 | 21 | 18 | 20 | 24 |
| Delusional thinking | 593 | 660 | 612 | 601 | 659 | 618 |
| Derailment | 26 | 21 | 17 | 26 | 25 | 21 |
| Disturbed sleep | 1356 | 1505 | 1554 | 1474 | 1456 | 1391 |
| Diurnal mood | 29 | 22 | 21 | 26 | 27 | 24 |
| Early morning wakening | 43 | 51 | 47 | 45 | 43 | 23 |
| Echolalia | 4 | 6 | 5 | 3 | 3 | 2 |
| Elation | 245 | 251 | 248 | 240 | 250 | 251 |
| Emotional withdrawal | 292 | 336 | 311 | 289 | 266 | 266 |
| Feeling helpless | 98 | 85 | 111 | 121 | 116 | 92 |
| Feeling hopeless | 327 | 421 | 390 | 370 | 371 | 368 |
| Feeling lonely | 274 | 293 | 290 | 291 | 285 | 272 |
| Feeling worthless | 117 | 131 | 111 | 107 | 113 | 99 |
| Flight of ideas | 94 | 115 | 94 | 95 | 104 | 106 |
| Formal thought disorder | 27 | 30 | 28 | 30 | 24 | 44 |
| Grandiosity | 184 | 210 | 183 | 177 | 191 | 189 |
| Guilt | 356 | 406 | 415 | 378 | 375 | 355 |
| Hallucinations (all) | 757 | 842 | 841 | 825 | 830 | 845 |

| | | | | | | |
|---|---|---|---|---|---|---|
| Hostility | 284 | 309 | 337 | 310 | 313 | 312 |
| Insomnia | 248 | 301 | 283 | 270 | 263 | 246 |
| Irritability | 636 | 695 | 696 | 681 | 688 | 673 |
| Loss of coherence | 72 | 78 | 88 | 96 | 75 | 92 |
| Low energy | 222 | 250 | 281 | 235 | 199 | 193 |
| MDMA use | 15 | 19 | 12 | 13 | 14 | 17 |
| Mood instability | 620 | 666 | 698 | 690 | 638 | 611 |
| Mutism | 71 | 73 | 68 | 58 | 59 | 66 |
| Negative symptoms | 94 | 121 | 136 | 135 | 128 | 129 |
| Nightmares | 223 | 242 | 253 | 214 | 205 | 213 |
| Paranoia | 1117 | 1290 | 1280 | 1233 | 1276 | 1265 |
| Passivity | 31 | 54 | 58 | 45 | 43 | 37 |
| Poor appetite | 183 | 237 | 218 | 167 | 179 | 194 |
| Poor concentration | 678 | 770 | 698 | 683 | 598 | 619 |
| Poor insight | 703 | 799 | 798 | 758 | 756 | 776 |
| Poor motivation | 362 | 407 | 419 | 397 | 387 | 371 |
| Poverty of speech | 24 | 30 | 22 | 24 | 27 | 30 |
| Poverty of thought | 16 | 22 | 11 | 12 | 16 | 15 |
| Social withdrawal | 87 | 110 | 112 | 90 | 64 | 97 |
| Stupor | 6 | 3 | 8 | 6 | 2 | 4 |
| Suicidality | 419 | 480 | 469 | 426 | 422 | 383 |
| Tangential speech | 136 | 145 | 109 | 112 | 116 | 123 |
| Tearfulness | 751 | 874 | 836 | 788 | 773 | 686 |
| Thought block | 42 | 60 | 36 | 34 | 41 | 37 |
| Thought broadcast | 42 | 52 | 42 | 53 | 47 | 52 |
| Thought insertion | 42 | 53 | 54 | 45 | 49 | 55 |
| Thought withdrawal | 19 | 24 | 24 | 21 | 22 | 19 |
| Tobacco use | 550 | 624 | 595 | 627 | 636 | 646 |
| Waxy flexibility | 0 | 0 | 0 | 0 | 0 | 1 |
| Weight loss | 355 | 396 | 366 | 345 | 370 | 359 |

eResults 1 Sensitivity Analysis to test for sampling bias in the final population. Individuals excluded from the analysis (with four or fewer follow-up intervals) were compared to the included population (with five or more follow-up intervals) on (A) sociodemographic and clinical variables, as well as (B) severity of presenting features

A. Sociodemographic and clinical variables (N=22,628)

| Variables | | Inclusions (N=6,462) | Exclusions (N=16,166) |
|---|---|---|---|
| | | Mean(SD)/ n(%) | Mean(SD) n(%) |
| Index diagnostic group | BMD | 740 (11.45%) | 1374 (8.50%) |
| | UMD | 2066 (31.97%) | 8746 (54.10%) |
| | PSY | 3656 (56.58%) | 6046 (37.40%) |
| Age | | 43.5 (16.1) | 41.0 (17.8) |
| Gender | Female | 3321 (51.39%) | 9105 (56.32%) |
| | Male | 3138 (48.56%) | 7048 (43.60%) |
| | Other | 3 (0.05%) | 9 (0.06%) |
| | missing | NA | 4 (0.02%) |
| Self-assigned ethnicity | Asian | 459 (7.10%) | 986 (6.10%) |
| | Black | 1906 (29.50%) | 3927 (24.29%) |
| | Mixed | 209 (3.23%) | 509 (3.15%) |
| | Other | 223 (3.45%) | 995 (6.15%) |
| | White | 3536 (54.72%) | 8268 (51.14%) |
| | missing | 129 (2.00%) | 1481 (9.16%) |
| Antidepressants | 1 | 2537 (39.26%) | 5201 (32.17%) |
| Anxiolytics | 1 | 1569 (24.28%) | 2672 (16.53%) |
| Antipsychotics | 1 | 3430 (53.08%) | 4727 (29.24%) |
| Mood stabilisers | 1 | 1078 (16.68%) | 1283 (7.94%) |

B. Presenting symptom clusters (N=20,120). For each individual the mean frequency of symptom clusters were calculated in the antecedent period (6 month period prior to index date). Cohens D were computed to compare between inclusion vs exclusion population.

| | **Inclusion (N=6,257)** | **Exclusions (N=13,863)** | **Cohens D** | **Cohens D Effect Size** | **CI low** | **CI high** |
|---|---|---|---|---|---|---|
| **Positive** | 0.77 | 0.88 | -0.08 | Negligible | -0.11 | -0.05 |
| **Negative** | 0.14 | 0.19 | -0.13 | Negligible | -0.16 | -0.10 |
| **Depressive** | 0.55 | 0.85 | -0.29 | Small | -0.31 | -0.26 |
| **Manic** | 0.40 | 0.52 | -0.17 | Negligible | -0.20 | -0.14 |
| **Disorganised** | 0.12 | 0.17 | -0.14 | Negligible | -0.17 | -0.11 |
| **Catatonic** | 0.01 | 0.01 | -0.03 | Negligible | -0.06 | -0.01 |
| **Other** | 0.80 | 0.99 | -0.22 | Small | -0.24 | -0.19 |

| | | | | | | |
|---|---|---|---|---|---|---|
| **Substance use** | 0.19 | 0.21 | -0.05 | Negligible | -0.08 | -0.02 |

eTable 7 Near-zero variance classification (SMD network)

Frequency ratio was defined as the ratio between the frequency of the most common value to the second most common value. A feature was considered to be near zero variance if it had a frequency ratio value >20 in at least one follow-up interval. Near-zero variance features are highlighted in bold. The final 23 prodromal features used were: aggression, agitation, anxiety, cannabis use, cocaine use, cognitive impairment, delusional thinking, disturbed sleep, emotional withdrawal, feeling hopeless, guilt, hallucinations (all), hostility, irritability, mood instability, paranoia, poor concentration, poor insight, poor motivation, suicidality, tearfulness, tobacco use, and weight loss.

| | **Follow-up** interval | | | | | |
|---|---|---|---|---|---|---|
| **Feature** | **1** | **2** | **3** | **4** | **5** | **6** |
| Aggression | 8.2 | 7.5 | 6.8 | 7.4 | 8.2 | 7.6 |
| Agitation | 6.9 | 7 | 6.9 | 6.5 | 6.7 | 6.6 |
| **Anergia** | 338.7 | 460.1 | 268 | 238.1 | 402.4 | 402.4 |
| **Anhedonia** | 48.7 | 38.8 | 37.9 | 45.4 | 49.8 | 59.3 |
| Anxiety | 1.2 | 1.1 | 1.1 | 1.1 | 1.1 | 1.1 |
| **Apathy** | 128.1 | 82.8 | 108.4 | 118.5 | 128.1 | 108.4 |
| **Arousal** | 25 | 25 | 25 | 24.9 | 26.2 | 23.3 |
| **Bad dreams** | 152.7 | 145.7 | 145.7 | 238.1 | 173.5 | 188.9 |
| **Blunted affect** | 59.3 | 42 | 50.2 | 59.3 | 56.6 | 52.8 |
| Cannabis use | 9.3 | 8.3 | 8.4 | 8.2 | 8.7 | 8.4 |
| **Circumstantiality** | 103.1 | 86.2 | 106.6 | 128.1 | 116.4 | 116.4 |
| Cocaine use | 17.7 | 15.3 | 16.9 | 16.1 | 17.8 | 17.3 |
| Cognitive impairment | 1.5 | 1.4 | 1.3 | 1.4 | 1.4 | 1.4 |
| **Concrete thinking** | 402.4 | 268 | 306.4 | 357.6 | 321.8 | 247.3 |
| Delusional thinking | 9.4 | 8.7 | 9.5 | 9.7 | 8.7 | 9.3 |
| **Derailment of speech** | 221.6 | 306.4 | 378.7 | 247.3 | 257.2 | 306.4 |
| Disturbed sleep | 3.5 | 3.3 | 3.1 | 3.3 | 3.4 | 3.6 |
| **Diurnal mood** | 207.2 | 292.4 | 321.8 | 247.3 | 238.1 | 268 |
| **Early morning wakening** | 139.3 | 125.6 | 136.3 | 142.4 | 149.1 | 268 |
| **Echolalia** | 1612.8 | 1074.8 | 1290 | 2150.7 | 2150.7 | 3226.5 |
| **Elation** | 23.9 | 24.5 | 24.9 | 25.8 | 24.7 | 24.5 |
| Emotional withdrawal | 20 | 18 | 19.7 | 21.3 | 23.2 | 23 |
| **Feeling helpless** | 62.9 | 74.9 | 57.2 | 52.3 | 54.6 | 66.2 |
| Feeling hopeless | 17.5 | 14.3 | 15.5 | 16.5 | 16.4 | 16.3 |
| **Feeling lonely** | 21.6 | 21 | 21.1 | 21.3 | 21.6 | 22.4 |
| **Feeling worthless** | 51.9 | 48.3 | 57.2 | 59.9 | 56.1 | 63.5 |
| **Flight of ideas** | 62.9 | 54.6 | 66.9 | 66.9 | 61.1 | 59.9 |
| **Formal thought disorder** | 229.5 | 214.2 | 229.5 | 214.2 | 268 | 145.7 |
| **Grandiosity** | 33 | 29.4 | 34.1 | 35.3 | 32.6 | 33 |
| Guilt | 16 | 14.7 | 14.4 | 16.1 | 16.2 | 17 |
| Hallucinations (all) | 7 | 6.5 | 6.6 | 6.8 | 6.7 | 6.6 |
| Hostility | 20.9 | 19.8 | 18 | 19.7 | 19.5 | 19.6 |
| **Insomnia** | 24.2 | 20.3 | 21.7 | 22.9 | 23.4 | 25.1 |

| | | | | | | |
|---|---|---|---|---|---|---|
| Irritability | 8.6 | 8.2 | 8.2 | 8.4 | 8.3 | 8.5 |
| **Loss of coherence** | 83.9 | 81.8 | 70.7 | 65.5 | 85.1 | 68.4 |
| **Low energy** | 26.8 | 24.8 | 21.9 | 26.5 | 31.4 | 32.1 |
| **MDMA use** | 402.4 | 338.7 | 536.9 | 495.5 | 460.1 | 357.6 |
| Mood instability | 8.9 | 8.6 | 8.2 | 8.3 | 9.1 | 9.4 |
| **Mutism** | 89.9 | 87.4 | 93.9 | 110.3 | 108.4 | 93.9 |
| **Negative symptoms** | 63.5 | 51.9 | 46.5 | 46.8 | 49.4 | 48.3 |
| **Nightmares** | 27.6 | 25.7 | 24.5 | 29.2 | 30.5 | 28.7 |
| Paranoia | 4.5 | 4 | 4 | 4.2 | 4 | 4 |
| **Passivity** | 207.2 | 118.5 | 110.3 | 142.4 | 149.1 | 173.5 |
| **Poor appetite** | 33.2 | 26.1 | 28.5 | 37.4 | 35.1 | 32.1 |
| Poor concentration | 8.1 | 7.4 | 8.2 | 8.5 | 9.7 | 9.3 |
| Poor insight | 7.7 | 7 | 7 | 7.4 | 7.5 | 7.3 |
| Poor motivation | 15.9 | 14.8 | 14.3 | 15.3 | 15.7 | 16.2 |
| **Poverty of speech** | 257.2 | 214.2 | 292.4 | 268 | 238.1 | 200.7 |
| **Poverty of thought** | 378.7 | 292.4 | 585.8 | 536.9 | 402.4 | 429.3 |
| **Social withdrawal** | 69.9 | 57.7 | 56.6 | 70.7 | 99.9 | 65.5 |
| **Stupor** | 1074.8 | 2150.7 | 805.9 | 1074.8 | 3226.5 | 1612.8 |
| Suicidality | 13.6 | 12.4 | 12.7 | 14.2 | 14.3 | 15.7 |
| **Tangential speech** | 45.4 | 43.2 | 58.2 | 56.6 | 54.2 | 51.5 |
| Tearfulness | 7 | 6.4 | 6.7 | 7.2 | 7.3 | 8.3 |
| **Thought block** | 149.1 | 103.1 | 173.5 | 188.9 | 156.4 | 173.5 |
| **Thought broadcast** | 152.7 | 120.8 | 152.7 | 120.8 | 136.3 | 123.1 |
| **Thought insertion** | 145.7 | 120.8 | 118.5 | 142.4 | 130.7 | 114.3 |
| **Thought withdrawal** | 321.8 | 268 | 268 | 306.4 | 292.4 | 338.7 |
| Tobacco use | 10.1 | 9.3 | 9.8 | 9.2 | 9.1 | 8.9 |
| **Waxy flexibility** | 0 | 0 | 0 | 0 | 0 | 6454 |
| Weight loss | 16.4 | 15.2 | 16.6 | 17.7 | 16.4 | 16.9 |

eFigure 1 (A) Raw values and (B) detrended and normalised aggregated residuals over time (controlled for confounder variables (age, self-assigned ethnicity, sex, prescription of antipsychotics, antidepressants, mood stabilisers or anxiolytics at index, frequency and length of EHR entries). Violin plots show the distribution of values for the whole study sample at each follow-up interval. Red dots are means across participants for each time interval. Plots demonstrate no linear trend in mean values across all time intervals.

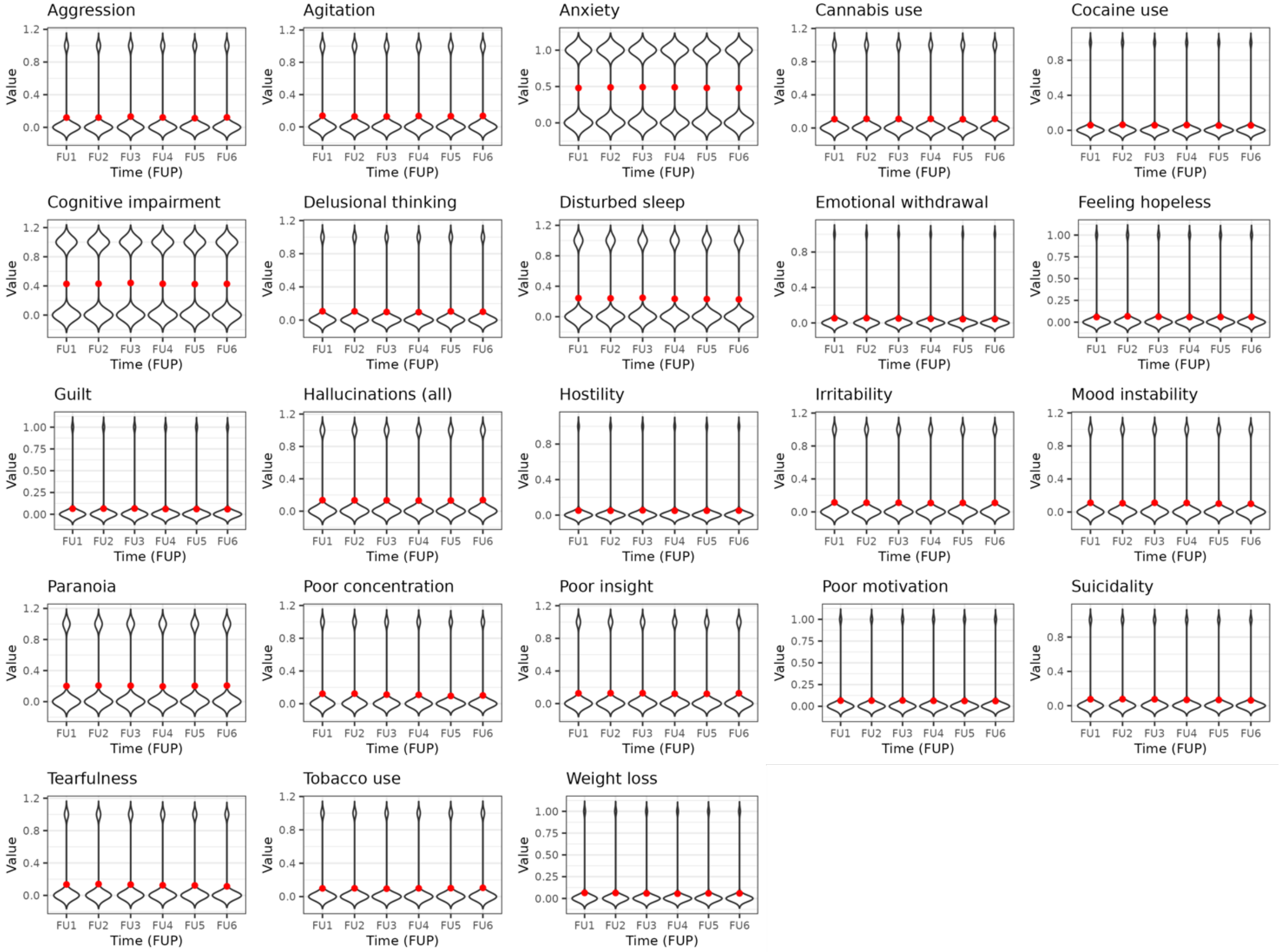

(A)

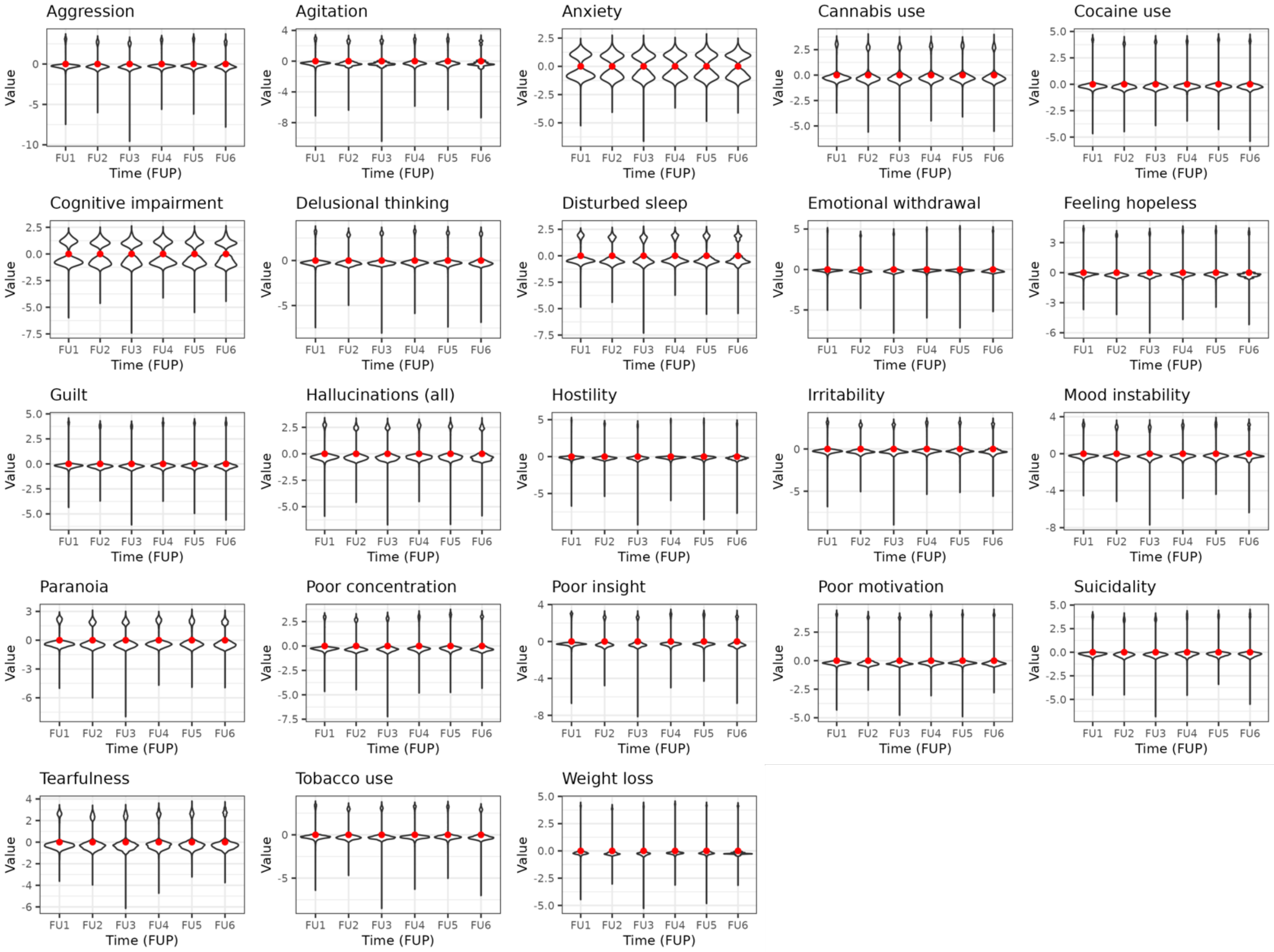

Aggression
Agitation
Anxiety
Cannabis use
Cocaine use
Cognitive impairment
Delusional thinking
Disturbed sleep
Emotional withdrawal
Feeling hopeless
Guilt
Hallucinations (all)
Hostility
Irritability
Mood instability
Paranoia
Poor concentration
Poor insight
Poor motivation
Suicidality
Tearfulness
Tobacco use
Weight loss
Value
Time (FUP)
FU1
FU2
FU3
FU4
FU5
FU6

(B)

(C) Linear regression results of node residual on time. Before scaling the data, a linear regression analysis was performed for each node, regressing the residuals on time to assess potential temporal trends. For all nodes, the regression results indicated no significant effect of time (time coefficient = 0, p-value=1). Results from the regression analysis are appended.

| **Prodromal feature** | **Time coefficient** | **SE time coefficient** | **P-value time** |
|---|---|---|---|
| Aggression | 0 | 0.001 | 1 |
| Agitation | 0 | 0.001 | 1 |
| Anxiety | 0 | 0.001 | 1 |
| Cannabis use | 0 | 0.001 | 1 |
| Cocaine use | 0 | 0.001 | 1 |
| Cognitive impairment | 0 | 0.001 | 1 |
| Delusional thinking | 0 | 0.001 | 1 |
| Disturbed sleep | 0 | 0.001 | 1 |
| Emotionally withdrawal | 0 | 0.001 | 1 |
| Feeling hopeless | 0 | 0.001 | 1 |
| Guilt | 0 | 0.001 | 1 |
| Hallucinations (any) | 0 | 0.001 | 1 |
| Hostility | 0 | 0.001 | 1 |
| Irritability | 0 | 0.001 | 1 |
| Mood instability | 0 | 0.001 | 1 |
| Paranoia | 0 | 0.001 | 1 |
| Poor concentration | 0 | 0.001 | 1 |
| Poor insight | 0 | 0.001 | 1 |
| Poor motivation | 0 | 0.001 | 1 |
| Suicidality | 0 | 0.001 | 1 |
| Tearfulness | 0 | 0.001 | 1 |
| Tobacco use | 0 | 0.001 | 1 |
| Weight loss | 0 | 0.001 | 1 |

eResults 2 SMD model fit

Simulated data (n=6,455) using the estimated model structure was generated and refitted to estimate recoverability, demonstrating good fit (RMSEA=.028 [95%CI:.028, .029]; $X^2(8625)=53531.88$, p<.002; CFI=.68; TLI=.65).

eTable 8 SMD temporal network edge estimates. (A) Actual model (green) and (B) bootstrapped estimates (blue, standard error of the mean). Any bootstrapped edges that had 95% CI crossing zero were forced to 0. Edge direction is defined from the node in the column to the node in the row.

(A)

| | AGGR | AGIT | ANX | CANN | COC | COG | DEL | SLEEP | EMOT | HOPE | GUILT | HALL | HOST | IRR | MOOD | PAR | CONC | INS | MOTIV | SUIC | TEAR | TOB | WGHT |
|---|---|---|---|---|---|---|---|---|---|---|---|---|---|---|---|---|---|---|---|---|---|---|---|
| ***AGGR*** | 0.06 | 0.04 | -0.02 | -0.02 | 0.01 | 0 | 0.01 | 0 | 0.01 | 0 | -0.01 | -0.01 | 0.04 | -0.01 | 0 | 0.02 | 0 | 0 | -0.01 | -0.01 | 0.01 | 0 | 0 |
| ***AGIT*** | 0.03 | 0.07 | -0.01 | -0.01 | -0.01 | 0 | 0.01 | -0.01 | 0 | 0 | 0 | 0.01 | 0 | 0.01 | 0 | 0 | 0 | 0 | -0.01 | 0 | 0.01 | 0.01 | 0 |
| ***ANX*** | 0.02 | 0 | 0.06 | -0.02 | -0.01 | 0.01 | 0.01 | 0.01 | 0.01 | 0 | 0 | 0 | 0 | -0.01 | 0.01 | 0.01 | 0 | 0 | 0 | 0 | 0.03 | 0 | -0.01 |
| ***CANN*** | 0.01 | 0.02 | -0.01 | 0.05 | 0.01 | 0 | 0 | 0 | 0 | 0 | -0.01 | 0 | 0.01 | 0 | 0.01 | 0.01 | -0.01 | -0.02 | 0 | 0 | 0 | 0 | 0 |
| ***COC*** | 0.02 | 0.01 | -0.01 | 0.01 | 0.09 | 0 | 0 | 0 | 0 | -0.01 | -0.01 | 0.01 | 0.01 | 0.01 | -0.01 | -0.01 | -0.01 | -0.02 | 0 | 0 | 0 | 0 | 0 |
| ***COG*** | 0.01 | 0.01 | 0.01 | -0.01 | 0.02 | 0.04 | 0.01 | 0.02 | 0.01 | 0.01 | 0.01 | 0 | 0 | 0 | 0.01 | 0.01 | 0.01 | 0 | 0 | 0 | 0.02 | -0.01 | 0.01 |
| ***DEL*** | 0.01 | 0.02 | -0.01 | -0.01 | 0 | 0.01 | 0.08 | -0.02 | 0.02 | -0.01 | -0.01 | 0.03 | 0.02 | -0.01 | -0.02 | 0.02 | 0 | 0 | 0 | 0 | 0 | 0 | 0 |
| ***SLEEP*** | 0.01 | 0.01 | -0.01 | 0.01 | -0.01 | -0.01 | -0.01 | 0.07 | 0.02 | 0.01 | 0.01 | 0 | 0 | -0.01 | 0 | 0 | 0.01 | 0.01 | 0 | -0.01 | 0.02 | 0 | -0.01 |
| ***EMOT*** | -0.01 | 0 | 0 | 0 | 0 | -0.01 | -0.01 | 0.01 | 0.05 | -0.01 | 0.01 | 0 | 0.02 | 0 | 0 | 0.02 | -0.01 | -0.01 | -0.01 | 0.02 | 0.02 | 0 | 0.01 |
| ***HOPE*** | 0 | 0.01 | 0 | -0.01 | 0 | -0.01 | 0 | 0.01 | 0.02 | 0.07 | 0 | -0.02 | -0.01 | -0.01 | 0 | 0 | 0.02 | 0.01 | 0 | 0.02 | 0.02 | 0 | 0.01 |
| ***GUILT*** | 0 | 0 | -0.01 | -0.01 | -0.01 | 0.01 | 0 | 0.01 | 0.01 | 0.01 | 0.1 | -0.01 | 0 | -0.01 | 0.01 | 0 | 0.01 | 0 | 0 | 0.01 | 0.03 | -0.02 | 0.01 |
| ***HALL*** | 0 | 0.01 | -0.01 | -0.02 | -0.01 | -0.01 | 0.02 | 0.01 | 0 | 0 | -0.01 | 0.09 | 0.01 | -0.01 | -0.01 | 0.03 | 0.01 | -0.01 | 0 | 0.01 | 0.01 | -0.01 | 0 |
| ***HOST*** | 0.02 | 0 | 0 | -0.01 | 0.01 | 0 | 0.02 | -0.02 | 0 | 0 | 0 | 0 | 0.08 | 0.02 | 0.01 | 0.01 | -0.01 | -0.01 | 0 | -0.01 | 0.01 | 0 | 0 |
| ***IRR*** | 0.02 | 0.02 | -0.01 | -0.01 | 0 | 0.02 | 0 | 0 | 0.01 | -0.01 | -0.01 | 0.01 | 0.02 | 0.07 | 0 | 0.01 | 0 | -0.01 | 0 | 0 | 0.01 | 0 | -0.01 |
| ***MOOD*** | 0.01 | 0.02 | -0.02 | -0.01 | -0.02 | 0.01 | 0 | 0 | 0 | 0.02 | 0 | 0 | 0.02 | 0 | 0.06 | 0 | 0.02 | -0.01 | -0.01 | 0 | 0.02 | 0 | 0 |
| ***PAR*** | 0.01 | 0.01 | -0.01 | -0.01 | -0.01 | 0.01 | 0.02 | 0.01 | 0.02 | -0.02 | -0.02 | 0.02 | 0.01 | 0 | 0 | 0.09 | 0.01 | 0 | -0.01 | -0.01 | 0.01 | -0.01 | 0 |
| ***CONC*** | 0.01 | 0.02 | -0.01 | 0 | -0.01 | -0.01 | -0.01 | 0.01 | -0.01 | 0.01 | -0.01 | 0 | 0 | 0 | 0.01 | 0.02 | 0.06 | 0 | 0 | 0 | 0.03 | 0 | 0 |
| ***INS*** | 0.01 | 0 | -0.01 | -0.02 | 0 | -0.01 | 0.01 | 0 | 0 | 0 | 0.01 | 0 | 0.02 | 0.01 | 0.01 | 0.01 | 0.01 | 0.07 | 0 | 0.02 | 0.03 | 0 | -0.01 |
| ***MOTIV*** | -0.01 | -0.01 | 0.01 | 0.01 | -0.01 | 0.01 | 0 | 0 | 0.01 | 0 | -0.01 | 0.01 | -0.01 | 0.01 | 0.01 | 0 | 0.01 | 0.02 | 0.06 | -0.01 | 0.01 | -0.01 | 0 |
| ***SUIC*** | 0 | 0.01 | 0 | -0.01 | -0.01 | -0.01 | 0 | -0.01 | 0.01 | 0.01 | 0 | 0.01 | 0 | 0 | -0.02 | -0.01 | 0 | 0 | 0.01 | 0.08 | 0.02 | 0 | 0.01 |
| ***TEAR*** | 0 | 0.01 | 0.01 | -0.01 | 0 | 0 | 0.01 | 0 | 0 | 0.01 | 0.01 | 0 | 0 | -0.01 | 0 | -0.01 | 0.02 | 0.02 | 0 | 0 | 0.12 | -0.02 | 0 |

| | | | | | | | | | | | | | | | | | | | | | | | |
|---|---|---|---|---|---|---|---|---|---|---|---|---|---|---|---|---|---|---|---|---|---|---|---|
| ***TOB*** | 0.02 | 0.01 | 0.01 | 0.01 | -0.01 | 0 | 0.01 | 0 | 0 | -0.01 | -0.01 | 0.01 | 0.01 | 0 | 0 | 0 | -0.01 | -0.01 | -0.01 | 0.02 | 0.01 | 0.08 | 0 |
| ***WGHT*** | -0.01 | 0 | -0.01 | 0 | -0.02 | -0.01 | 0 | 0 | 0.01 | -0.01 | 0 | 0 | 0 | -0.02 | 0.01 | 0 | 0.01 | -0.01 | 0.02 | 0.02 | 0.01 | -0.01 | 0.06 |

(B)

| | AGGR | AGIT | ANX | CANN | COC | COG | DEL | SLEEP | EMOT | HOPE | GUILT | HALL | HOST | IRR | MOOD | PAR | CONC | INS | MOTIV | SUIC | TEAR | TOB | WGHT |
|---|---|---|---|---|---|---|---|---|---|---|---|---|---|---|---|---|---|---|---|---|---|---|---|
| ***AGGR*** | 0.043 (0.0035) | 0.017 (0.004) | -0.028 (0.0029) | -0.034 (0.0024) | 0 (0) | -0.014 (0.0037) | 0 (0) | -0.019 (0.0031) | 0 (0) | -0.01 (0.0016) | -0.014 (0.0016) | -0.027 (0.0032) | 0.03 (0.0026) | -0.02 (0.0031) | -0.014 (0.0025) | 0 (0) | -0.016 (0.0026) | -0.015 (0.0027) | -0.01 (0.0015) | -0.018 (0.0019) | -0.005 (0.0022) | -0.012 (0.0026) | 0 (0) |
| ***AGIT*** | 0.015 (0.0039) | 0.053 (0.0045) | -0.021 (0.0031) | -0.024 (0.0027) | -0.016 (0.0021) | -0.016 (0.0039) | -0.008 (0.0038) | -0.022 (0.0034) | -0.005 (0.0016) | 0 (0) | -0.005 (0.0018) | 0 (0) | -0.009 (0.0026) | -0.009 (0.0034) | -0.011 (0.0027) | -0.018 (0.004) | -0.014 (0.003) | -0.014 (0.003) | -0.016 (0.0017) | -0.008 (0.0022) | 0 (0) | -0.006 (0.0027) | 0 (0) |
| ***ANX*** | 0 (0) | -0.012 (0.0038) | 0.048 (0.0029) | -0.031 (0.003) | -0.018 (0.0028) | -0.008 (0.0038) | 0 (0) | 0 (0) | 0 (0) | -0.008 (0.002) | 0 (0) | -0.011 (0.0035) | -0.014 (0.002) | -0.019 (0.0026) | 0 (0) | 0 (0) | -0.008 (0.0021) | -0.006 (0.0022) | 0 (0) | -0.009 (0.0015) | 0.015 (0.0022) | -0.009 (0.0022) | -0.007 (0.0012) |
| ***CANN*** | 0 (0) | 0 (0) | -0.025 (0.0027) | 0.046 (0.002) | 0 (0) | -0.019 (0.0033) | -0.01 (0.0027) | -0.01 (0.0029) | 0 (0) | -0.013 (0.002) | -0.02 (0.0019) | -0.014 (0.0027) | 0 (0) | -0.015 (0.0027) | 0 (0) | 0 (0) | -0.023 (0.0028) | -0.029 (0.0028) | -0.007 (0.002) | -0.013 (0.002) | -0.011 (0.0022) | -0.009 (0.0026) | -0.008 (0.0013) |
| ***COC*** | 0 (0) | 0 (0) | -0.02 (0.002) | 0 (0) | 0.083 (0.0016) | -0.012 (0.0025) | -0.01 (0.002) | -0.009 (0.0022) | -0.004 (0.0013) | -0.013 (0.0016) | -0.012 (0.0016) | 0 (0) | 0.004 (0.0015) | 0 (0) | -0.019 (0.0019) | -0.017 (0.0021) | -0.015 (0.0022) | -0.025 (0.0021) | -0.01 (0.0017) | -0.007 (0.0016) | -0.008 (0.0017) | -0.005 (0.0016) | -0.008 (0.0011) |
| ***COG*** | -0.015 (0.0046) | 0 (0) | -0.007 (0.0034) | -0.023 (0.003) | 0.006 (0.0027) | 0.023 (0.0042) | 0 (0) | 0 (0) | 0 (0) | 0 (0) | 0.007 (0.0021) | -0.017 (0.0038) | -0.01 (0.0028) | -0.019 (0.0033) | 0 (0) | -0.011 (0.0042) | 0 (0) | -0.015 (0.0028) | -0.005 (0.0024) | -0.006 (0.0019) | 0.008 (0.0026) | -0.026 (0.0028) | 0 (0) |
| ***DEL*** | 0 (0) | 0 (0) | -0.025 (0.0028) | -0.02 (0.0023) | -0.012 (0.0017) | 0 (0) | 0.063 (0.0034) | -0.037 (0.003) | 0.013 (0.0016) | -0.015 (0.002) | -0.018 (0.0018) | 0.009 (0.0034) | 0.01 (0.0026) | -0.02 (0.003) | -0.029 (0.0025) | 0 (0) | -0.012 (0.0028) | -0.016 (0.0029) | -0.005 (0.0017) | -0.014 (0.0021) | -0.008 (0.0024) | -0.009 (0.0025) | -0.004 (0.0014) |
| ***SLEEP*** | 0 (0.0037) | -0.01 (0.0041) | -0.017 (0.0029) | 0 (0.0032) | -0.018 (0.0027) | -0.027 (0.0037) | -0.025 (0.0037) | 0 (0) | 0.019 (0.0019) | 0 (0.0019) | 0.006 (0.0023) | -0.015 (0.0037) | -0.01 (0.0021) | -0.024 (0.0029) | -0.005 (0.0021) | -0.011 (0.0039) | 0 (0.0025) | 0 (0.0025) | 0 (0.003) | -0.015 (0.0016) | 0.011 (0.0024) | -0.01 (0.0024) | -0.013 (0.0013) |
| ***EMOT*** | -0.017 (0.0026) | -0.013 (0.0026) | -0.004 (0.0017) | -0.008 (0.0025) | -0.006 (0.0025) | -0.017 (0.0023) | -0.022 (0.0028) | 0 (0.0019) | 0.043 (0.002) | -0.008 (0.0021) | 0.015 (0.0025) | -0.015 (0.0025) | 0.012 (0.0014) | -0.009 (0.0018) | 0 (0) | 0.006 (0.0028) | 0 (0) | 0 (0) | -0.009 (0.0034) | 0.013 (0.0011) | 0.009 (0.0013) | -0.01 (0.0015) | 0.005 (0.0012) |
| ***HOPE*** | -0.009 (0.0027) | 0 (0.0028) | -0.01 (0.0017) | -0.022 (0.0027) | 0 (0) | -0.022 (0.0024) | -0.013 (0.003) | 0 (0.002) | 0.021 (0.0022) | 0.07 (0.0024) | 0 (0.0028) | -0.029 (0.0028) | -0.015 (0.0015) | -0.019 (0.0018) | 0 (0.0017) | 0 (0) | 0.013 (0.0017) | 0 (0.0017) | 0 (0) | 0.017 (0.0013) | 0.014 (0.0014) | -0.005 (0.0015) | 0.006 (0.0013) |
| ***GUILT*** | -0.013 (0.0026) | -0.01 (0.0027) | -0.019 (0.0016) | -0.016 (0.0028) | -0.017 (0.0027) | -0.005 (0.0024) | 0 (0) | 0 (0) | 0.007 (0.0022) | 0.006 (0.0023) | 0.097 (0.003) | -0.018 (0.0026) | 0 (0) | -0.018 (0.0017) | 0 (0.0016) | -0.009 (0.0027) | 0 (0.0014) | 0 (0.0014) | 0.008 (0.0039) | 0.003 (0.001) | 0 (0) | -0.023 (0.0014) | 0.012 (0.0013) |
| ***HALL*** | -0.011 (0.0032) | 0 (0) | -0.02 (0.0029) | -0.028 (0.0023) | -0.015 (0.0018) | -0.022 (0.0036) | 0 (0.0035) | 0 (0) | -0.01 (0.0017) | -0.012 (0.0018) | 0 (0) | 0.074 (0.0036) | 0 (0.0025) | -0.024 (0.0031) | 0 (0) | 0.012 (0.0038) | -0.006 (0.0029) | 0 (0) | 0 (0) | -0.005 (0.0023) | 0 (0.0024) | -0.019 (0.0026) | -0.009 (0.0014) |
| ***HOST*** | 0 (0) | -0.01 (0.0031) | -0.007 (0.0018) | -0.018 (0.0023) | 0 (0.0023) | -0.011 (0.0027) | 0 (0) | -0.03 (0.0021) | 0 (0) | 0 (0) | -0.005 (0.0019) | -0.014 (0.0027) | 0.073 (0.0021) | 0 (0) | 0 (0) | 0 (0) | 0 (0) | -0.017 (0.0017) | 0 (0) | -0.012 (0.001) | 0.003 (0.0015) | -0.007 (0.0017) | -0.005 (0.001) |
| ***IRR*** | 0 (0) | 0 (0) | -0.024 (0.0025) | -0.022 (0.0031) | -0.007 (0.0029) | 0 (0) | -0.011 (0.0037) | -0.012 (0.0028) | 0 (0) | -0.016 (0.0021) | 0 (0) | -0.009 (0.0035) | 0.005 (0.0024) | 0.056 (0.0032) | -0.009 (0.0021) | -0.011 (0.0039) | -0.015 (0.0023) | -0.021 (0.0023) | 0 (0) | -0.003 (0.0015) | -0.004 (0.002) | -0.009 (0.0022) | 0 (0) |
| ***MOOD*** | 0 (0.0034) | 0 (0.0036) | 0 (0) | 0 (0) | -0.024 (0.0032) | 0 (0) | -0.013 (0.0037) | -0.012 (0.0024) | 0 (0.0025) | 0 (0) | 0 (0.0033) | 0 (0) | 0.009 (0.0018) | -0.007 (0.0024) | 0 (0) | -0.011 (0.0036) | 0.008 (0.002) | -0.016 (0.0019) | 0 (0.0044) | -0.003 (0.0013) | 0.009 (0.0017) | -0.01 (0.0019) | 0 (0) |

| | | | | | | | | | | | | | | | | | | | | | | | |
|---|---|---|---|---|---|---|---|---|---|---|---|---|---|---|---|---|---|---|---|---|---|---|---|
| ***PAR*** | -0.009 (0.0037) | -0.008 (0.004) | -0.023 (0.0034) | -0.02 (0.0024) | -0.019 (0.0017) | -0.013 (0.004) | 0 (0) | -0.012 (0.0036) | 0.005 (0.0023) | -0.028 (0.0026) | -0.029 (0.0026) | 0 (0) | 0 (0) | -0.013 (0.0035) | -0.013 (0.0032) | 0.073 (0.0041) | -0.01 (0.0035) | -0.021 (0.0036) | -0.017 (0.0023) | -0.021 (0.0026) | -0.007 (0.003) | -0.019 (0.0029) | -0.006 (0.0018) |
| ***CONC*** | 0 (0) | 0 (0.004) | -0.024 (0.0024) | 0 (0) | -0.02 (0.0034) | -0.023 (0.0037) | -0.028 (0.0039) | -0.006 (0.0029) | -0.011 (0.0026) | 0 (0) | -0.007 (0.0033) | -0.014 (0.0038) | -0.007 (0.0019) | -0.016 (0.0026) | 0 (0.0021) | 0 (0.0041) | 0 (0) | 0 (0) | 0 (0) | -0.005 (0.0015) | 0.021 (0.0021) | -0.014 (0.0021) | 0 (0.0016) |
| ***INS*** | 0 (0) | 0 (0) | -0.023 (0.0023) | -0.035 (0.0035) | -0.015 (0.0034) | 0 (0) | 0 (0.0041) | -0.017 (0.0027) | 0 (0) | 0 (0.0026) | 0.007 (0.0033) | -0.011 (0.0038) | 0.01 (0.0019) | 0 (0.0024) | 0 (0.002) | 0 (0.004) | 0 (0.0021) | 0.063 (0.0025) | 0 (0) | 0.017 (0.0014) | 0.016 (0.0019) | -0.008 (0.0021) | -0.01 (0.0016) |
| ***MOTIV*** | -0.017 (0.0027) | 0 (0) | 0 (0.0014) | 0 (0.0028) | -0.018 (0.0028) | 0 (0.0023) | -0.008 (0.0028) | -0.009 (0.0017) | 0 (0) | 0 (0) | -0.007 (0.0032) | 0 (0) | -0.013 (0.0013) | 0.004 (0.0016) | 0 (0.0016) | -0.015 (0.0026) | 0.004 (0.0015) | 0.011 (0.0014) | 0.061 (0.0042) | -0.014 (0.0012) | 0.003 (0.0011) | -0.013 (0.0012) | 0 (0.0014) |
| ***SUIC*** | -0.015 (0.0028) | 0 (0) | -0.009 (0.002) | -0.016 (0.0025) | -0.014 (0.0023) | -0.026 (0.0027) | 0 (0) | -0.023 (0.0022) | 0 (0.0017) | 0 (0) | 0 (0) | 0 (0.003) | -0.013 (0.0017) | -0.006 (0.0022) | -0.022 (0.0016) | -0.026 (0.0029) | -0.01 (0.0019) | -0.006 (0.002) | 0.011 (0.0026) | 0.074 (0.0017) | 0.008 (0.0018) | -0.006 (0.0019) | 0 (0) |
| ***TEAR*** | 0 (0) | 0 (0) | 0 (0) | 0 (0) | -0.008 (0.002) | -0.018 (0.003) | 0 (0) | -0.016 (0.0025) | 0 (0) | 0 (0.0015) | 0 (0.0018) | -0.014 (0.0027) | 0 (0) | -0.017 (0.0024) | -0.005 (0.0018) | -0.018 (0.0029) | 0.006 (0.0022) | 0.005 (0.0021) | -0.006 (0.0021) | -0.008 (0.0015) | 0 (0) | -0.033 (0.0019) | 0 (0.001) |
| ***TOB*** | 0 (0) | 0 (0) | 0 (0.0023) | 0 (0.0029) | -0.017 (0.0024) | -0.018 (0.0031) | 0 (0.0033) | -0.014 (0.0026) | 0 (0.0018) | 0 (0) | -0.015 (0.0021) | -0.007 (0.0031) | 0 (0.0019) | -0.016 (0.0024) | 0 (0) | 0 (0) | -0.019 (0.0021) | -0.023 (0.0021) | 0 (0) | 0.009 (0.0014) | 0 (0.0017) | 0.07 (0.0024) | 0 (0) |
| ***WGHT*** | -0.017 (0.0021) | -0.006 (0.0023) | -0.021 (0.0015) | -0.006 (0.002) | -0.025 (0.0018) | -0.019 (0.0021) | -0.007 (0.0023) | -0.004 (0.0017) | 0 (0) | -0.013 (0.0014) | 0 (0) | -0.01 (0.0021) | -0.009 (0.0012) | -0.022 (0.0016) | 0.003 (0.0012) | -0.008 (0.0022) | 0 (0) | -0.017 (0.0014) | 0.016 (0.0023) | 0.014 (9e-04) | 0 (0) | -0.019 (0.0013) | 0.056 (0.0011) |

eTable 9 SMD contemporaneous and between-subject network edge estimates. (A) Contemporaneous network edge estimates and (B) between-subject network edge estimates (actual model in green and bootstrapped estimates in blue). Any bootstrapped edges that had 95% CI crossing zero were forced to 0.

(A)

| | AGGR | AGIT | ANX | CANN | COC | COG | DEL | SLEEP | EMOT | HOPE | GUILT | HALL | HOST | IRR | MOOD | PAR | CONC | INS | MOTIV | SUIC | TEAR | TOB | WGHT |
|---|---|---|---|---|---|---|---|---|---|---|---|---|---|---|---|---|---|---|---|---|---|---|---|
| ***AGGR*** | NA | 0.15 (0.0092) | 0.069 (0.0077) | 0.057 (0.0073) | 0.067 (0.0051) | 0.103 (0.0091) | 0.103 (0.0084) | 0.063 (0.0089) | 0.065 (0.0066) | 0.031 (0.0056) | 0.034 (0.0056) | 0.079 (0.0084) | 0.168 (0.0071) | 0.143 (0.0087) | 0.082 (0.0075) | 0.11 (0.0092) | 0.039 (0.0079) | 0.073 (0.0079) | 0.032 (0.005) | 0.058 (0.0063) | 0.028 (0.0068) | 0.096 (0.0074) | 0.034 (0.0047) |
| ***AGIT*** | 0.1 | NA | 0.104 (0.0084) | 0.055 (0.0074) | 0.058 (0.0055) | 0.078 (0.0105) | 0.111 (0.0089) | 0.103 (0.0092) | 0.047 (0.0061) | 0.045 (0.0065) | 0.038 (0.006) | 0.092 (0.0092) | 0.115 (0.0074) | 0.138 (0.0091) | 0.076 (0.0081) | 0.117 (0.0098) | 0.076 (0.0086) | 0.075 (0.0086) | 0.025 (0.0055) | 0.076 (0.0069) | 0.089 (0.0074) | 0.096 (0.0077) | 0.04 (0.0051) |
| ***ANX*** | 0.03 | 0.06 | NA | 0.052 (0.0063) | 0.028 (0.0047) | 0.219 (0.0086) | 0.048 (0.0075) | 0.133 (0.0082) | 0.04 (0.0048) | 0.039 (0.0053) | 0.047 (0.005) | 0.06 (0.0077) | 0.025 (0.0058) | 0.052 (0.0075) | 0.051 (0.0067) | 0.109 (0.0088) | 0.054 (0.0078) | 0.071 (0.0072) | 0.035 (0.0044) | 0.043 (0.0059) | 0.105 (0.0066) | 0.055 (0.0064) | 0.045 (0.0043) |
| ***CANN*** | 0.02 | 0.02 | 0.02 | NA | 0.264 (0.0047) | 0.066 (0.0073) | 0.082 (0.0066) | 0.045 (0.0067) | 0.024 (0.0047) | 0.042 (0.005) | 0.035 (0.0049) | 0.07 (0.007) | 0.052 (0.0053) | 0.061 (0.0067) | 0.065 (0.0061) | 0.12 (0.008) | 0.036 (0.0065) | 0.077 (0.0067) | 0.04 (0.0045) | 0.04 (0.0054) | 0.018 (0.0055) | 0.155 (0.006) | 0.042 (0.0042) |
| ***COC*** | 0.04 | 0.03 | 0 | 0.25 | NA | 0.043 (0.0053) | 0.057 (0.0049) | 0.028 (0.0051) | 0.029 (0.0036) | 0.017 (0.004) | 0.032 (0.0038) | 0.07 (0.0049) | 0.028 (0.0041) | 0.05 (0.005) | 0.034 (0.0049) | 0.042 (0.0057) | 0.028 (0.005) | 0.039 (0.0051) | 0.022 (0.0037) | 0.034 (0.004) | 0.038 (0.004) | 0.042 (0.0047) | 0.033 (0.0032) |
| ***COG*** | 0.06 | 0.03 | 0.18 | 0.03 | 0.02 | NA | 0.079 (0.0088) | 0.122 (0.0097) | 0.045 (0.0059) | 0.057 (0.0065) | 0.061 (0.0061) | 0.085 (0.0092) | 0.046 (0.0069) | 0.082 (0.0088) | 0.066 (0.008) | 0.115 (0.0098) | 0.192 (0.0087) | 0.063 (0.0087) | 0.074 (0.006) | 0.047 (0.0071) | 0.097 (0.0078) | 0.067 (0.0076) | 0.042 (0.0052) |
| ***DEL*** | 0.06 | 0.07 | 0.01 | 0.05 | 0.03 | 0.03 | NA | 0.037 (0.008) | 0.035 (0.0055) | 0.029 (0.0056) | 0.024 (0.0053) | 0.212 (0.0078) | 0.103 (0.0066) | 0.077 (0.008) | 0.049 (0.0072) | 0.188 (0.0086) | 0.047 (0.0075) | 0.099 (0.0079) | 0.03 (0.005) | 0.049 (0.0061) | 0.021 (0.0063) | 0.067 (0.0069) | 0.045 (0.0046) |
| ***SLEEP*** | 0.02 | 0.06 | 0.09 | 0.01 | 0 | 0.07 | 0 | NA | 0.053 (0.0052) | 0.059 (0.0058) | 0.063 (0.0055) | 0.086 (0.008) | 0.035 (0.0062) | 0.093 (0.0081) | 0.076 (0.0073) | 0.086 (0.009) | 0.127 (0.0083) | 0.095 (0.0078) | 0.06 (0.0047) | 0.073 (0.0066) | 0.106 (0.0073) | 0.077 (0.0069) | 0.053 (0.0047) |
| ***EMOT*** | 0.03 | 0.02 | 0.02 | 0 | 0.01 | 0.01 | 0.01 | 0.03 | NA | 0.039 (0.0033) | 0.038 (0.003) | 0.055 (0.0055) | 0.05 (0.0041) | 0.063 (0.0052) | 0.041 (0.0043) | 0.066 (0.0061) | 0.047 (0.0049) | 0.041 (0.0047) | 0.047 (0.0028) | 0.055 (0.0039) | 0.041 (0.0043) | 0.036 (0.0044) | 0.049 (0.0027) |
| ***HOPE*** | 0 | 0.01 | 0.01 | 0.02 | 0 | 0.03 | 0 | 0.03 | 0.02 | NA | 0.117 (0.004) | 0.042 (0.0058) | 0.008 (0.0039) | 0.022 (0.0052) | 0.071 (0.0047) | 0.027 (0.0065) | 0.093 (0.0054) | 0.066 (0.0051) | 0 (0) | 0.129 (0.0047) | 0.078 (0.0051) | 0.028 (0.0046) | 0.048 (0.0031) |
| ***GUILT*** | 0.01 | 0.01 | 0.02 | 0.01 | 0.01 | 0.03 | 0 | 0.03 | 0.02 | 0.1 | NA | 0.019 (0.0055) | 0.011 (0.0037) | 0.04 (0.005) | 0.056 (0.0043) | 0.03 (0.0062) | 0.067 (0.0051) | 0.058 (0.0046) | 0.059 (0.0028) | 0.055 (0.0041) | 0.101 (0.0048) | 0.037 (0.0043) | 0.04 (0.0027) |
| ***HALL*** | 0.04 | 0.05 | 0.02 | 0.04 | 0.05 | 0.04 | 0.17 | 0.05 | 0.03 | 0.01 | -0.01 | NA | 0.066 (0.0065) | 0.053 (0.008) | 0.048 (0.0072) | 0.174 (0.0087) | 0.065 (0.0078) | 0.099 (0.008) | 0.048 (0.0051) | 0.08 (0.0063) | 0.034 (0.0065) | 0.064 (0.007) | 0.03 (0.0047) |
| ***HOST*** | 0.13 | 0.08 | 0 | 0.03 | 0.01 | 0.01 | 0.07 | 0 | 0.03 | -0.01 | -0.01 | 0.03 | NA | 0.151 (0.0068) | 0.062 (0.0054) | 0.068 (0.0073) | 0.021 (0.0057) | 0.035 (0.0058) | 0 (0.0033) | 0.018 (0.0047) | 0.042 (0.0049) | 0.063 (0.0056) | 0.017 (0.0034) |
| ***IRR*** | 0.1 | 0.09 | 0.01 | 0.03 | 0.02 | 0.04 | 0.04 | 0.05 | 0.04 | 0 | 0.01 | 0.01 | 0.12 | NA | 0.102 (0.0071) | 0.087 (0.0088) | 0.071 (0.0074) | 0.056 (0.0074) | 0.021 (0.0045) | 0.031 (0.0059) | 0.066 (0.0065) | 0.075 (0.0069) | 0.037 (0.0044) |

| | | | | | | | | | | | | | | | | | | | | | | | |
|---|---|---|---|---|---|---|---|---|---|---|---|---|---|---|---|---|---|---|---|---|---|---|---|
| ***MOOD*** | 0.05 | 0.04 | 0.02 | 0.03 | 0.01 | 0.03 | 0.01 | 0.04 | 0.02 | 0.05 | 0.04 | 0.01 | 0.04 | 0.07 | NA | 0.069 (0.008) | 0.086 (0.0067) | 0.065 (0.0065) | 0.042 (0.0037) | 0.053 (0.0054) | 0.096 (0.006) | 0.059 (0.006) | 0.043 (0.0038) |
| ***PAR*** | 0.06 | 0.07 | 0.06 | 0.08 | 0.01 | 0.07 | 0.15 | 0.04 | 0.04 | 0 | 0 | 0.13 | 0.03 | 0.04 | 0.03 | NA | 0.059 (0.0086) | 0.11 (0.0088) | 0.027 (0.0058) | 0.045 (0.007) | 0.05 (0.0073) | 0.071 (0.0079) | 0.042 (0.0053) |
| ***CONC*** | 0 | 0.03 | 0.02 | 0 | 0 | 0.15 | 0.01 | 0.08 | 0.02 | 0.07 | 0.04 | 0.03 | -0.01 | 0.03 | 0.05 | 0.02 | NA | 0.092 (0.0073) | 0.071 (0.0044) | 0.073 (0.0061) | 0.095 (0.0068) | 0.046 (0.0064) | 0.059 (0.0043) |
| ***INS*** | 0.03 | 0.03 | 0.03 | 0.04 | 0.01 | 0.02 | 0.06 | 0.06 | 0.02 | 0.04 | 0.04 | 0.06 | 0 | 0.02 | 0.03 | 0.07 | 0.06 | NA | 0.042 (0.004) | 0.07 (0.0058) | 0.071 (0.0063) | 0.061 (0.0065) | 0.029 (0.0042) |
| ***MOTIV*** | 0.01 | 0 | 0.01 | 0.02 | 0 | 0.05 | 0.01 | 0.04 | 0.03 | 0.06 | 0.05 | 0.02 | -0.02 | 0 | 0.02 | 0 | 0.05 | 0.02 | NA | 0.059 (0.0035) | 0.032 (0.004) | 0.019 (0.0039) | 0.036 (0.0024) |
| ***SUIC*** | 0.02 | 0.04 | 0.01 | 0.01 | 0.01 | 0.01 | 0.02 | 0.04 | 0.04 | 0.11 | 0.03 | 0.05 | -0.01 | 0 | 0.03 | 0.01 | 0.04 | 0.04 | 0.04 | NA | 0.101 (0.0055) | 0.049 (0.0051) | 0.037 (0.0035) |
| ***TEAR*** | -0.01 | 0.05 | 0.07 | -0.01 | 0.02 | 0.06 | -0.01 | 0.07 | 0.02 | 0.05 | 0.08 | 0 | 0.02 | 0.03 | 0.07 | 0.01 | 0.06 | 0.04 | 0.01 | 0.07 | NA | 0.043 (0.0056) | 0.043 (0.0038) |
| ***TOB*** | 0.06 | 0.06 | 0.02 | 0.13 | 0.02 | 0.03 | 0.03 | 0.04 | 0.01 | 0.01 | 0.02 | 0.03 | 0.04 | 0.04 | 0.03 | 0.03 | 0.02 | 0.03 | 0 | 0.02 | 0.02 | NA | 0.04 (0.0039) |
| ***WGHT*** | 0.01 | 0.01 | 0.02 | 0.02 | 0.02 | 0.02 | 0.02 | 0.03 | 0.04 | 0.03 | 0.03 | 0.01 | 0 | 0.02 | 0.02 | 0.01 | 0.04 | 0.01 | 0.02 | 0.02 | 0.02 | 0.02 | NA |

(B)

| | AGGR | AGIT | ANX | CANN | COC | COG | DEL | SLEEP | EMOT | HOPE | GUILT | HALL | HOST | IRR | MOOD | PAR | CONC | INS | MOTIV | SUIC | TEAR | TOB | WGHT |
|---|---|---|---|---|---|---|---|---|---|---|---|---|---|---|---|---|---|---|---|---|---|---|---|
| ***AGGR*** | NA | 0.228 (0.005) | -0.05 (0.0035) | 0.12 (0.0027) | 0 (0) | 0.196 (0.005) | -0.027 (0.0024) | -0.117 (0.0049) | 0.06 (0.0049) | -0.042 (0.0043) | 0.012 (0.0038) | 0.047 (0.003) | 0.198 (0.0058) | 0.234 (0.0055) | 0.043 (0.0039) | 0.018 (0.0018) | -0.086 (0.0044) | 0.032 (0.0039) | -0.07 (0.0042) | 0.092 (0.0031) | -0.007 (0.0025) | 0.067 (0.0024) | -0.077 (0.0036) |
| ***AGIT*** | 0.25 | NA | 0.114 (0.0036) | 0.014 (0.0024) | 0.09 (0.0028) | 0 (0) | 0.099 (0.0026) | 0.107 (0.0041) | 0.142 (0.005) | -0.037 (0.0045) | 0.069 (0.0035) | 0.097 (0.0032) | 0.124 (0.0041) | 0.156 (0.0053) | 0.011 (0.0041) | 0.058 (0.0019) | 0.051 (0.0038) | -0.106 (0.0046) | 0.016 (0.0041) | 0.018 (0.0032) | 0.019 (0.0025) | -0.115 (0.0037) | -0.021 (0.0038) |
| ***ANX*** | -0.06 | 0.12 | NA | 0 (0) | -0.006 (0.0025) | 0.389 (0.0074) | -0.032 (0.0023) | 0.139 (0.0053) | -0.047 (0.0046) | 0.008 (0.0042) | 0.115 (0.0041) | -0.075 (0.0036) | -0.081 (0.0039) | -0.02 (0.0045) | 0.063 (0.0038) | 0.116 (0.0023) | 0.047 (0.0029) | 0.059 (0.0039) | -0.016 (0.0041) | -0.022 (0.003) | 0.034 (0.0022) | -0.032 (0.0023) | 0.06 (0.0036) |
| ***CANN*** | 0.12 | 0.01 | -0.01 | NA | 0.366 (0.0071) | 0.012 (0.0027) | 0.017 (0.0017) | -0.073 (0.0031) | -0.071 (0.0036) | -0.015 (0.0031) | -0.021 (0.0022) | -0.016 (0.0025) | 0.009 (0.0024) | 0.013 (0.0032) | -0.044 (0.0029) | 0.111 (0.0021) | -0.009 (0.002) | 0.178 (0.004) | 0.083 (0.0031) | -0.054 (0.0027) | 0.065 (0.0022) | 0.295 (0.0054) | 0.012 (0.0029) |
| ***COC*** | 0 | 0.1 | 0 | 0.4 | NA | -0.048 (0.0033) | -0.021 (0.0017) | -0.024 (0.0029) | -0.081 (0.0036) | 0.028 (0.0031) | 0.032 (0.0022) | 0.039 (0.0027) | 0.011 (0.0024) | -0.069 (0.0035) | 0.112 (0.0032) | 0.035 (0.0014) | -0.033 (0.002) | -0.063 (0.0036) | -0.018 (0.0033) | 0.032 (0.0027) | -0.043 (0.0022) | 0.018 (0.0014) | 0.065 (0.0032) |
| ***COG*** | 0.21 | 0.01 | 0.42 | 0.02 | -0.06 | NA | 0.047 (0.0023) | -0.024 (0.0052) | 0.136 (0.0054) | 0.085 (0.0048) | -0.021 (0.0041) | 0.08 (0.0035) | 0.03 (0.0041) | 0.033 (0.0051) | -0.028 (0.0043) | 0 (0) | 0.167 (0.0046) | -0.009 (0.0047) | 0.023 (0.0048) | 0.064 (0.0033) | 0.057 (0.0027) | 0.027 (0.0022) | 0.063 (0.0045) |
| ***DEL*** | -0.03 | 0.11 | -0.04 | 0.01 | -0.03 | 0.05 | NA | -0.013 (0.0025) | 0.01 (0.0031) | 0.09 (0.0027) | -0.064 (0.0026) | 0.141 (0.0027) | 0.107 (0.003) | 0.037 (0.0026) | -0.032 (0.0024) | 0.358 (0.0064) | 0.024 (0.0022) | 0.09 (0.0028) | -0.086 (0.0031) | -0.039 (0.0024) | -0.089 (0.0026) | 0.026 (0.0017) | -0.027 (0.0025) |
| ***SLEEP*** | -0.13 | 0.11 | 0.16 | -0.08 | -0.03 | -0.04 | -0.02 | NA | -0.113 (0.0058) | 0.136 (0.0056) | -0.088 (0.0046) | 0 (0) | -0.066 (0.005) | 0.217 (0.0067) | 0 (0.0045) | 0.037 (0.0019) | 0.202 (0.0049) | 0 (0) | 0.085 (0.0046) | 0.089 (0.0038) | 0.134 (0.0033) | 0.049 (0.0025) | 0.043 (0.0041) |
| ***EMOT*** | 0.08 | 0.16 | -0.05 | -0.09 | -0.09 | 0.14 | 0.01 | -0.11 | NA | -0.026 (0.0061) | -0.061 (0.0052) | 0 (0.0045) | -0.096 (0.0057) | -0.055 (0.0066) | 0 (0) | 0.01 (0.0025) | 0.132 (0.0049) | 0 (0.0056) | 0.183 (0.0062) | -0.085 (0.0045) | -0.028 (0.0035) | 0.064 (0.0034) | 0.117 (0.0053) |
| ***HOPE*** | -0.05 | -0.04 | 0 | -0.01 | 0.03 | 0.1 | 0.1 | 0.16 | -0.04 | NA | 0.3 (0.007) | 0.07 (0.0041) | 0.035 (0.0045) | -0.066 (0.0054) | 0 (0.0048) | -0.041 (0.0023) | -0.164 (0.0055) | 0.062 (0.0049) | 0.262 (0.0066) | 0.317 (0.0073) | 0.115 (0.0035) | -0.078 (0.0031) | 0.07 (0.0042) |
| ***GUILT*** | 0.01 | 0.07 | 0.13 | -0.02 | 0.03 | -0.04 | -0.08 | -0.09 | -0.05 | 0.33 | NA | -0.127 (0.0039) | -0.046 (0.0045) | -0.023 (0.0049) | 0 (0.004) | 0.02 (0.0019) | 0.217 (0.0053) | 0.05 (0.0038) | -0.073 (0.0044) | 0.018 (0.0034) | 0.114 (0.0031) | 0.121 (0.0032) | 0 (0) |
| ***HALL*** | 0.05 | 0.1 | -0.08 | -0.02 | 0.04 | 0.08 | 0.15 | 0.11 | 0 | 0.07 | -0.14 | NA | -0.101 (0.0039) | -0.092 (0.0047) | 0 (0) | 0.122 (0.0023) | 0.081 (0.0029) | 0.143 (0.004) | -0.08 (0.0045) | -0.034 (0.0029) | -0.01 (0.0022) | 0.106 (0.0025) | 0 (0.0036) |

| | | | | | | | | | | | | | | | | | | | | | | | |
|---|---|---|---|---|---|---|---|---|---|---|---|---|---|---|---|---|---|---|---|---|---|---|---|
| ***HOST*** | 0.23 | 0.13 | -0.1 | 0.01 | 0.01 | 0.02 | 0.12 | -0.07 | -0.1 | 0.04 | -0.04 | -0.11 | NA | 0.316 (0.0075) | -0.037 (0.0047) | 0.1 (0.0025) | -0.041 (0.0045) | -0.013 (0.0045) | -0.029 (0.0043) | -0.057 (0.0031) | -0.038 (0.0026) | 0.017 (0.0028) | 0.092 (0.004) |
| ***IRR*** | 0.25 | 0.17 | -0.01 | 0.02 | -0.07 | 0.03 | 0.04 | 0.24 | -0.06 | -0.08 | -0.03 | -0.1 | 0.34 | NA | 0.267 (0.0062) | -0.023 (0.0024) | 0.035 (0.0045) | -0.02 (0.0054) | 0 (0.0052) | -0.119 (0.0044) | 0.032 (0.003) | 0.094 (0.0033) | 0.089 (0.0048) |
| ***MOOD*** | 0.05 | 0.02 | 0.07 | -0.05 | 0.12 | -0.03 | -0.04 | 0 | 0.12 | 0.01 | -0.01 | -0.04 | -0.04 | 0.29 | NA | -0.008 (0.0022) | -0.043 (0.0042) | 0.22 (0.0061) | 0.133 (0.0052) | 0.148 (0.0042) | 0.207 (0.0048) | -0.02 (0.0026) | -0.092 (0.0043) |
| ***PAR*** | 0.01 | 0.05 | 0.13 | 0.12 | 0.04 | 0 | 0.39 | 0.04 | 0 | -0.05 | 0.02 | 0.13 | 0.11 | -0.03 | -0.01 | NA | -0.051 (0.0025) | 0.087 (0.0023) | 0.063 (0.0022) | 0.036 (0.0019) | -0.043 (0.0019) | 0.033 (0.0014) | -0.027 (0.0021) |
| ***CONC*** | -0.1 | 0.06 | 0.04 | -0.01 | -0.04 | 0.2 | 0.03 | 0.22 | 0.12 | -0.17 | 0.23 | 0.09 | -0.05 | 0.03 | -0.03 | -0.06 | NA | 0.117 (0.0042) | 0.177 (0.0051) | 0.122 (0.0035) | 0.03 (0.0025) | -0.072 (0.0031) | 0.078 (0.0029) |
| ***INS*** | 0.04 | -0.11 | 0.05 | 0.19 | -0.07 | -0.01 | 0.1 | 0.12 | 0.01 | 0.06 | 0.05 | 0.15 | -0.02 | -0.03 | 0.24 | 0.09 | 0.13 | NA | 0 (0) | 0.037 (0.0036) | -0.126 (0.0041) | -0.028 (0.0026) | 0.041 (0.0047) |
| ***MOTIV*** | -0.08 | 0.02 | -0.03 | 0.09 | -0.03 | 0.02 | -0.09 | 0.08 | 0.2 | 0.29 | -0.09 | -0.1 | -0.04 | -0.01 | 0.14 | 0.06 | 0.21 | -0.04 | NA | -0.069 (0.0043) | -0.105 (0.0036) | 0.126 (0.0034) | -0.034 (0.0047) |
| ***SUIC*** | 0.1 | 0.01 | -0.02 | -0.07 | 0.04 | 0.06 | -0.04 | 0.09 | -0.09 | 0.34 | 0.02 | -0.03 | -0.07 | -0.12 | 0.15 | 0.03 | 0.12 | 0.05 | -0.08 | NA | 0.04 (0.0027) | 0.064 (0.0022) | 0 (0) |
| ***TEAR*** | -0.01 | 0.02 | 0.03 | 0.07 | -0.05 | 0.07 | -0.1 | 0.14 | -0.03 | 0.12 | 0.12 | -0.01 | -0.05 | 0.04 | 0.22 | -0.05 | 0.03 | -0.13 | -0.11 | 0.05 | NA | -0.041 (0.0021) | 0 (0.0029) |
| ***TOB*** | 0.08 | -0.13 | -0.04 | 0.32 | 0.02 | 0.02 | 0.03 | 0.05 | 0.08 | -0.09 | 0.13 | 0.12 | 0.02 | 0.1 | -0.02 | 0.03 | -0.08 | -0.02 | 0.14 | 0.07 | -0.04 | NA | 0.075 (0.0026) |
| ***WGHT*** | -0.09 | -0.02 | 0.07 | 0.01 | 0.06 | 0.07 | -0.02 | 0.04 | 0.12 | 0.07 | -0.01 | 0 | 0.11 | 0.1 | -0.1 | -0.03 | 0.08 | 0.04 | -0.03 | 0.04 | 0 | 0.08 | NA |

eFigure 2 Un-thresholded SMD network graphs. (A) Temporal, (B) Contemporaneous and (C)Between-subject. Edge weights are not presented for visualisation pruposes but can be found in eTable 9.

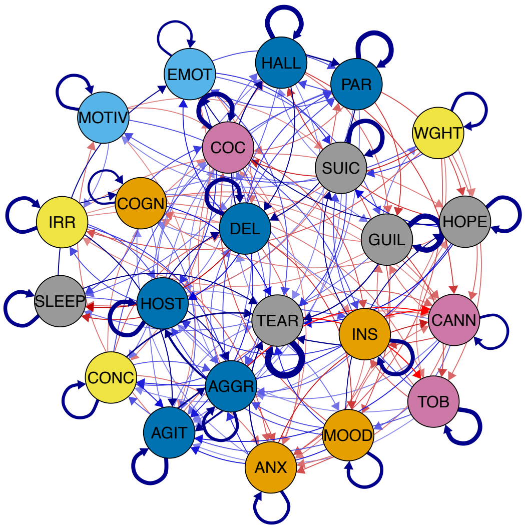

A.

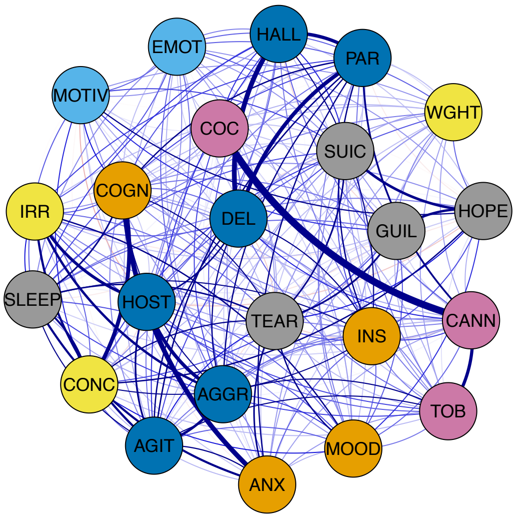

B.

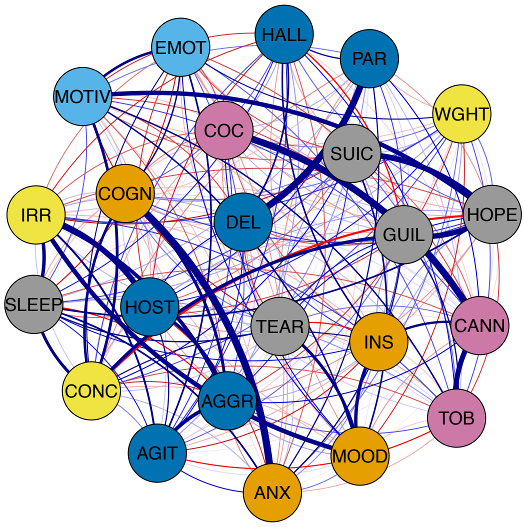

C.

eTable 10. SMD network centrality results.

For temporal networks, centrality was defined as the sum of absolute (directed) edge weights in (in-centrality, $c_{in}$) and out (out-centrality, $c_{out}$) of a node (including autocorrelative edges). For contemporaneous and between-subject networks, centrality was defined as the sum of absolute (undirected) edge weights for a node (autocorrelative edges do not exist).

| **Feature** | **Temporal (out-centrality)** | **Temporal (in-centrality)** | **Contemporaneous** | **Between-subject** |
|---|---|---|---|---|
| Aggression | 0.103 | 0.057 | 0.339 | 0.94 |
| Agitation | 0.033 | 0.061 | 0.198 | 0.252 |
| Anxiety | 0.026 | 0 | 0.268 | 0.419 |
| Cannabis use | 0 | 0.047 | 0.454 | 0.904 |
| Cocaine use | 0 | 0 | 0.248 | 0.4 |
| Cognitive impairment | 0 | 0.023 | 0.322 | 0.832 |
| Delusional thinking | 0.071 | 0.071 | 0.321 | 0.392 |
| Disturbed sleep | 0.048 | 0.023 | 0.177 | 0.458 |
| Emotional withdrawal | 0 | 0.047 | 0 | 0.197 |
| Feeling hopeless | 0.023 | 0.023 | 0.207 | 1.139 |
| Guilt | 0.026 | 0 | 0.1 | 0.564 |
| Hallucinations (all) | 0.053 | 0.026 | 0.305 | 0 |
| Hostility | 0.048 | 0.066 | 0.252 | 0.569 |
| Irritability | 0.047 | 0 | 0.311 | 1.124 |
| Mood instability | 0.023 | 0 | 0 | 0.745 |
| Paranoia | 0.024 | 0.03 | 0.357 | 0.392 |
| Poor concentration | 0.032 | 0 | 0.231 | 1.031 |
| Poor insight | 0.073 | 0 | 0 | 0.421 |
| Poor motivation | 0 | 0 | 0 | 0.693 |
| Suicidality | 0 | 0.023 | 0.107 | 0.344 |
| Tearfulness | 0.025 | 0.134 | 0 | 0.219 |
| Tobacco use | 0 | 0.025 | 0.125 | 0.318 |
| Weight loss | 0 | 0 | 0 | 0 |

eResults 3 SMD contemporaneous and between-subject network results

Inspection of associations between residuals, i.e., the partial correlation between nodes after controlling for time and between-subject effects (eFigure 3), showed that cannabis use-cocaine use shared the strongest positive relationship ($z$ = .25), followed by anxiety-cognitive impairment ($z$ = .18), and hallucinations (all)-delusional thinking ($z$ = .17). None of the negative relationships held after thresholding. Within this network, the most central items cannabis use ($c$ = .454), paranoia ($c$= .357) and aggression ($c$= .339) (eTable 10).

Between-individuals estimates (eFigure 3) suggested that anxiety-cognitive impairment had the strongest positive relationship ($z$ = .42), followed by cannabis-cocaine use (z = .40) and delusional thinking-paranoia ($z$ = .39). The only negative association that survived thresholding was feeling hopeless-poor concentration ($z$ = -.17). The most central items were feeling hopeless ($c$ = 1.139), irritability ($c$ = 1.124) and poor concentration ($c$ = 1.031) (eTable 10).

eFigure 3 SMD contemporaneous and between-subject network graphs.
Edges are displayed as lines, with the thickness representing the strength of the edge weight estimate (partial correlation coefficient, *z*).

**Contemporaneous network.**
**A.** Contemporaneous network displaying relationships between nodes from actual model estimates. Edges are thresholded (|z| > .8) and labelled (|z| > .10). **B.** Centrality measures for all nodes.

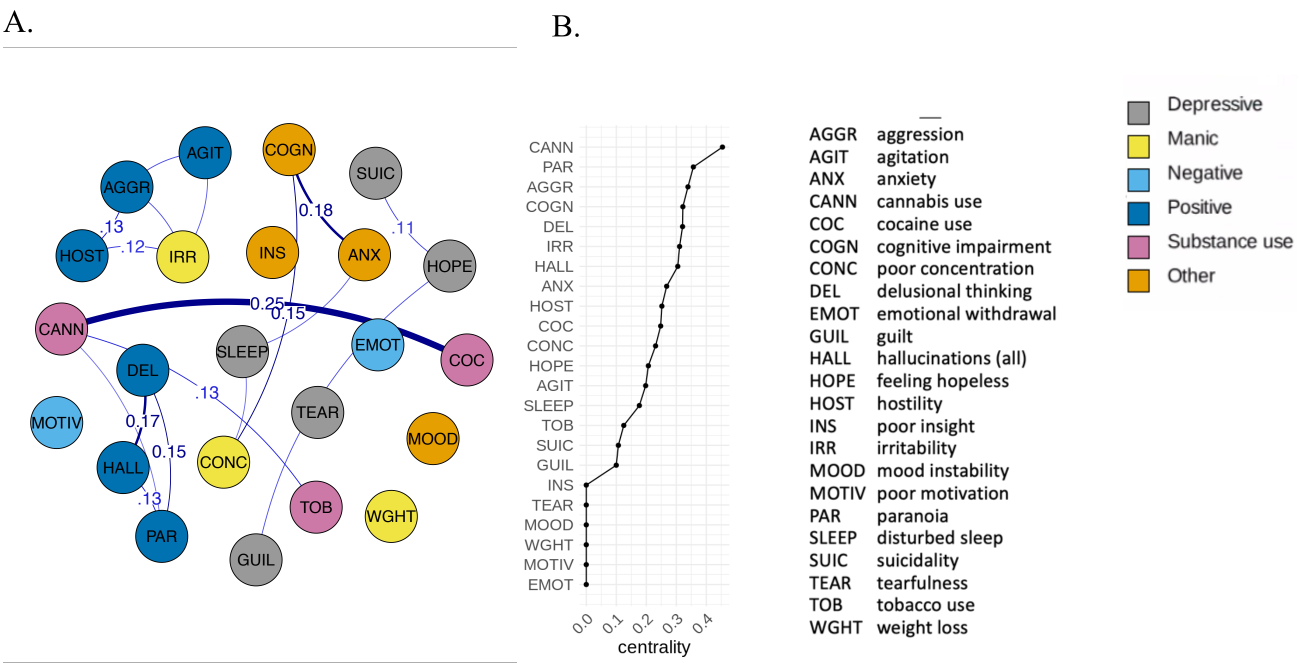

**Between-subject network.**
**A.** Between-subject network displaying relationships between nodes from actual model estimates. Edges are displayed as lines (|z| > .17) and are labelled (|z| > .25). Edge thickness represents the estimate magnitude (z). **B.** Centrality measures for all nodes

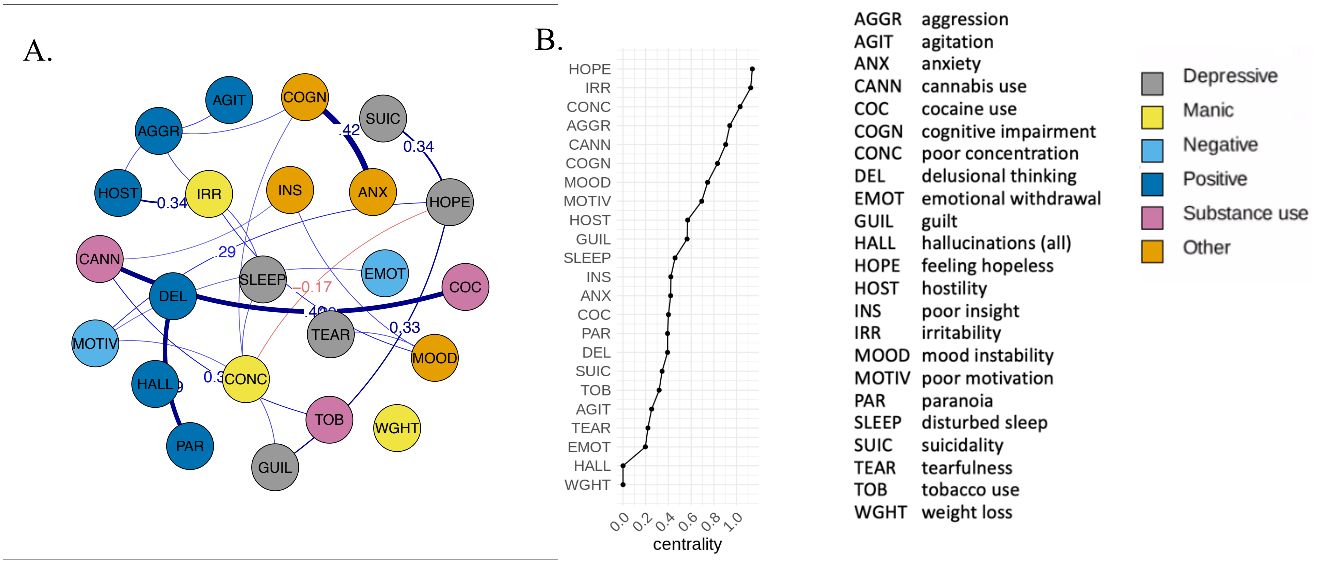

eMethods 6 Sub-networks node selection

(a) UMD

Out of the 61 NLP-derived prodromal features that were originally extracted from the EHRs, 39 features displayed near-zero variance and were excluded (see table below). Of the remaining 22 features, an additional variable (feeling hopeless) was excluded as it caused model instability and poor model fit, leaving a total of 21 features for the analyses: Aggression, Agitation, Anxiety, Cannabis use, Cognitive impairment, Disturbed sleep, Feeling lonely, Guilt, Hallucinations (all), Irritability, Low energy, Mood instability, Nightmares, Paranoia, Poor concentration, Poor insight, Poor motivation, Suicidality , Tearfulness, Tobacco use and Weight loss.

| **Follow-up interval** | **1** | **2** | **3** | **4** | **5** | **6** |
|---|---|---|---|---|---|---|
| Aggression | 13.4 | 13.1 | 10.4 | 15.1 | 14.5 | 14.1 |
| Agitation | 9.8 | 11.5 | 10.8 | 10.9 | 11.4 | 11.5 |
| **Anergia** | 342.3 | 685.7 | 186.3 | 256.5 | 227.9 | 227.9 |
| **Anhedonia** | 31.7 | 30.2 | 30.2 | 34.5 | 37.1 | 35.8 |
| Anxiety | 1 | 1.1 | 1.2 | 1.1 | 1.1 | 1 |
| **Apathy** | 411 | 146.1 | 136.3 | 127.8 | 170.7 | 146.1 |
| **Arousal** | 48 | 63.4 | 44.8 | 57.9 | 67.7 | 48 |
| **Bad dreams** | 127.8 | 120.2 | 113.4 | 205 | 127.8 | 186.3 |
| **Blunted affect** | 63.4 | 53.2 | 84.8 | 78.2 | 65.5 | 65.5 |
| Cannabis use | 18.8 | 16.2 | 17.4 | 19.4 | 17.9 | 17.2 |
| **Circumstantial speech** | 256.5 | 411 | 1029 | 514 | 342.3 | 411 |
| **Cocaine use** | 31.2 | 23.5 | 29.7 | 27.6 | 28 | 31.2 |
| Cognitive impairment | 1.5 | 1.3 | 1.2 | 1.3 | 1.4 | 1.5 |
| **Concrete thinking** | 2059 | 1029 | 685.7 | 2059 | 1029 | 685.7 |
| **Delusional thinking** | 42.8 | 37.9 | 51.8 | 81.4 | 33.9 | 54.7 |
| **Derailment of speech** | 1029 | 1029 | 2059 | 1029 | 685.7 | 1029 |
| Disturbed sleep | 3.2 | 2.9 | 2.8 | 3.1 | 3.4 | 3.7 |
| **Diurnal mood** | 107.4 | 186.3 | 342.3 | 186.3 | 186.3 | 256.5 |
| **Early morning** | 84.8 | 72.6 | 88.6 | 81.4 | 84.8 | 256.5 |
| **Echolalia** | 2059 | 2059 | 0 | 0 | 0 | 2059 |
| **Elation** | 102 | 102 | 205 | 127.8 | 97.1 | 205 |
| **Emotional withdrawn** | 26.5 | 23.2 | 25.8 | 31.2 | 30.2 | 37.9 |
| **Feeling helpless** | 38.6 | 51.8 | 34.5 | 35.1 | 37.1 | 45.8 |
| Feeling hopeless | 12.6 | 9.9 | 10.5 | 11.3 | 10.6 | 11.5 |
| Feeling lonely | 19.4 | 17.2 | 16.5 | 19.4 | 18.6 | 21.2 |

| | | | | | | |
|---|---|---|---|---|---|---|
| **Feeling worthless** | 32.2 | 29.3 | 38.6 | 41 | 35.8 | 38.6 |
| **Flight of ideas** | 256.5 | 411 | 342.3 | 1029 | 411 | 227.9 |
| **Formal thought disorder** | 685.7 | 0 | 0 | 1029 | 0 | 514 |
| **Grandiosity** | 342.3 | 411 | 411 | 685.7 | 342.3 | 342.3 |
| Guilt | 11.2 | 10.3 | 9.8 | 10 | 11 | 13.4 |
| Hallucinations (all) | 16 | 16.9 | 18.4 | 20.5 | 19.6 | 21.2 |
| **Hostility** | 54.7 | 61.4 | 53.2 | 61.4 | 65.5 | 72.6 |
| **Insomnia** | 17.2 | 16.3 | 16.5 | 19 | 20.9 | 20.9 |
| Irritability | 13.7 | 12.9 | 12.7 | 14.8 | 14.3 | 16.6 |
| **Loss of coherence** | 186.3 | 514 | 227.9 | 293.3 | 685.7 | 227.9 |
| Low energy | 17.4 | 17.1 | 15.3 | 17.4 | 20.5 | 19.8 |
| **MDMA use** | 342.3 | 411 | 514 | 2059 | 685.7 | 685.7 |
| Mood instability | 9.7 | 9.8 | 9.1 | 8.9 | 9.7 | 12.2 |
| **Mutism** | 256.5 | 186.3 | 685.7 | 293.3 | 342.3 | 342.3 |
| **Negative symptom** | 514 | 411 | 1029 | 685.7 | 685.7 | 1029 |
| Nightmare | 16.5 | 16.5 | 16.3 | 19.8 | 20.5 | 17.9 |
| Paranoia | 11.2 | 10.6 | 11.9 | 16 | 13.8 | 13.3 |
| **Passivity** | 2059 | 685.7 | 411 | 1029 | 0 | 2059 |
| **Poor appetite** | 37.1 | 24.1 | 24.8 | 32.8 | 35.8 | 30.2 |
| Poor concentration | 7 | 6.7 | 7.2 | 7.4 | 8.9 | 8.8 |
| Poor insight | 9.7 | 10.1 | 9.3 | 9.1 | 9.8 | 9.6 |
| Poor motivation | 15.9 | 13.8 | 15.7 | 15.3 | 15.2 | 16 |
| **Poverty of speech** | 685.7 | 2059 | 411 | 0 | 685.7 | 514 |
| **Poverty of thought** | 2059 | 0 | 0 | 0 | 1029 | 2059 |
| **Social withdrawal** | 81.4 | 75.3 | 88.6 | 120.2 | 170.7 | 107.4 |
| **Stupor** | 2059 | 0 | 0 | 0 | 0 | 0 |
| Suicidality | 10.6 | 9 | 9.1 | 10.5 | 11.1 | 11.3 |
| **Tangential speech** | 113.4 | 157.5 | 146.1 | 227.9 | 170.7 | 342.3 |
| Tearfulness | 4.7 | 4.1 | 4.4 | 4.9 | 5 | 5.8 |
| **Thought block** | 514 | 514 | 514 | 1029 | 2059 | 685.7 |
| **Thought broadcast** | 514 | 514 | 1029 | 685.7 | 1029 | 2059 |
| **Thought insertion** | 411 | 685.7 | 1029 | 514 | 514 | 685.7 |
| **Thought withdrawal** | 1029 | 2059 | 685.7 | 685.7 | 685.7 | 1029 |
| Tobacco use | 15.7 | 15.6 | 18.6 | 16.8 | 15.7 | 16.9 |
| **Waxy flexibility** | **NA** | **NA** | **NA** | **NA** | **NA** | **NA** |
| Weight loss | 17.1 | 15.1 | 16.3 | 18.4 | 17.9 | 23 |

(b) BMD

Out of the 61 NLP-derived prodromal features that were originally extracted from the EHRs, 42 features displayed near-zero variance and were excluded (see table below), leaving a total of 19 features for the analyses: Aggression, Agitation, Anxiety, Cannabis use, Cognitive impairment, Disturbed sleep, Elation, Feeling hopeless, Guilt, Hallucinations (all), Irritability, Mood instability, Paranoia, Poor concentration, Poor insight, Poor motivation, Suicidality , Tearfulness and Tobacco use.

| **Follow-up interval** | **1** | **2** | **3** | **4** | **5** | **6** |
|---|---|---|---|---|---|---|
| Aggression | 7.6 | 7.8 | 9 | 8.2 | 9.7 | 12 |
| Agitation | 5.8 | 6.2 | 5.7 | 6 | 6.5 | 6.9 |
| **Anergia** | 245.3 | 368.5 | 122.2 | 72.9 | 738 | 0 |
| **Anhedonia** | 31.1 | 28.6 | 26.4 | 27.4 | 72.9 | 48.3 |
| Anxiety | 1 | 1.1 | 1 | 1 | 1.1 | 1.2 |
| **Apathy** | 60.6 | 104.6 | 104.6 | 91.4 | 104.6 | 183.8 |
| **Arousal** | 22.8 | 31.1 | 28.6 | 24.5 | 22.1 | 37.9 |
| **Bad dreams** | 368.5 | 146.8 | 183.8 | 368.5 | 245.3 | 245.3 |
| **Blunted affect** | 66.2 | 60.6 | 81.1 | 91.4 | 81.1 | 81.1 |
| Cannabis use | 9 | 10.7 | 9.3 | 12.2 | 17 | 12.2 |
| **Circumstantial speech** | 81.1 | 81.1 | 91.4 | 91.4 | 368.5 | 368.5 |
| **Cocaine use** | 15.1 | 12.7 | 16.2 | 17 | 26.4 | 22.1 |
| Cognitive impairment | 1.4 | 1.3 | 1.5 | 1.6 | 1.7 | 1.6 |
| **Concrete thinking** | 738 | 368.5 | 738 | 738 | 738 | 738 |
| **Delusional thinking** | 15.1 | 15.4 | 20.1 | 28.6 | 32.6 | 20.1 |
| **Derailment of speech** | 183.8 | 738 | 368.5 | 368.5 | 183.8 | 0 |
| Disturbed sleep | 2.8 | 2.8 | 2.6 | 2.8 | 2.9 | 2.8 |
| **Diurnal mood** | 245.3 | 122.2 | 104.6 | 104.6 | 183.8 | 104.6 |
| **Early morning** | 122.2 | 81.1 | 81.1 | 104.6 | 122.2 | 368.5 |
| **Echolalia** | 0 | 738 | 738 | 0 | 0 | 0 |
| Elation | 9.6 | 10 | 9.9 | 11.1 | 11.5 | 9.9 |
| **Emotional withdrawn** | 16.6 | 17.5 | 16.6 | 24.5 | 22.1 | 32.6 |
| **Feeling helpless** | 66.2 | 81.1 | 81.1 | 81.1 | 60.6 | 51.8 |
| Feeling hopeless | 14.4 | 12.9 | 12.9 | 14.7 | 15.8 | 15.4 |
| **Feeling lonely** | 22.1 | 22.1 | 23.6 | 29.8 | 22.1 | 29.8 |
| **Feeling worthless** | 37.9 | 37.9 | 45.2 | 51.8 | 51.8 | 42.5 |
| **Flight of ideas** | 32.6 | 25.4 | 36 | 36 | 29.8 | 32.6 |

| | | | | | | |
|---|---|---|---|---|---|---|
| **Formal thought disorder** | 146.8 | 146.8 | 245.3 | 183.8 | 368.5 | 738 |
| **Grandiosity** | 20.7 | 19 | 27.4 | 32.6 | 27.4 | 28.6 |
| Guilt | 17 | 11.7 | 12.7 | 12.9 | 13.5 | 12.9 |
| Hallucinations (all) | 15.4 | 15.8 | 14.1 | 16.6 | 17.5 | 18.4 |
| **Hostility** | 26.4 | 29.8 | 21.4 | 23.6 | 32.6 | 28.6 |
| **Insomnia** | 22.1 | 17.5 | 15.1 | 15.4 | 17 | 16.6 |
| Irritability | 6.5 | 6.1 | 6.5 | 6.1 | 6.5 | 7 |
| **Loss of coherence** | 146.8 | 81.1 | 183.8 | 81.1 | 183.8 | 91.4 |
| **Low energy** | 16.2 | 20.1 | 12.4 | 21.4 | 23.6 | 27.4 |
| **MDMA use** | 245.3 | 245.3 | 368.5 | 368.5 | 368.5 | 368.5 |
| Mood instability | 5.9 | 4 | 4 | 4.2 | 5.1 | 4.7 |
| **Mutism** | 104.6 | 183.8 | 122.2 | 183.8 | 368.5 | 146.8 |
| **Negative symptom** | 368.5 | 104.6 | 183.8 | 183.8 | 183.8 | 146.8 |
| **Nightmare** | 42.5 | 37.9 | 40.1 | 37.9 | 51.8 | 60.6 |
| Paranoia | 6.5 | 6.8 | 7 | 8.7 | 9 | 8.6 |
| **Passivity** | 0 | 368.5 | 368.5 | 0 | 738 | 368.5 |
| **Poor appetite** | 29.8 | 28.6 | 29.8 | 42.5 | 45.2 | 31.1 |
| Poor concentration | 6.5 | 6.3 | 6.5 | 7.1 | 8.1 | 8.7 |
| Poor insight | 6.4 | 6.5 | 6.9 | 6.6 | 7.6 | 7.8 |
| Poor motivation | 13.2 | 12.7 | 13.2 | 12.7 | 17 | 17 |
| **Poverty of speech** | 738 | 738 | 368.5 | 368.5 | 368.5 | 245.3 |
| **Poverty of thought** | 368.5 | 738 | 738 | 0 | 738 | 0 |
| **Social withdrawal** | 60.6 | 66.2 | 40.1 | 91.4 | 104.6 | 81.1 |
| **Stupor** | 0 | 0 | 0 | 738 | 738 | 0 |
| Suicidality | 12.4 | 14.4 | 12 | 12.7 | 14.7 | 15.4 |
| **Tangential speech** | 36 | 55.8 | 122.2 | 60.6 | 122.2 | 146.8 |
| Tearfulness | 5.5 | 4.9 | 5.1 | 5.4 | 6 | 5.9 |
| **Thought block** | 0 | 738 | 183.8 | 738 | 0 | 738 |
| **Thought broadcast** | 183.8 | 368.5 | 183.8 | 0 | 368.5 | 245.3 |
| **Thought insertion** | 146.8 | 738 | 245.3 | 368.5 | 368.5 | 245.3 |
| **Thought withdrawal** | 368.5 | 0 | 0 | 738 | 738 | 0 |
| Tobacco use | 11.1 | 10.2 | 12.9 | 9.3 | 12.4 | 11.7 |
| **Waxy flexibility** | **NA** | **NA** | **NA** | **NA** | **NA** | **NA** |
| **Weight loss** | 18.4 | 17 | 17.9 | 21.4 | 26.4 | 19.5 |

(c) PSY

Out of the 61 NLP-derived prodromal features that were originally extracted from the EHRs, 37 features displayed near-zero variance and were excluded (see table below), leaving a total of 24 features for the analyses: Aggression, Agitation, Anxiety, Arousal, Cannabis use, Cocaine use, Cognitive impairment, Delusional thinking, Disturbed sleep, Elation, Emotional withdrawal, Feeling hopeless, Hallucinations (all), Hostility, Irritability, Mood instability, Paranoia, Poor concentration, Poor insight, Poor motivation , Suicidality, Tearfulness, Tobacco use, Weight loss.

| **Follow up interval** | **1** | **2** | **3** | **4** | **5** | **6** |
|---|---|---|---|---|---|---|
| Aggression | 6.7 | 6 | 5.3 | 5.6 | 6.3 | 5.5 |
| Agitation | 6 | 5.7 | 5.9 | 5.3 | 5.4 | 5.2 |
| **Anergia** | 364.6 | 405.2 | 521.3 | 405.2 | 608.3 | 521.3 |
| **Anhedonia** | 82.1 | 50.5 | 49.1 | 65.5 | 57 | 100.6 |
| Anxiety | 1.4 | 1.2 | 1.3 | 1.2 | 1.2 | 1.2 |
| **Apathy** | 109.8 | 64.3 | 97.8 | 120.9 | 116.9 | 88.2 |
| Arousal | 19.9 | 17.9 | 19.5 | 18.8 | 19.9 | 16.8 |
| **bad dreams** | 151.3 | 165.2 | 165.2 | 242.7 | 202.1 | 181.8 |
| **Blunted affect** | 56.1 | 35.6 | 38.3 | 49.1 | 49.8 | 44.7 |
| Cannabis use | 7.2 | 6.1 | 6.3 | 5.8 | 6.1 | 6 |
| **Circumstantial speech** | 80.2 | 59.9 | 72.1 | 95.2 | 76.8 | 75.2 |
| Cocaine use | 14.5 | 13.2 | 13.6 | 12.8 | 13.9 | 13.3 |
| Cognitive impairment | 1.5 | 1.4 | 1.3 | 1.3 | 1.3 | 1.3 |
| **Concrete thinking** | 260.1 | 181.8 | 214.1 | 227.5 | 214.1 | 165.2 |
| Delusional thinking | 5.9 | 5.4 | 5.8 | 5.6 | 5.3 | 5.6 |
| **Derailment of speech** | 158 | 202.1 | 260.1 | 165.2 | 202.1 | 191.4 |
| Disturbed sleep | 3.8 | 3.6 | 3.5 | 3.6 | 3.5 | 3.8 |
| **Diurnal mood** | 405.2 | 730.2 | 521.3 | 456 | 303.7 | 405.2 |
| **Early morning** | 227.5 | 260.1 | 242.7 | 280.2 | 280.2 | 260.1 |
| **Echolalia** | 1217.7 | 913 | 913 | 1217.7 | 1217.7 | 3655 |
| Elation | 20.6 | 21 | 20.4 | 21.3 | 20.4 | 19.9 |
| Emotional withdrawn | 18.2 | 16.1 | 17.9 | 17.7 | 20.6 | 17.8 |
| **Feeling helpless** | 95.2 | 97.8 | 82.1 | 65.5 | 72.1 | 95.2 |
| Feeling hopeless | 23.5 | 19.2 | 21.9 | 22.6 | 23.4 | 21.3 |
| **Feeling lonely** | 23.1 | 23.5 | 24.4 | 21.2 | 23.5 | 22 |
| **Feeling worthless** | 88.2 | 82.1 | 84 | 84 | 84 | 116.9 |
| **Flight of ideas** | 50.5 | 43 | 52 | 49.1 | 47.7 | 47.7 |

| | | | | | | |
|---|---|---|---|---|---|---|
| **Formal thought disorder** | 181.8 | 145.2 | 145.2 | 151.3 | 165.2 | 92.7 |
| **Grandiosity** | 23.4 | 20.5 | 22.9 | 22.9 | 21.9 | 22 |
| **Guilt** | 20.5 | 20.6 | 20.1 | 25.7 | 23.1 | 21.6 |
| Hallucinations (all) | 4.7 | 4.3 | 4.3 | 4.3 | 4.3 | 4.1 |
| Hostility | 14.8 | 13.5 | 12.6 | 13.7 | 13 | 13.1 |
| **Insomnia** | 31.9 | 24.4 | 29.5 | 29 | 27.1 | 31.9 |
| Irritability | 7.4 | 7.2 | 7.1 | 7.1 | 7 | 6.8 |
| **Loss of coherence** | 59.9 | 55.2 | 46.5 | 44.1 | 52.8 | 47.1 |
| **Low energy** | 46.5 | 35.2 | 35.2 | 39.6 | 49.1 | 51.2 |
| **MDMA use** | 521.3 | 331.4 | 608.3 | 364.6 | 405.2 | 280.2 |
| Mood instability | 9.4 | 9.8 | 9.4 | 9.6 | 10.2 | 10 |
| **Mutism** | 64.3 | 62 | 61 | 76.8 | 70.7 | 63.1 |
| **Negative symptom** | 37.9 | 32.2 | 27.1 | 27.6 | 29.2 | 28.5 |
| **Nightmare** | 39.2 | 33.8 | 30.5 | 37.1 | 37.5 | 37.1 |
| Paranoia | 3 | 2.6 | 2.5 | 2.5 | 2.4 | 2.4 |
| **Passivity** | 120.9 | 73.6 | 70.7 | 84 | 86 | 106.5 |
| **Poor appetite** | 31.9 | 26.9 | 30.8 | 39.6 | 33.2 | 33.5 |
| Poor concentration | 9.3 | 8.1 | 9.4 | 9.6 | 10.8 | 9.8 |
| Poor insight | 7.1 | 6 | 6.2 | 6.9 | 6.6 | 6.3 |
| Poor motivation | 16.7 | 15.9 | 13.8 | 15.8 | 15.7 | 16.2 |
| **Poverty of speech** | 173.1 | 129.6 | 242.7 | 165.2 | 165.2 | 145.2 |
| **Poverty of thought** | 260.1 | 173.1 | 364.6 | 303.7 | 280.2 | 260.1 |
| **Social withdrawal** | 66.7 | 49.8 | 50.5 | 55.2 | 80.2 | 52 |
| **Stupor** | 730.2 | 1217.7 | 456 | 730.2 | 3655 | 913 |
| Suicidality | 16.4 | 15.2 | 16.4 | 17.9 | 16.8 | 20 |
| **Tangential speech** | 35.2 | 29.5 | 40.1 | 39.2 | 35.9 | 31.6 |
| Tearfulness | 10.2 | 9.4 | 9.9 | 10.2 | 10.3 | 11.8 |
| **Thought block** | 92.7 | 63.1 | 125.1 | 116.9 | 90.4 | 109.8 |
| **Thought broadcast** | 106.5 | 76.8 | 100.6 | 72.1 | 84 | 75.2 |
| **Thought insertion** | 106.5 | 73.6 | 73.6 | 92.7 | 84 | 72.1 |
| **Thought withdrawal** | 227.5 | 158 | 173.1 | 214.1 | 202.1 | 214.1 |
| Tobacco use | 8.2 | 7.3 | 7.3 | 7.2 | 6.9 | 6.6 |
| **Waxy flexibility** | **NA** | **NA** | **NA** | **NA** | **NA** | 3655 |
| Weight loss | 15.8 | 15 | 16.5 | 16.7 | 14.6 | 14.3 |

eResults 4 Sub-networks model fit

Saturated networks demonstrated excellent fit (UMD: RMSEA=.013 [95%CI: .012, .014]; $X^2(7203)=9729.45$, p<.0001; CFI=.90; TLI=.90 ; BMD: RMSEA=.023 [95%CI: .022, .024]; $X^2(5909)=8219.85$, p<.0001; CFI=.83; TLI=.81 ; PSY: RMSEA=.013 [95%CI: .013, .013]; $X^2(9384)=15171.76$, p<.0001; CFI=.94; TLI=.93).

Simulated data using the estimated model structure was generated and refitted to estimate recoverability, demonstrating excellent fit for UMD (RMSEA=.0019 [95%CI: <.0001, .0033]; $X^2(7203)=7305.68$, p=0.20; CFI=1.0; TLI=1.0), BMD (RMSEA=.0021 [95%CI: <.0001, .0035]; $X^2(5909)=6010.49$, p=.18; CFI=1.0; TLI=1.0) and PSY (RMSEA=.0030 [95%CI: .0017, .0039]; $X^2(9384)=9716.10$, p=.0082; CFI=1.0; TLI=1.0).

eTable 11 Temporal sub-network edge estimates. (A) Actual model (green) and (B) bootstrapped estimates (blue, standard error of the mean). Any bootstrapped edges that had 95% CI crossing zero were forced to 0. Edge direction is defined from the node in the column to the node in the row.

**a. UMD**

| | AGGR | AGIT | ANX | CANN | COG | SLEEP | LONE | GUILT | HALLL | IRR | ENERGY | MOOD | NIGHT | PAR | CONC | INS | MOTIV | SUIC | TEAR | TOB | WGHT |
|---|---|---|---|---|---|---|---|---|---|---|---|---|---|---|---|---|---|---|---|---|---|
| ***AGGR*** | 0.07 | 0.03 | 0 | 0.01 | 0 | -0.01 | -0.01 | -0.01 | 0 | -0.01 | 0.01 | 0.03 | 0.01 | 0.01 | -0.01 | 0.02 | 0 | 0 | -0.01 | -0.01 | 0.01 |
| ***AGIT*** | 0.02 | 0.06 | -0.01 | 0 | 0.01 | -0.01 | -0.03 | 0.02 | 0 | 0 | 0.01 | 0.02 | 0 | 0.01 | 0.01 | -0.01 | -0.01 | 0.02 | 0 | -0.02 | 0.03 |
| ***ANX*** | 0.03 | 0.02 | 0.08 | -0.01 | 0.01 | 0 | 0 | 0.01 | 0 | -0.01 | 0.02 | 0.02 | 0 | 0 | 0 | 0.01 | -0.01 | 0 | 0.02 | -0.01 | -0.01 |
| ***CANN*** | 0 | 0.01 | -0.01 | 0.1 | 0.02 | 0 | -0.03 | -0.01 | -0.01 | 0.01 | 0.01 | 0.03 | -0.02 | 0.03 | -0.02 | -0.01 | 0 | 0.02 | 0 | 0 | -0.01 |
| ***COG*** | 0.01 | 0.01 | 0.02 | 0 | 0.03 | 0 | 0 | 0.02 | 0 | 0 | 0.03 | 0.01 | 0.01 | 0.01 | 0.02 | -0.01 | 0.02 | 0 | 0 | -0.02 | 0.01 |
| ***SLEEP*** | 0 | 0.01 | -0.02 | 0.01 | -0.01 | 0.08 | -0.02 | 0.01 | 0.01 | -0.01 | 0.03 | -0.01 | 0 | 0 | 0 | 0.01 | 0.02 | 0 | 0.03 | 0 | -0.01 |
| ***LONE*** | 0 | -0.01 | 0.02 | 0.01 | 0.03 | -0.01 | 0.07 | -0.01 | -0.02 | -0.01 | -0.01 | 0 | 0.01 | 0.01 | -0.03 | -0.01 | -0.01 | 0 | 0.01 | 0 | -0.01 |
| ***GUILT*** | 0.01 | 0.01 | -0.01 | -0.03 | 0.03 | 0.01 | 0 | 0.11 | 0 | 0 | 0.04 | 0.01 | 0 | 0 | 0 | 0 | 0 | 0 | 0.02 | -0.01 | 0.01 |
| ***HALLL*** | 0.02 | 0.01 | -0.03 | -0.03 | 0 | 0 | 0.01 | -0.02 | 0.11 | 0.02 | 0.02 | 0.03 | 0 | 0 | 0.01 | -0.01 | 0 | 0.01 | 0.02 | -0.01 | -0.01 |
| ***IRR*** | 0.03 | 0.02 | -0.01 | 0.01 | 0.02 | -0.02 | -0.01 | -0.02 | 0.01 | 0.06 | 0.02 | -0.02 | 0.02 | 0 | 0 | -0.02 | 0 | 0.02 | 0.01 | 0 | -0.02 |
| ***ENERGY*** | 0.01 | 0.02 | 0.01 | 0.02 | -0.01 | -0.02 | -0.01 | -0.01 | 0.01 | 0.01 | 0.05 | 0.01 | 0 | 0.02 | 0.04 | -0.02 | 0.02 | 0 | 0.02 | 0.01 | 0 |
| ***MOOD*** | 0.04 | 0.01 | -0.01 | -0.01 | 0.02 | -0.01 | 0 | 0.01 | 0.01 | -0.02 | 0.03 | 0.07 | 0.02 | -0.02 | 0.01 | 0.01 | -0.01 | 0.01 | 0.03 | 0.01 | 0.02 |
| ***NIGHT*** | 0.03 | -0.01 | 0.01 | -0.01 | -0.02 | 0 | 0.01 | 0.02 | 0.01 | 0.01 | 0.02 | 0.01 | 0.07 | -0.02 | 0 | 0 | 0.02 | -0.01 | 0.01 | 0 | 0 |
| ***PAR*** | 0 | 0 | 0 | 0.01 | -0.01 | -0.01 | 0.01 | -0.04 | 0.02 | 0.03 | 0.02 | 0.01 | 0.02 | 0.07 | 0.02 | -0.02 | 0.01 | -0.02 | 0 | 0.01 | 0.01 |
| ***CONC*** | 0.01 | 0.01 | -0.02 | 0.01 | 0 | 0 | -0.05 | 0 | 0.01 | -0.01 | 0.02 | 0.02 | -0.01 | 0.02 | 0.07 | 0 | 0.03 | 0 | 0.04 | -0.01 | 0 |
| ***INS*** | 0 | 0.02 | -0.01 | -0.01 | -0.01 | -0.01 | 0 | -0.01 | 0 | 0.01 | 0.02 | 0.02 | -0.01 | -0.01 | 0.02 | 0.09 | 0 | 0.01 | 0.02 | 0.01 | -0.01 |
| ***MOTIV*** | 0 | -0.03 | -0.01 | 0.02 | 0.02 | -0.01 | -0.01 | 0.01 | 0.01 | 0 | 0.05 | 0.02 | -0.01 | -0.01 | 0.01 | 0 | 0.05 | 0 | 0.02 | 0 | 0.02 |

| | | | | | | | | | | | | | | | | | | | | | |
|---|---|---|---|---|---|---|---|---|---|---|---|---|---|---|---|---|---|---|---|---|---|
| ***SUIC*** | -0.02 | 0.01 | 0 | -0.01 | -0.01 | -0.01 | -0.01 | 0 | 0.03 | -0.01 | -0.01 | -0.02 | -0.02 | -0.03 | 0.01 | -0.02 | 0.02 | 0.07 | 0.02 | 0.01 | 0.01 |
| ***TEAR*** | 0.01 | 0 | 0.01 | 0 | 0 | -0.01 | -0.02 | 0 | 0.02 | 0.01 | 0.01 | -0.01 | 0.02 | -0.01 | 0.02 | 0.02 | 0.03 | 0.01 | 0.1 | -0.02 | 0 |
| ***TOB*** | 0.03 | 0 | -0.02 | 0 | 0.01 | 0 | -0.02 | 0 | 0.01 | 0 | 0.01 | 0.01 | 0 | 0 | -0.02 | -0.02 | 0.01 | 0.04 | 0 | 0.07 | 0 |
| ***WGHT*** | -0.02 | 0.01 | -0.01 | 0 | 0.01 | -0.01 | -0.01 | 0.01 | 0.01 | -0.01 | 0 | 0 | 0 | -0.01 | 0.01 | 0.01 | 0.02 | 0.01 | 0.02 | 0 | 0.03 |

| | AGGR | AGIT | ANX | CANN | COG | SLEEP | LONE | GUILT | HALLL | IRR | ENERGY | MOOD | NIGHT | PAR | CONC | INS | MOTIV | SUIC | TEAR | TOB | WGHT |
|---|---|---|---|---|---|---|---|---|---|---|---|---|---|---|---|---|---|---|---|---|---|
| *AGGR* | 0.063 (9e-04) | 0.031 (6e-04) | -0.004 (5e-04) | 0.009 (6e-04) | -0.004 (5e-04) | -0.006 (5e-04) | -0.011 (5e-04) | -0.008 (5e-04) | -0.004 (6e-04) | -0.011 (5e-04) | 0.007 (6e-04) | 0.028 (6e-04) | 0.008 (5e-04) | 0.014 (6e-04) | -0.006 (6e-04) | 0.018 (5e-04) | 0.002 (5e-04) | 0 (0) | -0.012 (6e-04) | -0.009 (5e-04) | 0.013 (5e-04) |
| *AGIT* | 0.016 (6e-04) | 0.056 (8e-04) | -0.014 (5e-04) | 0.004 (6e-04) | 0.015 (5e-04) | -0.005 (5e-04) | -0.026 (6e-04) | 0.015 (5e-04) | 0.004 (6e-04) | 0 (0) | 0.006 (5e-04) | 0.019 (5e-04) | -0.006 (5e-04) | 0.005 (6e-04) | 0.009 (5e-04) | -0.01 (5e-04) | -0.011 (5e-04) | 0.018 (6e-04) | 0.003 (6e-04) | -0.021 (5e-04) | 0.024 (6e-04) |
| *ANX* | 0.028 (5e-04) | 0.016 (5e-04) | 0.07 (8e-04) | -0.007 (5e-04) | 0.004 (5e-04) | 0.004 (5e-04) | 0.002 (5e-04) | 0.005 (4e-04) | 0 (0) | -0.011 (4e-04) | 0.015 (5e-04) | 0.018 (5e-04) | 0.002 (5e-04) | 0.002 (5e-04) | 0.005 (5e-04) | 0.013 (5e-04) | -0.008 (5e-04) | -0.005 (5e-04) | 0.023 (5e-04) | -0.006 (4e-04) | -0.01 (5e-04) |
| *CANN* | -0.002 (5e-04) | 0.011 (5e-04) | -0.014 (4e-04) | 0.102 (0.0011) | 0.018 (5e-04) | 0.001 (5e-04) | -0.032 (5e-04) | -0.015 (5e-04) | -0.012 (5e-04) | 0.014 (5e-04) | 0.005 (5e-04) | 0.028 (5e-04) | -0.021 (5e-04) | 0.032 (6e-04) | -0.016 (5e-04) | -0.008 (4e-04) | 0.001 (4e-04) | 0.021 (6e-04) | -0.002 (5e-04) | 0.001 (6e-04) | -0.013 (5e-04) |
| *COG* | 0.01 (5e-04) | 0.009 (5e-04) | 0.015 (6e-04) | 0.006 (5e-04) | 0.031 (6e-04) | 0.002 (5e-04) | -0.001 (6e-04) | 0.023 (5e-04) | 0.003 (6e-04) | 0.005 (5e-04) | 0.029 (6e-04) | 0.013 (5e-04) | 0.008 (5e-04) | 0.008 (5e-04) | 0.014 (5e-04) | -0.011 (4e-04) | 0.02 (5e-04) | 0.005 (5e-04) | 0 (0) | -0.024 (5e-04) | 0.012 (5e-04) |
| *SLEEP* | -0.003 (6e-04) | 0.016 (6e-04) | -0.015 (5e-04) | 0.011 (5e-04) | -0.005 (5e-04) | 0.078 (7e-04) | -0.018 (5e-04) | 0.007 (5e-04) | 0.006 (6e-04) | -0.01 (5e-04) | 0.026 (6e-04) | -0.006 (5e-04) | -0.002 (5e-04) | 0.004 (5e-04) | 0 (5e-04) | 0.013 (5e-04) | 0.016 (5e-04) | 0 (0) | 0.023 (6e-04) | -0.002 (5e-04) | 0 (0) |
| *LONE* | 0 (5e-04) | -0.012 (6e-04) | 0.018 (5e-04) | 0.015 (6e-04) | 0.029 (6e-04) | -0.008 (5e-04) | 0.065 (0.001) | -0.004 (5e-04) | -0.015 (5e-04) | -0.005 (5e-04) | -0.015 (4e-04) | 0.005 (5e-04) | 0.013 (5e-04) | 0.014 (6e-04) | -0.026 (6e-04) | 0 (0) | -0.006 (4e-04) | -0.003 (6e-04) | 0.009 (6e-04) | 0.002 (5e-04) | -0.007 (5e-04) |
| *GUILT* | 0.007 (6e-04) | 0.004 (5e-04) | -0.011 (5e-04) | -0.03 (5e-04) | 0.025 (5e-04) | 0.008 (5e-04) | 0.003 (6e-04) | 0.104 (0.001) | 0.003 (5e-04) | -0.003 (4e-04) | 0.037 (6e-04) | 0.007 (5e-04) | 0 (0) | 0.002 (6e-04) | -0.004 (6e-04) | 0 (5e-04) | 0.003 (5e-04) | -0.002 (6e-04) | 0.023 (5e-04) | -0.012 (5e-04) | 0.008 (5e-04) |
| *HALLL* | 0.019 (6e-04) | 0.008 (6e-04) | -0.026 (5e-04) | -0.033 (6e-04) | -0.002 (5e-04) | -0.004 (5e-04) | 0.007 (5e-04) | -0.019 (4e-04) | 0.104 (0.0011) | 0.02 (4e-04) | 0.016 (6e-04) | 0.025 (5e-04) | 0 (5e-04) | 0.003 (7e-04) | 0.015 (6e-04) | -0.013 (5e-04) | 0.003 (5e-04) | 0.006 (6e-04) | 0.025 (5e-04) | -0.01 (5e-04) | -0.009 (5e-04) |
| *IRR* | 0.032 (7e-04) | 0.021 (6e-04) | -0.012 (5e-04) | 0.014 (6e-04) | 0.023 (5e-04) | -0.015 (5e-04) | -0.007 (5e-04) | -0.024 (5e-04) | 0.016 (6e-04) | 0.055 (8e-04) | 0.018 (6e-04) | -0.015 (6e-04) | 0.018 (5e-04) | -0.004 (5e-04) | 0 (0) | -0.023 (5e-04) | 0 (0) | 0.014 (6e-04) | 0.005 (5e-04) | -0.003 (5e-04) | -0.018 (5e-04) |
| *ENERGY* | 0 (0) | 0.018 (5e-04) | 0.01 (5e-04) | 0.018 (6e-04) | -0.005 (5e-04) | -0.015 (5e-04) | -0.01 (5e-04) | -0.012 (6e-04) | 0.014 (6e-04) | 0.008 (5e-04) | 0.044 (7e-04) | 0.008 (5e-04) | -0.004 (5e-04) | 0.017 (4e-04) | 0.039 (6e-04) | -0.017 (6e-04) | 0.016 (6e-04) | 0.002 (5e-04) | 0.021 (6e-04) | 0.007 (5e-04) | -0.004 (6e-04) |
| *MOOD* | 0.041 (6e-04) | 0.01 (5e-04) | -0.011 (5e-04) | -0.007 (6e-04) | 0.015 (5e-04) | -0.01 (5e-04) | 0 (5e-04) | 0.013 (5e-04) | 0.005 (6e-04) | -0.021 (5e-04) | 0.03 (6e-04) | 0.068 (8e-04) | 0.015 (6e-04) | -0.022 (6e-04) | 0.015 (5e-04) | 0.003 (5e-04) | -0.009 (6e-04) | 0.007 (6e-04) | 0.025 (5e-04) | 0.006 (6e-04) | 0.024 (5e-04) |
| *NIGHT* | 0.024 (5e-04) | -0.007 (5e-04) | 0.007 (5e-04) | -0.016 (4e-04) | -0.015 (5e-04) | 0.003 (5e-04) | 0 (0) | 0.018 (5e-04) | 0.006 (6e-04) | 0.01 (5e-04) | 0.022 (6e-04) | 0.004 (5e-04) | 0.072 (9e-04) | -0.018 (5e-04) | 0.003 (5e-04) | -0.004 (5e-04) | 0.016 (5e-04) | -0.007 (5e-04) | 0.015 (5e-04) | 0.003 (5e-04) | 0 (5e-04) |
| *PAR* | 0 (0) | 0.003 (5e-04) | -0.005 (4e-04) | 0.015 (6e-04) | -0.01 (5e-04) | -0.014 (5e-04) | 0.007 (6e-04) | -0.034 (5e-04) | 0.023 (7e-04) | 0.029 (5e-04) | 0.022 (5e-04) | 0.009 (5e-04) | 0.016 (5e-04) | 0.069 (9e-04) | 0.017 (6e-04) | -0.022 (5e-04) | 0.008 (5e-04) | -0.023 (5e-04) | 0.002 (5e-04) | 0.003 (5e-04) | 0.008 (5e-04) |
| *CONC* | 0.01 (5e-04) | 0.013 (5e-04) | -0.02 (5e-04) | 0.006 (6e-04) | -0.005 (5e-04) | -0.003 (5e-04) | -0.05 (6e-04) | -0.002 (5e-04) | 0.011 (6e-04) | -0.013 (5e-04) | 0.02 (6e-04) | 0.019 (5e-04) | -0.006 (6e-04) | 0.017 (6e-04) | 0.069 (8e-04) | -0.005 (5e-04) | 0.025 (6e-04) | 0 (6e-04) | 0.036 (6e-04) | -0.014 (6e-04) | 0 (0) |

| | | | | | | | | | | | | | | | | | | | | | |
|---|---|---|---|---|---|---|---|---|---|---|---|---|---|---|---|---|---|---|---|---|---|
| ***INS*** | 0 (0) | 0.021 (5e-04) | -0.004 (5e-04) | -0.009 (5e-04) | -0.006 (6e-04) | -0.007 (5e-04) | -0.003 (6e-04) | -0.006 (5e-04) | 0.002 (5e-04) | 0.006 (5e-04) | 0.02 (6e-04) | 0.018 (5e-04) | -0.009 (6e-04) | -0.008 (6e-04) | 0.02 (6e-04) | 0.091 (9e-04) | 0.003 (6e-04) | 0.009 (6e-04) | 0.022 (5e-04) | 0.014 (5e-04) | -0.016 (4e-04) |
| ***MOTIV*** | 0 (6e-04) | -0.033 (5e-04) | -0.009 (5e-04) | 0.019 (7e-04) | 0.017 (6e-04) | -0.013 (5e-04) | -0.011 (6e-04) | 0.004 (5e-04) | 0.009 (6e-04) | 0.004 (5e-04) | 0.045 (6e-04) | 0.015 (5e-04) | -0.01 (5e-04) | -0.006 (6e-04) | 0 (0) | 0.004 (5e-04) | 0.045 (7e-04) | -0.005 (5e-04) | 0.015 (6e-04) | -0.003 (5e-04) | 0.02 (6e-04) |
| ***SUIC*** | -0.022 (6e-04) | 0.01 (6e-04) | 0.003 (5e-04) | -0.007 (5e-04) | -0.006 (5e-04) | -0.006 (5e-04) | -0.012 (5e-04) | -0.005 (5e-04) | 0.028 (7e-04) | -0.007 (5e-04) | -0.007 (5e-04) | -0.016 (5e-04) | -0.015 (6e-04) | -0.032 (6e-04) | 0.005 (5e-04) | -0.016 (5e-04) | 0.018 (5e-04) | 0.066 (8e-04) | 0.015 (6e-04) | 0.009 (5e-04) | 0.013 (5e-04) |
| ***TEAR*** | 0.005 (6e-04) | -0.002 (5e-04) | 0.006 (5e-04) | 0.001 (5e-04) | 0 (0) | -0.01 (5e-04) | -0.014 (5e-04) | 0 (0) | 0.019 (5e-04) | 0.006 (5e-04) | 0.008 (6e-04) | -0.01 (5e-04) | 0.016 (6e-04) | -0.004 (5e-04) | 0.022 (6e-04) | 0.02 (5e-04) | 0.026 (5e-04) | 0.005 (6e-04) | 0.102 (0.001) | -0.023 (5e-04) | 0 (0) |
| ***TOB*** | 0.026 (5e-04) | 0 (0) | -0.019 (5e-04) | 0.005 (7e-04) | 0.013 (5e-04) | -0.004 (4e-04) | -0.02 (6e-04) | 0 (0) | 0.009 (6e-04) | 0.001 (5e-04) | 0.009 (5e-04) | 0.009 (5e-04) | -0.001 (5e-04) | -0.003 (6e-04) | -0.024 (5e-04) | -0.018 (5e-04) | 0.011 (5e-04) | 0.042 (6e-04) | 0.004 (5e-04) | 0.067 (9e-04) | 0 (0) |
| ***WGHT*** | -0.015 (5e-04) | 0.011 (5e-04) | -0.01 (5e-04) | -0.002 (6e-04) | 0.009 (5e-04) | -0.013 (5e-04) | -0.011 (5e-04) | 0.007 (5e-04) | 0.014 (6e-04) | -0.012 (5e-04) | 0.001 (5e-04) | 0.001 (4e-04) | 0 (0) | -0.007 (5e-04) | 0.011 (5e-04) | 0.008 (5e-04) | 0.022 (5e-04) | 0.007 (5e-04) | 0.025 (6e-04) | 0 (0) | 0.03 (8e-04) |

### b. BMD

| | AGGR | AGIT | ANX | CANN | COGN | SLEEP | ELAT | HOPE | GUILT | HALL | IRR | MOOD | PAR | CONC | INS | MOTIV | SUIC | TEAR | TOB |
|---|---|---|---|---|---|---|---|---|---|---|---|---|---|---|---|---|---|---|---|
| *AGGR* | 0.09 | 0.04 | 0 | -0.01 | 0.01 | 0.02 | 0.05 | -0.03 | -0.02 | 0.02 | -0.01 | 0.01 | 0.01 | 0.02 | -0.01 | 0 | -0.02 | -0.01 | -0.06 |
| *AGIT* | 0.05 | 0.08 | -0.01 | -0.02 | -0.01 | 0.04 | 0.03 | 0.01 | 0.01 | 0.01 | -0.02 | 0 | 0.02 | -0.01 | 0.01 | 0 | -0.01 | 0.04 | 0 |
| *ANX* | 0 | 0 | 0.01 | -0.05 | 0.07 | -0.02 | 0.01 | -0.01 | -0.02 | 0.01 | -0.01 | 0.05 | 0.05 | -0.02 | 0 | -0.01 | 0.02 | 0.04 | 0.01 |
| *CANN* | 0.05 | 0.03 | 0.02 | 0.02 | -0.04 | 0.02 | -0.05 | 0.02 | 0 | -0.01 | 0 | 0.02 | 0.01 | -0.03 | 0 | -0.01 | -0.01 | 0.01 | -0.02 |
| *COGN* | 0.01 | 0.02 | 0.03 | -0.02 | 0.02 | 0.01 | 0.02 | 0 | 0 | 0 | -0.04 | 0.01 | 0.05 | 0.03 | 0 | -0.02 | 0.01 | 0.05 | -0.01 |
| *SLEEP* | 0.01 | 0.03 | 0.02 | 0.05 | 0 | 0.07 | 0.02 | 0.01 | 0 | 0.01 | -0.01 | 0 | 0.01 | 0.02 | 0 | 0.02 | 0.03 | 0.05 | -0.02 |
| *ELAT* | 0.03 | 0.07 | -0.01 | -0.06 | 0 | 0 | 0.12 | -0.01 | -0.04 | 0.01 | 0.04 | -0.02 | 0.08 | 0.03 | -0.02 | -0.02 | 0 | 0.01 | 0 |
| *HOPE* | -0.02 | 0.05 | 0.02 | 0 | -0.05 | 0.03 | -0.06 | 0.12 | -0.01 | -0.05 | 0 | 0 | -0.01 | 0.03 | 0.04 | -0.02 | 0.03 | 0.03 | 0.01 |
| *GUILT* | -0.01 | -0.01 | 0 | 0.01 | -0.02 | 0.01 | 0.01 | 0.04 | 0.04 | 0 | 0 | 0.02 | -0.03 | 0 | 0 | 0.03 | 0 | -0.01 | 0.02 |
| *HALL* | 0.02 | -0.01 | -0.01 | -0.01 | 0 | -0.01 | 0.01 | 0 | -0.02 | 0.08 | -0.01 | -0.02 | 0.1 | 0.03 | 0.02 | 0.01 | 0.02 | -0.02 | -0.02 |
| *IRR* | 0.03 | 0 | 0.01 | -0.05 | 0.05 | 0.03 | 0.02 | 0 | -0.02 | 0.02 | 0.07 | 0.01 | 0.04 | -0.01 | 0.04 | -0.02 | -0.03 | 0.01 | -0.07 |
| *MOOD* | 0.01 | 0.05 | 0.03 | -0.03 | 0 | 0 | -0.01 | 0.04 | -0.01 | 0.02 | 0 | 0.07 | 0.02 | 0.01 | -0.02 | -0.02 | -0.02 | -0.01 | 0.02 |
| *PAR* | 0.01 | 0.03 | 0.02 | -0.04 | 0.01 | 0.02 | 0.01 | -0.01 | -0.04 | 0.03 | -0.01 | 0.01 | 0.1 | -0.01 | 0.04 | 0 | 0 | 0.02 | -0.03 |
| *CONC* | 0.06 | 0.03 | 0.02 | 0 | -0.03 | 0.01 | -0.03 | 0 | -0.02 | 0.02 | -0.02 | -0.03 | 0.01 | 0.1 | 0.01 | -0.01 | -0.01 | 0.04 | 0.02 |
| *INS* | 0.02 | 0.03 | 0.02 | 0.01 | -0.01 | -0.03 | -0.02 | 0 | -0.02 | 0.05 | 0.05 | 0 | 0.04 | 0.02 | 0.11 | 0 | 0.05 | 0.01 | -0.05 |
| *MOTIV* | 0.01 | 0.02 | 0.04 | 0.05 | -0.04 | 0.04 | 0 | -0.01 | 0 | -0.01 | -0.01 | -0.01 | -0.03 | 0.03 | 0.04 | 0.08 | 0.03 | -0.02 | 0 |
| *SUIC* | 0.01 | 0.06 | 0.03 | 0.03 | -0.01 | 0 | 0 | 0.01 | 0.04 | 0.01 | 0 | -0.04 | -0.03 | -0.01 | 0.04 | 0.01 | 0.06 | 0 | 0.02 |
| *TEAR* | 0 | 0.02 | 0.02 | 0.01 | 0 | 0.05 | -0.01 | 0.02 | 0.02 | -0.01 | -0.06 | 0.03 | -0.03 | -0.01 | 0.04 | -0.02 | 0.01 | 0.08 | 0 |
| *TOB* | 0.01 | 0.03 | 0.03 | -0.03 | -0.01 | -0.02 | 0.02 | -0.04 | -0.01 | -0.01 | -0.01 | 0.01 | 0.03 | -0.02 | -0.01 | -0.02 | 0.02 | 0 | 0.07 |

| | AGGR | AGIT | ANX | CANN | COGN | SLEEP | ELAT | HOPE | GUILT | HALL | IRR | MOOD | PAR | CONC | INS | MOTIV | SUIC | TEAR | TOB |
|---|---|---|---|---|---|---|---|---|---|---|---|---|---|---|---|---|---|---|---|
| ***AGGR*** | 0.091 (0.0012) | 0.039 (9e-04) | 0.004 (8e-04) | -0.011 (0.001) | 0.016 (8e-04) | 0.011 (9e-04) | 0.041 (9e-04) | -0.035 (9e-04) | -0.02 (9e-04) | 0.017 (0.001) | -0.01 (9e-04) | 0.007 (9e-04) | 0.009 (9e-04) | 0.024 (9e-04) | -0.009 (7e-04) | -0.003 (9e-04) | -0.022 (8e-04) | -0.014 (9e-04) | -0.06 (8e-04) |
| ***AGIT*** | 0.053 (0.001) | 0.077 (0.0011) | -0.008 (9e-04) | -0.014 (0.0011) | -0.005 (0.001) | 0.038 (9e-04) | 0.025 (9e-04) | 0.008 (9e-04) | 0.008 (9e-04) | 0.003 (8e-04) | -0.018 (0.001) | -0.004 (8e-04) | 0.027 (0.001) | -0.012 (9e-04) | 0.012 (0.001) | -0.006 (8e-04) | -0.011 (8e-04) | 0.043 (9e-04) | 0 (0) |
| ***ANX*** | 0.003 (8e-04) | 0 (0) | 0.012 (0.001) | -0.041 (9e-04) | 0.071 (9e-04) | -0.021 (8e-04) | 0.008 (8e-04) | -0.014 (8e-04) | -0.021 (7e-04) | 0.004 (8e-04) | -0.007 (8e-04) | 0.044 (7e-04) | 0.048 (8e-04) | -0.022 (8e-04) | 0.002 (7e-04) | -0.007 (7e-04) | 0.02 (7e-04) | 0.036 (9e-04) | 0.004 (9e-04) |
| ***CANN*** | 0.051 (9e-04) | 0.028 (9e-04) | 0.019 (8e-04) | 0.019 (0.0016) | -0.035 (9e-04) | 0.019 (8e-04) | -0.046 (8e-04) | 0.022 (9e-04) | 0 (0) | -0.007 (9e-04) | -0.002 (8e-04) | 0.022 (8e-04) | 0.013 (8e-04) | -0.031 (8e-04) | -0.006 (8e-04) | -0.013 (8e-04) | -0.011 (6e-04) | 0.008 (8e-04) | -0.021 (8e-04) |
| ***COGN*** | 0.009 (9e-04) | 0.023 (8e-04) | 0.031 (8e-04) | -0.018 (9e-04) | 0.021 (9e-04) | 0.009 (8e-04) | 0.013 (8e-04) | 0 (0) | 0 (0) | 0 (0) | -0.036 (8e-04) | 0.013 (8e-04) | 0.053 (8e-04) | 0.027 (8e-04) | 0.005 (8e-04) | -0.02 (8e-04) | 0.012 (8e-04) | 0.047 (8e-04) | -0.009 (8e-04) |
| ***SLEEP*** | 0.006 (9e-04) | 0.031 (9e-04) | 0.017 (9e-04) | 0 (0) | 0.003 (8e-04) | 0.07 (9e-04) | 0.012 (9e-04) | 0 (0) | -0.002 (8e-04) | 0.008 (8e-04) | -0.012 (9e-04) | 0.003 (8e-04) | 0.013 (9e-04) | 0.019 (9e-04) | -0.002 (8e-04) | 0.019 (8e-04) | 0.033 (8e-04) | 0.053 (8e-04) | -0.018 (9e-04) |
| ***ELAT*** | 0.026 (9e-04) | 0.064 (8e-04) | -0.013 (8e-04) | -0.062 (0.001) | 0.004 (8e-04) | 0.003 (8e-04) | 0.113 (0.0011) | -0.011 (9e-04) | -0.039 (8e-04) | 0.012 (0.001) | 0.041 (9e-04) | -0.023 (8e-04) | 0.077 (9e-04) | 0.027 (8e-04) | -0.02 (8e-04) | -0.014 (8e-04) | -0.003 (8e-04) | 0.015 (9e-04) | 0.002 (9e-04) |
| ***HOPE*** | -0.02 (8e-04) | 0.05 (0.001) | 0 (0) | -0.003 (9e-04) | -0.045 (9e-04) | 0.027 (9e-04) | -0.056 (7e-04) | 0.113 (0.0015) | -0.013 (0.001) | 0 (0) | -0.003 (0.001) | 0.002 (8e-04) | -0.011 (9e-04) | 0.025 (9e-04) | 0.037 (9e-04) | -0.012 (9e-04) | 0 (0) | 0.031 (0.001) | 0.014 (0.001) |
| ***GUILT*** | -0.014 (9e-04) | -0.005 (9e-04) | 0 (8e-04) | 0.008 (0.001) | -0.013 (9e-04) | 0.01 (9e-04) | 0.01 (9e-04) | 0 (0) | 0.038 (0.0013) | 0 (0.001) | 0.005 (9e-04) | 0.017 (9e-04) | -0.03 (9e-04) | -0.005 (0.001) | -0.002 (8e-04) | 0.032 (8e-04) | 0 (9e-04) | -0.008 (0.001) | 0.018 (0.001) |
| ***HALL*** | 0.019 (9e-04) | -0.01 (9e-04) | -0.016 (9e-04) | -0.009 (0.0011) | 0.002 (9e-04) | -0.016 (9e-04) | 0.012 (8e-04) | 0 (0.001) | -0.022 (8e-04) | 0.075 (0.0016) | -0.008 (0.001) | -0.022 (8e-04) | 0.1 (9e-04) | 0.03 (0.0011) | 0.021 (8e-04) | 0.011 (8e-04) | 0.02 (8e-04) | -0.025 (8e-04) | -0.023 (9e-04) |
| ***IRR*** | 0.028 (9e-04) | 0 (8e-04) | 0.006 (8e-04) | -0.044 (0.001) | 0 (0) | 0.025 (9e-04) | 0.025 (9e-04) | 0 (0) | -0.017 (9e-04) | 0.017 (9e-04) | 0.066 (0.0011) | 0 (0) | 0.041 (9e-04) | -0.014 (9e-04) | 0.042 (7e-04) | -0.018 (8e-04) | -0.029 (8e-04) | 0.009 (9e-04) | -0.073 (9e-04) |
| ***MOOD*** | 0.012 (9e-04) | 0 (0) | 0.03 (8e-04) | -0.031 (8e-04) | 0.002 (9e-04) | 0 (0.001) | -0.009 (9e-04) | 0.038 (9e-04) | -0.01 (9e-04) | 0.021 (9e-04) | -0.002 (0.001) | 0.068 (0.001) | 0.021 (0.001) | 0.01 (9e-04) | -0.023 (8e-04) | -0.016 (7e-04) | -0.022 (8e-04) | -0.006 (9e-04) | 0.021 (8e-04) |
| ***PAR*** | 0.009 (9e-04) | 0.027 (8e-04) | 0.014 (8e-04) | -0.04 (9e-04) | 0.011 (8e-04) | 0 (0) | 0.003 (0.001) | -0.011 (9e-04) | -0.046 (9e-04) | 0.028 (9e-04) | -0.014 (9e-04) | 0.01 (8e-04) | 0.1 (0.0011) | -0.008 (8e-04) | 0.032 (7e-04) | -0.004 (7e-04) | -0.002 (8e-04) | 0.023 (9e-04) | -0.032 (0.001) |
| ***CONC*** | 0.064 (9e-04) | 0.026 (9e-04) | 0.025 (8e-04) | 0 (0.0011) | -0.029 (9e-04) | 0.005 (9e-04) | -0.031 (9e-04) | 0 (0.001) | 0 (0) | 0.012 (0.001) | -0.019 (9e-04) | -0.032 (0.001) | 0.007 (0.001) | 0.1 (0.0011) | 0.009 (8e-04) | -0.013 (9e-04) | -0.014 (8e-04) | 0.035 (9e-04) | 0 (0) |
| ***INS*** | 0.021 (0.001) | 0.029 (9e-04) | 0.016 (8e-04) | 0.008 (0.0012) | 0 (9e-04) | -0.035 (0.001) | -0.017 (9e-04) | 0 (9e-04) | -0.024 (9e-04) | 0.047 (0.001) | 0.046 (9e-04) | 0 (9e-04) | 0.037 (0.001) | 0.017 (8e-04) | 0.108 (0.001) | -0.002 (8e-04) | 0.049 (9e-04) | 0.013 (9e-04) | -0.054 (8e-04) |
| ***MOTIV*** | 0.008 (9e-04) | 0.016 (9e-04) | 0.037 (8e-04) | 0.045 (0.0011) | -0.04 (9e-04) | 0.041 (0.001) | 0.003 (0.001) | -0.006 (0.001) | 0 (9e-04) | -0.012 (9e-04) | -0.011 (9e-04) | -0.006 (9e-04) | -0.032 (9e-04) | 0.034 (9e-04) | 0.038 (9e-04) | 0.072 (0.0012) | 0.028 (9e-04) | -0.025 (0.001) | 0 (9e-04) |
| ***SUIC*** | 0.01 (0.001) | 0.056 (9e-04) | 0.029 (8e-04) | 0.032 (0.0011) | -0.008 (8e-04) | 0 (0) | -0.004 (8e-04) | 0.002 (0.0011) | 0.04 (9e-04) | 0.006 (9e-04) | -0.003 (0.001) | -0.04 (9e-04) | -0.025 (9e-04) | -0.008 (0.001) | 0.036 (8e-04) | 0.014 (9e-04) | 0.058 (0.0012) | 0.005 (0.001) | 0.02 (9e-04) |
| ***TEAR*** | -0.002 (8e-04) | 0.025 (9e-04) | 0.02 (8e-04) | 0.009 (9e-04) | 0 (0) | 0.049 (0.001) | -0.014 (7e-04) | 0.019 (0.001) | 0.019 (9e-04) | -0.006 (8e-04) | -0.056 (0.001) | 0.026 (9e-04) | -0.029 (8e-04) | -0.009 (9e-04) | 0.039 (8e-04) | -0.022 (8e-04) | 0.005 (9e-04) | 0.079 (0.0012) | 0.003 (8e-04) |
| ***TOB*** | 0.017 (9e-04) | 0.028 (8e-04) | 0.027 (8e-04) | -0.032 (0.0011) | -0.011 (8e-04) | -0.022 (9e-04) | 0.014 (8e-04) | -0.039 (8e-04) | -0.01 (9e-04) | -0.003 (0.001) | -0.01 (9e-04) | 0.012 (9e-04) | 0.025 (9e-04) | -0.015 (8e-04) | -0.006 (8e-04) | -0.016 (9e-04) | 0.021 (8e-04) | 0 (0) | 0.069 (0.0012) |

### c. PSY

| | AGGR | AGIT | ANX | AROUS | CANN | COC | COG | DEL | SLEEP | ELAT | EMOT | HOPE | HALL | HOST | IRR | MOOD | PAR | CONC | INS | MOTIV | SUIC | TEAR | TOB | WGHT |
|---|---|---|---|---|---|---|---|---|---|---|---|---|---|---|---|---|---|---|---|---|---|---|---|---|
| ***AGGR*** | 0.06 | 0.04 | -0.02 | 0.02 | -0.03 | -0.01 | 0.01 | 0.01 | -0.01 | 0.01 | 0.02 | -0.02 | -0.01 | 0.03 | 0 | -0.02 | 0.02 | -0.01 | -0.01 | -0.01 | -0.01 | 0.02 | 0.01 | 0 |
| ***AGIT*** | 0.04 | 0.07 | 0 | 0.04 | -0.02 | -0.02 | 0 | -0.01 | -0.02 | 0.02 | 0 | -0.01 | 0.02 | -0.01 | 0.02 | 0 | 0 | 0 | 0 | -0.01 | 0 | 0.01 | 0.02 | 0 |
| ***ANX*** | 0.01 | 0 | 0.06 | 0.02 | -0.01 | -0.01 | 0 | 0.02 | 0.01 | 0 | 0.01 | 0 | 0.01 | -0.01 | -0.01 | -0.01 | 0.01 | 0 | 0 | 0 | -0.01 | 0.02 | 0.01 | 0 |
| ***AROUS*** | 0.01 | 0.02 | -0.02 | 0.09 | 0 | -0.02 | 0.02 | -0.01 | -0.02 | 0.03 | -0.01 | -0.01 | 0.03 | 0.01 | 0.03 | -0.01 | -0.01 | 0 | 0 | -0.02 | -0.01 | 0 | 0.01 | -0.01 |
| ***CANN*** | 0.01 | 0.02 | -0.01 | 0.01 | 0.04 | 0.02 | -0.01 | 0 | 0 | 0.01 | 0.01 | 0 | 0 | 0 | -0.01 | 0.01 | 0.01 | -0.01 | -0.02 | 0 | -0.01 | 0 | 0.01 | 0 |
| ***COC*** | 0.01 | 0.01 | -0.02 | 0 | -0.01 | 0.09 | 0.01 | 0 | 0 | 0.02 | 0.01 | -0.01 | 0.01 | 0.01 | 0.02 | -0.02 | -0.01 | 0 | -0.02 | 0 | 0 | 0.01 | 0.01 | 0 |
| ***COG*** | 0 | 0.01 | 0 | 0.02 | -0.01 | 0.03 | 0.05 | 0.02 | 0.02 | 0 | 0 | 0 | 0 | 0 | 0 | 0 | 0.01 | 0 | 0 | -0.01 | 0 | 0.03 | 0 | 0 |
| ***DEL*** | 0.01 | 0.02 | -0.01 | 0.02 | -0.01 | -0.01 | 0.02 | 0.07 | -0.03 | 0.03 | 0.02 | -0.01 | 0.02 | 0.01 | -0.01 | -0.02 | 0.03 | 0 | -0.01 | 0 | 0 | 0 | 0 | 0.01 |
| ***SLEEP*** | 0.02 | -0.01 | 0 | 0.01 | 0.01 | -0.03 | -0.01 | -0.01 | 0.06 | 0.03 | 0.02 | 0 | 0.01 | 0 | -0.01 | 0.01 | 0.01 | 0.01 | 0 | -0.01 | -0.02 | 0.01 | 0.01 | -0.01 |
| ***ELAT*** | 0 | 0.01 | -0.02 | 0.04 | -0.02 | -0.01 | 0.01 | 0 | 0.01 | 0.12 | 0 | 0.01 | 0.01 | 0 | 0.01 | -0.01 | -0.01 | 0 | -0.01 | -0.02 | 0.01 | 0 | -0.01 | 0 |
| ***EMOT*** | -0.01 | -0.01 | 0 | 0.01 | 0 | 0 | -0.02 | -0.01 | 0 | -0.01 | 0.04 | -0.01 | 0 | 0.02 | 0.02 | 0 | 0.03 | 0 | 0 | -0.01 | 0 | 0.03 | 0 | 0.02 |
| ***HOPE*** | 0.01 | 0 | 0 | -0.02 | -0.01 | -0.02 | -0.01 | 0 | -0.01 | 0.03 | 0.02 | 0.08 | 0 | -0.01 | 0 | -0.01 | 0 | 0.02 | 0.02 | -0.01 | 0.02 | 0.03 | -0.01 | 0 |
| ***HALL*** | 0 | 0.01 | 0 | 0.01 | -0.02 | 0 | -0.01 | 0.02 | 0.02 | 0.02 | 0 | -0.01 | 0.08 | 0 | -0.02 | -0.02 | 0.03 | 0 | -0.02 | 0 | 0.01 | 0.02 | 0 | 0 |
| ***HOST*** | 0.02 | 0 | 0 | 0.03 | -0.01 | 0 | 0 | 0.02 | -0.02 | 0.03 | 0 | -0.01 | 0 | 0.07 | 0.02 | 0.01 | 0 | -0.01 | -0.01 | 0 | 0 | 0.01 | 0 | 0 |
| ***IRR*** | 0.01 | 0.03 | -0.02 | 0.03 | -0.01 | 0 | 0.02 | 0 | 0 | 0.02 | 0.01 | -0.02 | 0 | 0.01 | 0.07 | 0 | 0 | -0.01 | -0.02 | -0.01 | 0.01 | 0.01 | 0.02 | 0.01 |
| ***MOOD*** | -0.01 | 0.02 | -0.03 | 0.01 | 0 | -0.03 | 0.01 | 0 | 0 | 0.01 | 0 | 0 | -0.01 | 0.01 | 0.03 | 0.06 | 0.02 | 0.01 | -0.01 | 0 | 0.01 | 0.02 | -0.01 | 0 |
| ***PAR*** | 0.01 | 0.01 | -0.01 | 0.01 | -0.01 | -0.01 | 0.02 | 0.02 | 0.01 | -0.01 | 0.02 | -0.02 | 0.01 | 0 | 0 | 0 | 0.1 | 0.01 | -0.01 | -0.01 | 0 | 0.01 | 0 | 0 |
| ***CONC*** | 0.01 | 0.02 | -0.01 | 0.01 | 0 | -0.02 | 0 | -0.02 | 0.01 | 0 | -0.01 | 0.01 | 0 | -0.01 | 0 | 0.01 | 0.02 | 0.04 | 0 | -0.01 | 0.01 | 0.03 | 0 | 0 |
| ***INS*** | 0.02 | -0.01 | -0.02 | 0.01 | -0.03 | -0.01 | -0.01 | 0.01 | 0 | 0.02 | 0.01 | -0.01 | 0 | 0.01 | 0 | 0 | 0.02 | 0 | 0.05 | -0.01 | 0.03 | 0.03 | 0.01 | -0.01 |
| ***MOTIV*** | -0.01 | 0 | 0.01 | 0 | 0 | -0.01 | 0.01 | 0 | 0 | -0.01 | 0 | -0.01 | 0.02 | -0.01 | 0.02 | 0.01 | 0 | 0 | 0.01 | 0.06 | -0.03 | 0.01 | 0 | -0.01 |
| ***SUIC*** | 0 | 0 | -0.01 | 0 | -0.01 | -0.02 | -0.01 | 0 | -0.02 | 0 | 0 | 0.02 | 0 | 0 | 0.02 | -0.01 | 0 | -0.01 | 0 | 0.01 | 0.09 | 0.02 | 0 | 0 |
| ***TEAR*** | 0 | 0.01 | 0.01 | 0.01 | -0.02 | -0.01 | -0.01 | 0.02 | -0.02 | 0.01 | 0 | 0.01 | -0.01 | -0.01 | 0 | 0.01 | 0.01 | 0.01 | 0.01 | -0.03 | -0.01 | 0.13 | -0.02 | -0.01 |
| ***TOB*** | 0.01 | 0 | 0.01 | 0.01 | 0.01 | -0.01 | 0 | 0.01 | 0 | 0.01 | 0.01 | -0.02 | 0 | 0.01 | -0.01 | 0 | 0 | 0 | -0.02 | -0.01 | 0.01 | 0.01 | 0.08 | 0 |
| ***WGHT*** | 0 | 0 | -0.01 | 0 | -0.01 | -0.01 | -0.02 | 0 | 0.02 | 0 | 0.01 | -0.01 | -0.01 | -0.01 | -0.01 | 0.01 | 0.01 | -0.01 | -0.01 | 0.01 | 0.03 | 0 | -0.02 | 0.08 |

| | AGGR | AGIT | ANX | AROUS | CANN | COC | COG | DEL | SLEEP | ELAT | EMOT | HOPE | HALL | HOST | IRR | MOOD | PAR | CONC | INS | MOTIV | SUIC | TEAR | TOB | WGHT |
|---|---|---|---|---|---|---|---|---|---|---|---|---|---|---|---|---|---|---|---|---|---|---|---|---|
| ***AGGR*** | 0.033 (0.008) | -0.017 (0.0072) | -0.073 (0.0056) | 0.032 (0.0089) | -0.058 (0.0044) | -0.031 (0.0036) | -0.061 (0.0064) | -0.038 (0.0061) | -0.069 (0.0058) | 0 (0) | -0.015 (0.0041) | -0.069 (0.006) | -0.063 (0.0057) | 0.017 (0.0066) | -0.035 (0.0064) | -0.055 (0.0043) | -0.041 (0.0065) | -0.068 (0.0062) | -0.067 (0.006) | -0.062 (0.007) | -0.039 (0.0035) | 0 (0) | -0.029 (0.0047) | -0.031 (0.0037) |
| ***AGIT*** | 0 (0) | 0.034 (0.0091) | -0.059 (0.0058) | 0.036 (0.0095) | -0.052 (0.0049) | -0.041 (0.0039) | -0.068 (0.0068) | -0.057 (0.0066) | -0.08 (0.0062) | 0 (0) | -0.034 (0.0041) | -0.058 (0.0065) | -0.046 (0.0063) | -0.017 (0.0073) | -0.023 (0.0068) | -0.047 (0.0046) | -0.064 (0.007) | -0.065 (0.0068) | -0.057 (0.0062) | -0.064 (0.0074) | -0.046 (0.0049) | -0.025 (0.0051) | -0.021 (0.005) | -0.027 (0.0038) |
| ***ANX*** | 0 (0) | -0.034 (0.0057) | 0.024 (0.0048) | 0.035 (0.007) | -0.038 (0.0036) | -0.026 (0.0029) | -0.063 (0.0053) | -0.024 (0.0049) | -0.04 (0.0045) | 0 (0) | -0.014 (0.0026) | -0.034 (0.0048) | -0.036 (0.0047) | 0 (0) | -0.026 (0.0048) | -0.029 (0.0032) | -0.038 (0.0052) | -0.048 (0.0049) | -0.046 (0.0048) | -0.044 (0.0057) | -0.027 (0.0027) | 0 (0) | -0.022 (0.0035) | -0.017 (0.0026) |
| ***AROUS*** | -0.022 (0.0045) | -0.026 (0.0057) | -0.055 (0.0038) | 0.094 (0.0069) | -0.029 (0.0031) | -0.034 (0.0025) | -0.029 (0.0042) | -0.047 (0.0041) | -0.055 (0.0037) | 0.014 (0.0043) | -0.027 (0.0025) | -0.046 (0.0044) | -0.021 (0.004) | 0 (0) | 0 (0) | -0.04 (0.0028) | -0.049 (0.0042) | -0.047 (0.004) | -0.044 (0.004) | -0.058 (0.0054) | -0.023 (0.0022) | -0.02 (0.003) | -0.018 (0.003) | -0.023 (0.0024) |
| ***CANN*** | 0 (0) | 0 (0) | -0.063 (0.0052) | 0.025 (0.0087) | 0.037 (0.0049) | -0.015 (0.0032) | -0.068 (0.0059) | -0.044 (0.0054) | -0.052 (0.0052) | 0 (0) | -0.022 (0.0031) | -0.051 (0.0054) | -0.046 (0.005) | 0 (0) | -0.027 (0.0057) | -0.025 (0.004) | -0.047 (0.0056) | -0.066 (0.0059) | -0.078 (0.006) | -0.055 (0.0064) | -0.043 (0.0034) | -0.016 (0.0041) | -0.035 (0.0043) | -0.033 (0.0034) |
| ***COC*** | -0.014 (0.0045) | -0.012 (0.0053) | -0.054 (0.004) | 0.019 (0.0059) | -0.048 (0.003) | 0.085 (0.0045) | -0.04 (0.0043) | -0.033 (0.0039) | -0.036 (0.0039) | 0.018 (0.0041) | -0.014 (0.0024) | -0.046 (0.0042) | -0.031 (0.0038) | 0 (0) | 0 (0) | -0.038 (0.003) | -0.041 (0.004) | -0.041 (0.0043) | -0.062 (0.0046) | -0.038 (0.0047) | -0.027 (0.0026) | 0 (0) | -0.015 (0.0031) | -0.019 (0.0024) |
| ***COG*** | -0.033 (0.0069) | -0.026 (0.0068) | -0.062 (0.0054) | 0.029 (0.0079) | -0.036 (0.0042) | 0 (0) | 0 (0) | -0.03 (0.0058) | -0.035 (0.0052) | 0 (0) | -0.025 (0.0032) | -0.046 (0.0056) | -0.047 (0.0054) | 0 (0) | -0.029 (0.0058) | -0.03 (0.0039) | -0.05 (0.0061) | -0.066 (0.006) | -0.054 (0.0056) | -0.065 (0.0067) | -0.022 (0.0032) | 0 (0) | -0.036 (0.0043) | -0.019 (0.0033) |
| ***DEL*** | -0.025 (0.0074) | 0 (0) | -0.069 (0.0059) | 0.025 (0.0094) | -0.044 (0.0046) | -0.028 (0.0037) | -0.057 (0.0068) | 0.032 (0.0077) | -0.086 (0.0063) | 0 (0) | 0 (0.0037) | -0.064 (0.0063) | -0.05 (0.0061) | 0 (0) | -0.04 (0.0067) | -0.064 (0.0048) | -0.045 (0.0069) | 0 (0) | -0.072 (0.0064) | -0.053 (0.0072) | -0.036 (0.004) | -0.029 (0.0049) | -0.031 (0.0049) | -0.028 (0.0038) |
| ***SLEEP*** | 0 (0.0058) | -0.041 (0.0061) | -0.055 (0.0046) | 0.023 (0.0072) | -0.022 (0.0039) | -0.037 (0.0031) | -0.069 (0.0054) | -0.04 (0.0051) | 0.014 (0.005) | 0.016 (0.005) | 0 (0) | -0.043 (0.005) | -0.05 (0.0049) | 0 (0) | -0.037 (0.005) | -0.026 (0.0034) | -0.042 (0.0053) | -0.051 (0.0051) | -0.053 (0.0049) | -0.054 (0.0059) | -0.039 (0.0028) | -0.018 (0.0034) | -0.022 (0.0037) | -0.034 (0.003) |
| ***ELAT*** | -0.022 (0.0044) | -0.013 (0.0045) | -0.053 (0.0037) | 0.043 (0.0061) | -0.036 (0.0029) | -0.024 (0.0023) | -0.034 (0.0042) | -0.035 (0.0039) | -0.035 (0.0037) | 0.106 (0.0049) | -0.023 (0.0023) | -0.03 (0.0041) | -0.031 (0.0037) | -0.008 (0.0041) | -0.012 (0.0039) | -0.032 (0.0028) | -0.043 (0.004) | -0.041 (0.004) | -0.052 (0.004) | -0.053 (0.005) | -0.005 (0.0021) | -0.018 (0.0029) | -0.032 (0.003) | -0.021 (0.0024) |
| ***EMOT*** | -0.036 (0.0033) | -0.03 (0.0033) | -0.027 (0.0026) | 0.017 (0.0039) | -0.022 (0.0022) | -0.019 (0.002) | -0.045 (0.003) | -0.037 (0.0029) | -0.03 (0.0027) | -0.011 (0.0026) | 0.02 (0.002) | -0.036 (0.0032) | -0.035 (0.0027) | 0.008 (0.003) | -0.006 (0.0029) | -0.016 (0.0018) | -0.015 (0.0031) | -0.029 (0.0029) | -0.036 (0.0029) | -0.035 (0.0036) | -0.019 (0.0016) | 0.012 (0.0019) | -0.021 (0.0021) | 0 (0) |
| ***HOPE*** | 0 (0.0031) | -0.016 (0.0029) | -0.025 (0.0026) | 0 (0.0035) | -0.028 (0.0021) | -0.024 (0.0016) | -0.037 (0.0027) | -0.022 (0.0025) | -0.028 (0.0024) | 0.026 (0.0027) | 0.006 (0.0016) | 0.053 (0.0031) | -0.024 (0.0023) | 0 (0) | -0.006 (0.0024) | -0.027 (0.0019) | -0.018 (0.0025) | -0.014 (0.0027) | -0.013 (0.0026) | -0.027 (0.0036) | -0.004 (0.0017) | 0 (0) | -0.018 (0.0017) | -0.009 (0.0017) |
| ***HALL)*** | 0 (0) | -0.034 (0.0071) | -0.05 (0.0055) | 0 (0) | -0.046 (0.0044) | -0.03 (0.0037) | -0.068 (0.0064) | -0.039 (0.0062) | -0.037 (0.0057) | 0.017 (0.0062) | -0.032 (0.0034) | -0.062 (0.0059) | 0.04 (0.0067) | 0 (0.0065) | -0.042 (0.0061) | -0.051 (0.0043) | -0.035 (0.0064) | -0.061 (0.0062) | -0.076 (0.0063) | -0.055 (0.0067) | -0.031 (0.0036) | 0 (0.0044) | -0.038 (0.0046) | -0.027 (0.0035) |
| ***HOST*** | -0.018 (0.0048) | -0.028 (0.005) | -0.039 (0.0038) | 0.028 (0.0064) | -0.03 (0.0032) | -0.011 (0.0025) | -0.046 (0.0045) | -0.018 (0.0043) | -0.061 (0.0042) | 0.027 (0.0045) | -0.027 (0.0025) | -0.047 (0.0046) | -0.043 (0.0039) | 0.072 (0.0051) | -0.015 (0.0044) | -0.017 (0.0028) | -0.038 (0.0044) | -0.05 (0.0044) | -0.052 (0.0044) | -0.03 (0.0052) | -0.029 (0.0023) | -0.011 (0.0029) | -0.022 (0.0031) | -0.021 (0.0026) |
| ***IRR*** | 0 (0) | -0.016 (0.0065) | -0.061 (0.0049) | 0.031 (0.0079) | -0.037 (0.004) | -0.026 (0.0032) | -0.04 (0.0056) | -0.036 (0.0053) | -0.054 (0.0052) | 0 (0.0054) | -0.02 (0.0031) | -0.064 (0.0055) | -0.047 (0.0052) | 0 (0.0056) | 0.055 (0.0065) | -0.04 (0.0039) | -0.048 (0.0057) | -0.068 (0.0057) | -0.072 (0.0055) | -0.05 (0.0063) | 0 (0) | -0.018 (0.0039) | -0.017 (0.004) | -0.02 (0.0031) |
| ***MOOD*** | -0.025 (0.0044) | 0 (0.0048) | -0.059 (0.0038) | 0.017 (0.0057) | -0.032 (0.0032) | -0.04 (0.0024) | -0.037 (0.0041) | -0.026 (0.0041) | -0.037 (0.0037) | 0 (0) | -0.02 (0.0023) | -0.032 (0.0043) | -0.04 (0.0039) | 0 (0) | 0 (0.0042) | 0.031 (0.0031) | -0.026 (0.0042) | -0.033 (0.0042) | -0.045 (0.0042) | -0.032 (0.0049) | -0.008 (0.002) | -0.007 (0.0027) | -0.026 (0.0028) | -0.026 (0.0025) |
| ***PAR*** | -0.025 (0.0077) | 0 (0) | -0.069 (0.0064) | 0.025 (0.01) | -0.048 (0.0048) | -0.033 (0.0038) | -0.055 (0.007) | -0.034 (0.0068) | -0.053 (0.0066) | 0 (0) | -0.017 (0.0043) | -0.08 (0.0068) | -0.055 (0.0064) | 0 (0) | 0 (0) | -0.04 (0.0053) | 0.044 (0.0085) | -0.054 (0.007) | -0.074 (0.007) | -0.079 (0.0081) | -0.037 (0.0047) | -0.019 (0.0055) | -0.041 (0.0053) | -0.035 (0.0044) |
| ***CONC*** | -0.01 (0.0049) | 0 (0.0051) | -0.042 (0.0037) | 0.018 (0.0059) | -0.026 (0.0033) | -0.033 (0.0027) | -0.052 (0.0044) | -0.048 (0.0042) | -0.031 (0.0038) | 0 (0.004) | -0.025 (0.0027) | -0.033 (0.0045) | -0.044 (0.0039) | 0 (0.0043) | -0.015 (0.0042) | -0.025 (0.0028) | -0.022 (0.0043) | 0 (0.0042) | -0.041 (0.0041) | -0.046 (0.0055) | -0.012 (0.0023) | 0.009 (0.0028) | -0.023 (0.003) | -0.023 (0.0025) |
| ***INS*** | 0 (0.0048) | -0.039 (0.005) | -0.059 (0.0042) | 0.02 (0.0063) | -0.049 (0.0034) | -0.025 (0.0028) | -0.054 (0.0046) | -0.03 (0.0044) | -0.046 (0.0042) | 0 (0) | -0.014 (0.0025) | -0.042 (0.0048) | -0.044 (0.0042) | 0 (0) | -0.012 (0.0044) | -0.018 (0.0028) | -0.033 (0.0047) | -0.041 (0.0046) | 0.011 (0.0046) | -0.042 (0.0054) | 0 (0.0023) | 0.009 (0.0029) | -0.016 (0.0031) | -0.019 (0.0026) |
| ***MOTIV*** | 0 (0) | -0.016 (0.0026) | -0.013 (0.002) | 0.01 (0.0032) | -0.019 (0.0018) | -0.017 (0.0014) | -0.015 (0.0023) | -0.02 (0.0022) | -0.022 (0.002) | -0.008 (0.002) | -0.007 (0.0019) | -0.027 (0.003) | -0.007 (0.0021) | 0 (0) | 0.005 (0.0022) | -0.009 (0.0016) | -0.016 (0.0022) | -0.026 (0.0026) | -0.008 (0.0023) | 0.037 (0.0033) | -0.043 (0.0019) | 0.003 (0.0016) | -0.014 (0.0017) | -0.02 (0.0018) |

| | | | | | | | | | | | | | | | | | | | | | | | | |
|---|---|---|---|---|---|---|---|---|---|---|---|---|---|---|---|---|---|---|---|---|---|---|---|---|
| ***SUIC*** | -0.019 (0.0039) | -0.031 (0.0041) | -0.037 (0.0031) | 0 (0) | -0.032 (0.0027) | -0.028 (0.002) | -0.053 (0.0036) | -0.032 (0.0034) | -0.053 (0.0033) | -0.007 (0.0033) | -0.015 (0.002) | -0.016 (0.0035) | -0.035 (0.0033) | -0.008 (0.0037) | 0 (0) | -0.033 (0.0025) | -0.034 (0.0035) | -0.05 (0.0037) | -0.04 (0.0035) | -0.019 (0.004) | 0.062 (0.0027) | 0 (0) | -0.025 (0.0025) | -0.023 (0.0022) |
| ***TEAR*** | -0.025 (0.0042) | -0.022 (0.0046) | -0.034 (0.0034) | 0.014 (0.0055) | -0.04 (0.0028) | -0.023 (0.0023) | -0.053 (0.0041) | -0.017 (0.004) | -0.057 (0.0037) | 0 (0) | -0.022 (0.0023) | -0.036 (0.0039) | -0.042 (0.0037) | -0.017 (0.004) | -0.02 (0.0039) | -0.023 (0.0027) | -0.034 (0.0042) | -0.038 (0.004) | -0.039 (0.0038) | -0.062 (0.0049) | -0.036 (0.0023) | 0.101 (0.004) | -0.047 (0.003) | -0.031 (0.0023) |
| ***TOB*** | -0.022 (0.0051) | -0.029 (0.0052) | -0.028 (0.004) | 0.02 (0.0064) | -0.022 (0.0034) | -0.023 (0.0027) | -0.052 (0.0048) | -0.029 (0.0045) | -0.044 (0.0042) | 0 (0) | -0.018 (0.0026) | -0.052 (0.0048) | -0.038 (0.0042) | 0 (0) | -0.029 (0.0045) | -0.029 (0.0029) | -0.045 (0.0046) | -0.052 (0.0047) | -0.062 (0.0046) | -0.051 (0.0056) | -0.02 (0.0025) | 0 (0) | 0.059 (0.004) | -0.018 (0.0026) |
| ***WGHT*** | -0.026 (0.0036) | -0.02 (0.0031) | -0.039 (0.0024) | 0 (0) | -0.028 (0.0019) | -0.02 (0.0015) | -0.048 (0.0027) | -0.034 (0.0026) | -0.016 (0.0023) | -0.008 (0.0025) | -0.015 (0.0017) | -0.036 (0.0027) | -0.04 (0.0024) | -0.011 (0.0027) | -0.031 (0.0025) | 0 (0) | -0.028 (0.0026) | -0.037 (0.0025) | -0.045 (0.0027) | -0.011 (0.0028) | 0.006 (0.0014) | -0.011 (0.0019) | -0.035 (0.002) | 0.052 (0.0018) |

eFigure 4 Top 24 temporal sub-network edge estimates (bootstrapped [black] vs actual [red]) in sub-networks: (A) UMD, (B) BMD and (C) PSY. For each bootstrap, 75% of the sample was randomly drawn without replacement and missing data imputed before being modelled in the same manner as the full data approximations.

**(A) UMD** **(B) BMD** **(C) PSY**

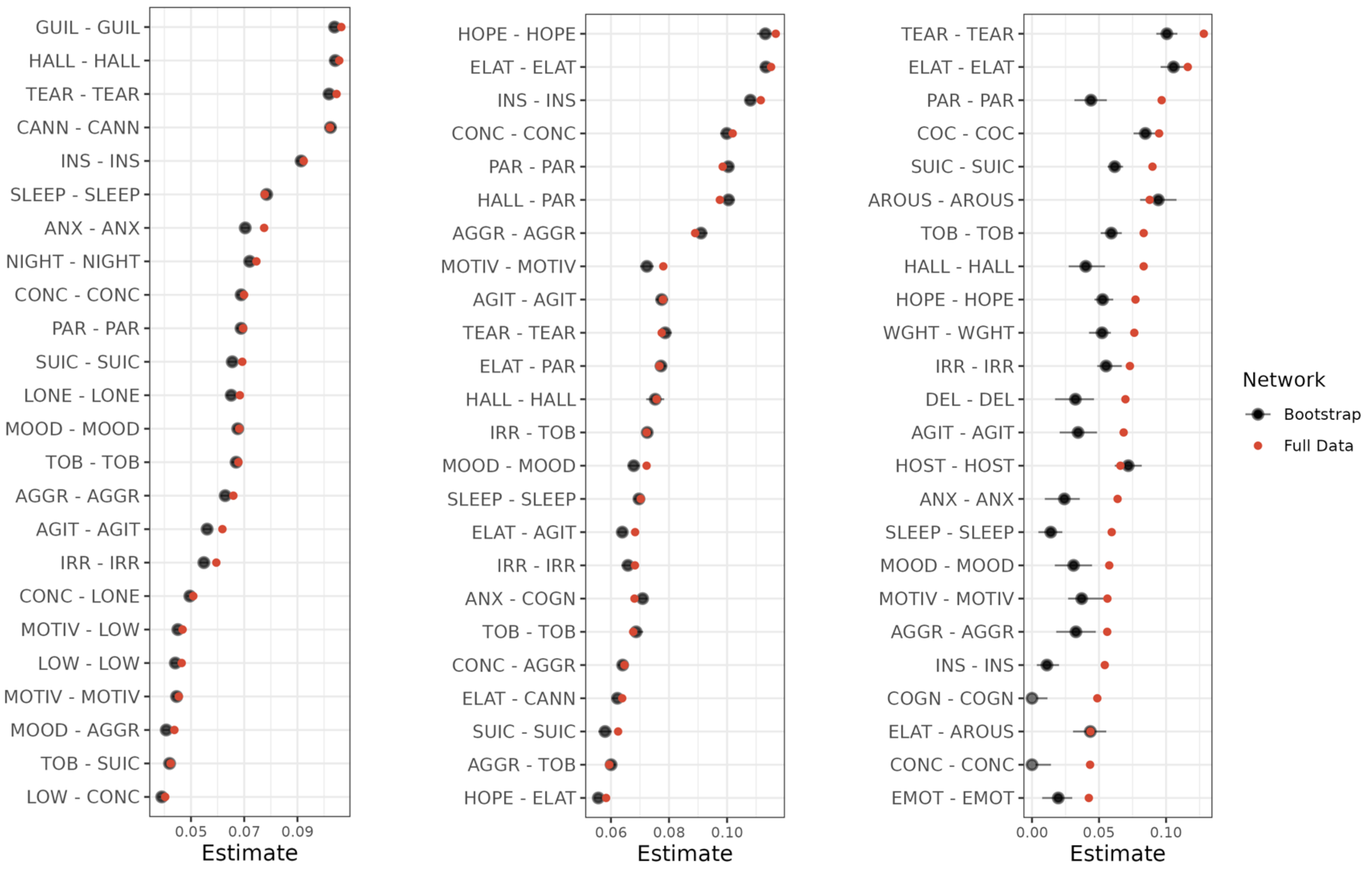
GUIL - GUIL
HALL - HALL
TEAR - TEAR
CANN - CANN
INS - INS
SLEEP - SLEEP
ANX - ANX
NIGHT - NIGHT
CONC - CONC
PAR - PAR
SUIC - SUIC
LONE - LONE
MOOD - MOOD
TOB - TOB
AGGR - AGGR
AGIT - AGIT
IRR - IRR
CONC - LONE
MOTIV - LOW
LOW - LOW
MOTIV - MOTIV
MOOD - AGGR
TOB - SUIC
LOW - CONC
0.05
0.07
0.09
Estimate
HOPE - HOPE
ELAT - ELAT
INS - INS
CONC - CONC
PAR - PAR
HALL - PAR
AGGR - AGGR
MOTIV - MOTIV
AGIT - AGIT
TEAR - TEAR
ELAT - PAR
HALL - HALL
IRR - TOB
MOOD - MOOD
SLEEP - SLEEP
ELAT - AGIT
IRR - IRR
ANX - COGN
TOB - TOB
CONC - AGGR
ELAT - CANN
SUIC - SUIC
AGGR - TOB
HOPE - ELAT
0.06
0.08
0.10
Estimate
TEAR - TEAR
ELAT - ELAT
PAR - PAR
COC - COC
SUIC - SUIC
AROUS - AROUS
TOB - TOB
HALL - HALL
HOPE - HOPE
WGHT - WGHT
IRR - IRR
DEL - DEL
AGIT - AGIT
HOST - HOST
ANX - ANX
SLEEP - SLEEP
MOOD - MOOD
MOTIV - MOTIV
AGGR - AGGR
INS - INS
COGN - COGN
ELAT - AROUS
CONC - CONC
EMOT - EMOT
0.00
0.05
0.10
Estimate
Network
Bootstrap
Full Data

eTable 12 Contemporaneous and between-subject edge estimates for sub-networks: (A) UMD, (B) BMD and (C) PSY. For each sub-network, (i) contemporaneous sub-network edge estimates and (ii) between-subject sub-network edge estimates (actual model in green and bootstrapped estimates in blue). Any bootstrapped edges that had 95% CI crossing zero were forced to 0.

### (a) UMD

#### i. Contemporaneous

| | AGGR | AGIT | ANX | CANN | COG | SLEEP | LONE | GUILT | HALLL | IRR | ENERGY | MOOD | NIGHT | PAR | CONC | INS | MOTIV | SUIC | TEAR | TOB | WGHT |
|---|---|---|---|---|---|---|---|---|---|---|---|---|---|---|---|---|---|---|---|---|---|
| ***AGGR*** | NA | 0.087 (0.0021) | 0.034 (0.0026) | 0.026 (0.0018) | 0.059 (0.0031) | 0.025 (0.003) | 0.014 (0.0013) | 0.017 (0.002) | 0.038 (0.0019) | 0.081 (0.0018) | 0.016 (0.0021) | 0.04 (0.0019) | 0.006 (0.0016) | 0.048 (0.0022) | 0 (0) | 0.053 (0.0023) | 0 (0) | 0.036 (0.0023) | 0 (0) | 0.04 (0.0017) | 0.02 (0.0016) |
| ***AGIT*** | 0.08 | NA | 0.035 (0.003) | 0.011 (0.0022) | 0.021 (0.0039) | 0.068 (0.0035) | 0.02 (0.0015) | 0.023 (0.0023) | 0.033 (0.0022) | 0.063 (0.0022) | 0 (0) | 0.033 (0.0023) | 0.023 (0.0019) | 0.074 (0.0025) | 0.039 (0.0036) | 0.05 (0.0027) | 0.02 (0.0022) | 0.048 (0.0026) | 0.049 (0.003) | 0.043 (0.002) | 0.024 (0.0018) |
| ***ANX*** | 0.03 | 0.03 | NA | 0.013 (0.0025) | 0.195 (0.0041) | 0.098 (0.0042) | 0.019 (0.0017) | 0.041 (0.0028) | 0.007 (0.0027) | 0.013 (0.0027) | 0 (0) | 0.021 (0.0028) | 0.048 (0.0022) | 0.048 (0.003) | 0.022 (0.0046) | 0.051 (0.0032) | 0.017 (0.0027) | 0.038 (0.0031) | 0.095 (0.0035) | 0.021 (0.0025) | 0.053 (0.0022) |
| ***CANN*** | 0.02 | 0.01 | 0.01 | NA | 0.051 (0.0029) | 0.036 (0.0029) | 0.032 (0.0012) | 0.009 (0.0019) | 0.049 (0.0018) | 0.016 (0.0018) | 0 (0) | 0.041 (0.0018) | 0.018 (0.0016) | 0.075 (0.0019) | 0.011 (0.0029) | 0.051 (0.0021) | 0.025 (0.0017) | 0.046 (0.0021) | 0.023 (0.0024) | 0.125 (0.0015) | 0.044 (0.0014) |
| ***COG*** | 0.05 | 0.01 | 0.19 | 0.04 | NA | 0.083 (0.0054) | 0.052 (0.002) | 0.06 (0.0033) | 0.031 (0.0032) | 0.045 (0.0032) | 0.041 (0.0037) | 0.046 (0.0033) | 0.045 (0.0028) | 0.051 (0.0037) | 0.202 (0.0048) | 0.026 (0.0041) | 0.045 (0.0032) | 0.03 (0.004) | 0.09 (0.0044) | 0.047 (0.0029) | 0.014 (0.0028) |
| ***SLEEP*** | 0.02 | 0.06 | 0.09 | 0.03 | 0.07 | NA | 0 (0) | 0.041 (0.0032) | 0.04 (0.003) | 0.054 (0.0031) | 0.064 (0.0035) | 0.05 (0.0032) | 0.099 (0.0025) | 0.035 (0.0036) | 0.109 (0.005) | 0.056 (0.0038) | 0.055 (0.003) | 0.062 (0.0037) | 0.076 (0.0043) | 0 (0) | 0.032 (0.0027) |
| ***LONE*** | 0.01 | 0.02 | 0.01 | 0.03 | 0.05 | 0.02 | NA | 0.053 (0.0012) | 0.02 (0.0012) | -0.019 (0.0014) | -0.006 (0.0015) | 0 (0.0013) | 0.003 (0.0011) | 0.018 (0.0014) | 0 (0.0021) | 0.013 (0.0015) | 0.03 (0.0012) | 0.018 (0.0015) | 0.055 (0.0016) | 0 (0.0013) | -0.007 (0.0012) |
| ***GUILT*** | 0.01 | 0.02 | 0.03 | 0 | 0.05 | 0.03 | 0.05 | NA | -0.006 (0.002) | 0.03 (0.002) | 0.047 (0.0022) | 0.027 (0.0021) | 0.033 (0.0017) | 0.012 (0.0024) | 0.035 (0.0033) | 0.033 (0.0025) | 0 (0) | 0.036 (0.0024) | 0.068 (0.0026) | 0.03 (0.0019) | 0.052 (0.0017) |
| ***HALLL*** | 0.03 | 0.03 | 0 | 0.04 | 0.02 | 0.03 | 0.02 | -0.01 | NA | 0 (0) | 0.024 (0.0021) | 0.03 (0.002) | 0.032 (0.0017) | 0.131 (0.002) | 0.035 (0.0031) | 0.033 (0.0024) | 0 (0.0019) | 0.087 (0.0021) | 0.024 (0.0025) | 0.019 (0.0018) | 0.023 (0.0016) |
| ***IRR*** | 0.08 | 0.06 | 0 | 0.01 | 0.04 | 0.04 | -0.02 | 0.03 | 0.03 | NA | 0.019 (0.0022) | 0.053 (0.002) | 0 (0) | 0.052 (0.0022) | 0.037 (0.0031) | 0 (0) | 0.043 (0.0019) | 0.017 (0.0023) | 0.04 (0.0026) | 0.03 (0.0018) | 0.005 (0.0017) |
| ***ENERGY*** | 0.01 | 0 | 0 | -0.01 | 0.03 | 0.05 | -0.01 | 0.04 | 0.02 | 0.02 | NA | 0 (0) | 0.016 (0.0019) | 0.031 (0.0024) | 0 (0) | 0.067 (0.0025) | 0.121 (0.0019) | 0.045 (0.0026) | 0.023 (0.003) | 0 (0) | 0.041 (0.0018) |
| ***MOOD*** | 0.04 | 0.03 | 0.01 | 0.04 | 0.04 | 0.04 | 0 | 0.02 | 0.02 | 0.05 | 0.01 | NA | 0.031 (0.0017) | 0.021 (0.0024) | 0.042 (0.0032) | 0.041 (0.0024) | 0.046 (0.002) | 0.039 (0.0024) | 0.041 (0.0027) | 0.041 (0.0018) | 0.021 (0.0017) |
| ***NIGHT*** | 0.01 | 0.02 | 0.04 | 0.02 | 0.04 | 0.09 | 0 | 0.03 | 0.03 | 0 | 0.01 | 0.03 | NA | 0.01 (0.0019) | 0.041 (0.0027) | -0.005 (0.0021) | 0.015 (0.0017) | 0.025 (0.002) | 0.024 (0.0023) | 0.016 (0.0015) | 0.014 (0.0015) |
| ***PAR*** | 0.04 | 0.07 | 0.04 | 0.07 | 0.04 | 0.03 | 0.01 | 0.01 | 0.13 | 0.04 | 0.02 | 0.01 | 0.01 | NA | 0.043 (0.0035) | 0.075 (0.0027) | -0.014 (0.0023) | 0.027 (0.0028) | 0.025 (0.0031) | 0.038 (0.0021) | 0.006 (0.0019) |

| | | | | | | | | | | | | | | | | | | | | | |
|---|---|---|---|---|---|---|---|---|---|---|---|---|---|---|---|---|---|---|---|---|---|
| ***CONC*** | 0 | 0.03 | 0.01 | 0 | 0.19 | 0.1 | 0 | 0.02 | 0.03 | 0.03 | 0.13 | 0.03 | 0.04 | 0.04 | NA | 0.075 (0.0037) | 0.033 (0.0032) | 0.05 (0.0037) | 0.061 (0.0042) | 0 (0.0029) | 0.05 (0.0026) |
| ***INS*** | 0.05 | 0.05 | 0.04 | 0.04 | 0.01 | 0.05 | 0.01 | 0.03 | 0.03 | 0 | 0.06 | 0.04 | -0.01 | 0.07 | 0.07 | NA | 0.021 (0.0023) | 0.035 (0.0028) | 0.058 (0.0031) | 0.016 (0.0022) | 0.016 (0.0021) |
| ***MOTIV*** | 0 | 0.01 | 0.01 | 0.02 | 0.04 | 0.05 | 0.03 | 0.04 | -0.01 | 0.04 | 0.12 | 0.04 | 0.01 | -0.02 | 0.02 | 0.01 | NA | 0.047 (0.0023) | 0.038 (0.0026) | 0.014 (0.0018) | 0.036 (0.0016) |
| ***SUIC*** | 0.03 | 0.04 | 0.03 | 0.04 | 0.02 | 0.05 | 0.02 | 0.03 | 0.08 | 0.01 | 0.04 | 0.04 | 0.02 | 0.02 | 0.04 | 0.03 | 0.04 | NA | 0.062 (0.0031) | 0.03 (0.0021) | 0.03 (0.0019) |
| ***TEAR*** | -0.01 | 0.04 | 0.09 | 0.02 | 0.08 | 0.07 | 0.05 | 0.06 | 0.02 | 0.04 | 0.01 | 0.03 | 0.02 | 0.02 | 0.05 | 0.05 | 0.03 | 0.06 | NA | 0.027 (0.0024) | 0.03 (0.0022) |
| ***TOB*** | 0.04 | 0.04 | 0.02 | 0.12 | 0.04 | 0.03 | 0 | 0.03 | 0.02 | 0.03 | 0.01 | 0.03 | 0.01 | 0.03 | 0 | 0.01 | 0.01 | 0.03 | 0.02 | NA | 0.01 (0.0015) |
| ***WGHT*** | 0.02 | 0.02 | 0.05 | 0.04 | 0 | 0.02 | -0.01 | 0.05 | 0.02 | 0 | 0.04 | 0.02 | 0.01 | 0 | 0.04 | 0.01 | 0.03 | 0.03 | 0.03 | 0.01 | NA |

### ii. Between-subject

| | **AGGR** | **AGIT** | **ANX** | **CANN** | **COG** | **SLEEP** | **LONE** | **GUILT** | **HALLL** | **IRR** | **ENERGY** | **MOOD** | **NIGHT** | **PAR** | **CONC** | **INS** | **MOTIV** | **SUIC** | **TEAR** | **TOB** | **WGHT** |
|---|---|---|---|---|---|---|---|---|---|---|---|---|---|---|---|---|---|---|---|---|---|
| ***AGGR*** | NA | 0.143 (0.0145) | -0.279 (0.0107) | 0.293 (0.0105) | 0.383 (0.0116) | 0.181 (0.0139) | -0.199 (0.0095) | 0.094 (0.0133) | 0 (0) | 0.443 (0.014) | -0.141 (0.0178) | -0.331 (0.0166) | -0.16 (0.0095) | 0.179 (0.0138) | -0.178 (0.0174) | 0 (0) | 0.045 (0.0141) | -0.093 (0.0133) | 0.052 (0.0137) | -0.21 (0.0117) | -0.245 (0.0145) |
| ***AGIT*** | 0.2 | NA | -0.019 (0.0095) | 0.067 (0.0088) | 0 (0) | 0.189 (0.0126) | -0.08 (0.0085) | -0.062 (0.0111) | 0.165 (0.0103) | 0.193 (0.017) | -0.306 (0.0141) | 0.08 (0.0177) | -0.173 (0.0088) | 0.101 (0.0124) | 0.279 (0.0157) | -0.143 (0.0145) | 0.26 (0.0114) | -0.137 (0.0117) | 0.173 (0.0114) | -0.097 (0.0109) | -0.115 (0.0127) |
| ***ANX*** | -0.23 | 0.03 | NA | 0.045 (0.0069) | 0.273 (0.0097) | 0.139 (0.0104) | -0.088 (0.0076) | 0.085 (0.0093) | -0.078 (0.0077) | 0.165 (0.0125) | -0.224 (0.0126) | -0.131 (0.0138) | -0.048 (0.008) | 0.063 (0.0096) | 0.088 (0.0134) | -0.024 (0.0115) | 0.224 (0.0096) | -0.026 (0.0093) | 0.121 (0.0097) | -0.08 (0.0082) | 0.062 (0.0117) |
| ***CANN*** | 0.25 | 0.04 | 0 | NA | -0.077 (0.0093) | -0.057 (0.0091) | 0.063 (0.0065) | 0.075 (0.0081) | -0.119 (0.0074) | -0.101 (0.0115) | 0.096 (0.0115) | 0.037 (0.0127) | 0.126 (0.0064) | 0.105 (0.0095) | -0.096 (0.0117) | 0.13 (0.0107) | 0 (0) | -0.06 (0.0085) | -0.08 (0.0086) | 0.441 (0.007) | 0.161 (0.0101) |
| ***COG*** | 0.33 | 0.03 | 0.27 | -0.06 | NA | -0.167 (0.012) | 0.123 (0.0091) | -0.185 (0.0109) | 0.121 (0.0097) | -0.404 (0.0123) | -0.133 (0.0139) | 0.445 (0.0126) | 0.091 (0.0089) | -0.266 (0.0112) | 0.421 (0.0121) | -0.178 (0.0133) | -0.053 (0.0117) | 0.302 (0.0106) | 0.112 (0.0111) | 0.093 (0.0107) | 0.297 (0.011) |
| ***SLEEP*** | 0.11 | 0.21 | 0.11 | -0.04 | -0.12 | NA | 0.043 (0.0103) | -0.186 (0.0121) | 0.167 (0.0101) | -0.127 (0.0166) | 0.332 (0.0156) | 0.21 (0.0164) | 0.32 (0.0095) | 0 (0.0127) | 0.149 (0.0165) | -0.029 (0.0144) | 0.037 (0.0131) | 0.245 (0.0119) | 0.265 (0.0126) | 0 (0.0105) | 0.28 (0.0117) |
| ***LONE*** | -0.16 | -0.09 | -0.05 | 0.04 | 0.11 | 0.06 | NA | 0.168 (0.0093) | -0.031 (0.0069) | 0.05 (0.0117) | -0.15 (0.0122) | -0.134 (0.0125) | -0.17 (0.0069) | 0.116 (0.0089) | 0.072 (0.012) | 0 (0.0106) | 0.18 (0.0094) | 0 (0.0082) | 0.287 (0.008) | 0 (0) | 0 (0.0102) |
| ***GUILT*** | 0.04 | -0.07 | 0.06 | 0.08 | -0.13 | -0.15 | 0.17 | NA | 0.02 (0.0085) | 0 (0) | 0.083 (0.0168) | 0.241 (0.0146) | 0.107 (0.0093) | -0.025 (0.0106) | 0.219 (0.0149) | -0.083 (0.0123) | -0.07 (0.0112) | 0.193 (0.0093) | 0.139 (0.0108) | 0 (0.0093) | 0 (0) |
| ***HALLL*** | 0.03 | 0.15 | -0.09 | -0.1 | 0.08 | 0.14 | -0.04 | 0.01 | NA | 0 (0.0131) | 0.09 (0.0126) | -0.175 (0.0128) | 0.153 (0.0071) | 0.272 (0.0089) | -0.161 (0.0132) | 0.269 (0.0103) | 0 (0) | -0.126 (0.0104) | -0.151 (0.0094) | 0.125 (0.0091) | 0 (0.0101) |
| ***IRR*** | 0.39 | 0.21 | 0.13 | -0.06 | -0.29 | -0.05 | 0 | -0.17 | -0.06 | NA | -0.149 (0.0188) | 0.653 (0.0126) | 0.126 (0.011) | -0.388 (0.0126) | 0.369 (0.0169) | -0.24 (0.0163) | -0.1 (0.0153) | 0.304 (0.0134) | 0 (0) | 0.295 (0.0128) | 0.308 (0.0144) |
| ***ENERGY*** | -0.17 | -0.25 | -0.19 | 0.06 | -0.05 | 0.38 | -0.17 | 0.12 | 0.05 | -0.1 | NA | 0.338 (0.0175) | -0.219 (0.0113) | -0.265 (0.0127) | 0.587 (0.0145) | 0 (0) | 0.357 (0.0141) | 0 (0) | 0.206 (0.0156) | -0.066 (0.0129) | 0 (0) |

| | | | | | | | | | | | | | | | | | | | | | |
|---|---|---|---|---|---|---|---|---|---|---|---|---|---|---|---|---|---|---|---|---|---|
| ***MOOD*** | -0.27 | 0.08 | -0.13 | 0.02 | 0.34 | 0.18 | -0.11 | 0.18 | -0.13 | 0.56 | 0.28 | NA | -0.125 (0.0116) | 0.438 (0.0126) | -0.556 (0.0143) | 0.448 (0.0144) | 0.079 (0.0173) | -0.321 (0.0137) | -0.036 (0.0165) | -0.184 (0.0147) | -0.31 (0.0149) |
| ***NIGHT*** | -0.12 | -0.16 | -0.01 | 0.09 | 0.07 | 0.29 | -0.16 | 0.07 | 0.16 | 0.09 | -0.21 | -0.1 | NA | -0.051 (0.0089) | 0.075 (0.0124) | -0.025 (0.0103) | 0 (0.0093) | -0.027 (0.0088) | 0.102 (0.0094) | -0.106 (0.0072) | -0.15 (0.0095) |
| ***PAR*** | 0.11 | 0.12 | 0.04 | 0.14 | -0.16 | 0.02 | 0.11 | 0.03 | 0.27 | -0.25 | -0.2 | 0.33 | -0.07 | NA | 0.305 (0.013) | 0 (0) | 0 (0.0117) | 0.228 (0.0111) | 0 (0.0109) | 0.084 (0.011) | 0.127 (0.0123) |
| ***CONC*** | -0.14 | 0.27 | 0.1 | -0.1 | 0.32 | 0.09 | 0.09 | 0.19 | -0.09 | 0.27 | 0.56 | -0.46 | 0.11 | 0.22 | NA | 0.445 (0.0135) | 0 (0) | -0.205 (0.0139) | -0.323 (0.0144) | 0.039 (0.0141) | -0.24 (0.0153) |
| ***INS*** | 0.01 | -0.11 | -0.01 | 0.11 | -0.07 | -0.01 | -0.02 | -0.04 | 0.22 | -0.17 | -0.21 | 0.36 | -0.04 | 0 | 0.36 | NA | 0 (0) | 0.358 (0.0111) | 0 (0) | 0.043 (0.0122) | 0.074 (0.0135) |
| ***MOTIV*** | 0.03 | 0.23 | 0.18 | 0.02 | -0.07 | 0.01 | 0.16 | -0.07 | -0.07 | -0.08 | 0.37 | 0.1 | -0.02 | 0.03 | -0.2 | -0.01 | NA | 0.135 (0.011) | -0.348 (0.0104) | 0 (0) | -0.109 (0.0121) |
| ***SUIC*** | -0.06 | -0.11 | 0.02 | -0.07 | 0.25 | 0.23 | 0.02 | 0.17 | -0.08 | 0.23 | 0.02 | -0.25 | 0.01 | 0.17 | -0.17 | 0.31 | 0.12 | NA | -0.024 (0.011) | -0.089 (0.01) | -0.07 (0.0111) |
| ***TEAR*** | 0.07 | 0.15 | 0.09 | -0.07 | 0.08 | 0.21 | 0.27 | 0.13 | -0.12 | -0.01 | 0.19 | 0.02 | 0.12 | -0.01 | -0.27 | 0.01 | -0.28 | -0.03 | NA | 0.135 (0.0102) | -0.183 (0.0123) |
| ***TOB*** | -0.13 | -0.07 | -0.02 | 0.42 | 0.03 | -0.01 | 0.02 | -0.05 | 0.14 | 0.22 | -0.07 | -0.1 | -0.08 | 0.01 | 0.09 | 0.02 | 0.11 | -0.02 | 0.1 | NA | 0 (0) |
| ***WGHT*** | -0.19 | -0.15 | 0.08 | 0.15 | 0.23 | 0.24 | 0 | 0 | 0.04 | 0.24 | 0.08 | -0.24 | -0.12 | 0.04 | -0.13 | 0.04 | -0.05 | -0.03 | -0.11 | 0.03 | NA |

### (B) BMD

#### i. Contemporaneous

| | AGGR | AGIT | ANX | CANN | COGN | SLEEP | ELAT | HOPE | GUILT | HALL | IRR | MOOD | PAR | CONC | INS | MOTIV | SUIC | TEAR | TOB |
|---|---|---|---|---|---|---|---|---|---|---|---|---|---|---|---|---|---|---|---|
| ***AGGR*** | NA | 0.087 (9e-04) | 0.044 (7e-04) | 0.058 (0.001) | 0.074 (7e-04) | 0.018 (8e-04) | 0.091 (9e-04) | -0.034 (8e-04) | -0.054 (9e-04) | 0.033 (9e-04) | 0.14 (9e-04) | 0.058 (8e-04) | 0.033 (9e-04) | 0.027 (9e-04) | 0.034 (8e-04) | 0.019 (8e-04) | 0.041 (9e-04) | -0.02 (8e-04) | 0 (0) |
| ***AGIT*** | 0.09 | NA | 0.058 (7e-04) | 0.038 (9e-04) | 0.047 (7e-04) | 0.023 (7e-04) | 0.118 (9e-04) | 0.005 (9e-04) | 0.008 (8e-04) | 0.022 (9e-04) | 0.065 (9e-04) | 0.007 (8e-04) | 0.069 (9e-04) | 0.049 (9e-04) | 0.019 (8e-04) | 0.011 (9e-04) | 0.04 (8e-04) | 0.072 (9e-04) | 0.034 (8e-04) |
| ***ANX*** | 0.05 | 0.06 | NA | 0.004 (6e-04) | 0.174 (8e-04) | 0.087 (7e-04) | 0.026 (7e-04) | -0.013 (6e-04) | 0.019 (7e-04) | 0 (0) | 0.023 (7e-04) | 0.04 (7e-04) | 0.047 (7e-04) | 0.016 (6e-04) | 0.042 (7e-04) | 0.042 (6e-04) | 0.012 (7e-04) | 0.097 (7e-04) | 0.042 (7e-04) |
| ***CANN*** | 0.06 | 0.04 | 0 | NA | -0.003 (7e-04) | 0.015 (7e-04) | 0.076 (9e-04) | 0.044 (9e-04) | 0.01 (8e-04) | 0.053 (9e-04) | 0.035 (8e-04) | 0.072 (8e-04) | 0.059 (0.001) | 0.003 (9e-04) | 0.043 (8e-04) | 0.028 (0.001) | -0.025 (9e-04) | -0.003 (8e-04) | 0.138 (0.001) |
| ***COGN*** | 0.07 | 0.05 | 0.18 | 0 | NA | 0.092 (7e-04) | 0.011 (7e-04) | 0.042 (7e-04) | 0.045 (7e-04) | 0.022 (7e-04) | 0.054 (7e-04) | 0.036 (7e-04) | 0.038 (7e-04) | 0.173 (7e-04) | 0.053 (7e-04) | 0.039 (7e-04) | 0.012 (7e-04) | 0.06 (7e-04) | 0.022 (7e-04) |
| ***SLEEP*** | 0.02 | 0.02 | 0.09 | 0.02 | 0.09 | NA | 0.062 (7e-04) | 0.021 (8e-04) | 0.024 (8e-04) | 0 (0) | 0.083 (8e-04) | 0.037 (7e-04) | 0.051 (8e-04) | 0.081 (8e-04) | 0.018 (7e-04) | 0.016 (8e-04) | 0.009 (7e-04) | 0.056 (8e-04) | 0.039 (8e-04) |
| ***ELAT*** | 0.09 | 0.12 | 0.03 | 0.07 | 0.01 | 0.06 | NA | -0.021 (8e-04) | 0 (0) | 0.02 (0.001) | 0.095 (9e-04) | 0.074 (8e-04) | 0.057 (0.001) | 0.062 (8e-04) | 0.072 (9e-04) | 0 (0) | -0.009 (9e-04) | 0.008 (8e-04) | 0.042 (8e-04) |
| ***HOPE*** | -0.03 | 0.01 | -0.01 | 0.05 | 0.04 | 0.02 | -0.02 | NA | 0.099 (0.0012) | 0.011 (9e-04) | -0.039 (8e-04) | 0.064 (9e-04) | 0 (0) | 0.077 (0.001) | 0.042 (8e-04) | 0.075 (0.0011) | 0.107 (0.0011) | 0.057 (9e-04) | -0.003 (9e-04) |
| ***GUILT*** | -0.06 | 0.01 | 0.02 | 0.01 | 0.05 | 0.02 | 0 | 0.1 | NA | -0.042 (9e-04) | 0.018 (8e-04) | 0.043 (8e-04) | 0 (9e-04) | 0 (0) | 0.028 (8e-04) | 0 (0) | -0.017 (8e-04) | 0.089 (9e-04) | -0.023 (9e-04) |
| ***HALL*** | 0.03 | 0.03 | 0 | 0.06 | 0.02 | 0.07 | 0.02 | 0.01 | -0.04 | NA | 0.013 (8e-04) | 0.065 (9e-04) | 0.1 (0.001) | 0 (9e-04) | 0.045 (9e-04) | 0 (9e-04) | 0.019 (0.001) | 0.02 (7e-04) | 0.01 (9e-04) |
| ***IRR*** | 0.14 | 0.06 | 0.03 | 0.03 | 0.06 | 0.08 | 0.09 | -0.04 | 0.02 | 0.01 | NA | 0.053 (8e-04) | 0.095 (8e-04) | -0.014 (9e-04) | 0.047 (8e-04) | -0.009 (8e-04) | 0 (8e-04) | 0.037 (8e-04) | 0.039 (9e-04) |
| ***MOOD*** | 0.06 | 0.01 | 0.04 | 0.07 | 0.03 | 0.04 | 0.07 | 0.06 | 0.04 | 0.06 | 0.05 | NA | 0.03 (8e-04) | 0.035 (9e-04) | 0.05 (7e-04) | -0.021 (8e-04) | 0 (0) | 0.089 (9e-04) | 0.033 (9e-04) |
| ***PAR*** | 0.03 | 0.07 | 0.05 | 0.06 | 0.04 | 0.05 | 0.06 | 0.02 | 0 | 0.1 | 0.1 | 0.03 | NA | 0.056 (0.001) | 0.077 (8e-04) | -0.041 (8e-04) | 0.046 (9e-04) | 0.058 (8e-04) | 0.016 (9e-04) |
| ***CONC*** | 0.03 | 0.05 | 0.01 | 0 | 0.17 | 0.08 | 0.06 | 0.08 | 0.07 | 0 | -0.01 | 0.03 | 0.06 | NA | 0.025 (0.001) | 0.025 (8e-04) | 0.018 (8e-04) | 0.067 (8e-04) | 0.019 (8e-04) |
| ***INS*** | 0.03 | 0.02 | 0.04 | 0.04 | 0.05 | 0.02 | 0.07 | 0.04 | 0.03 | 0.05 | 0.05 | 0.05 | 0.08 | 0.03 | NA | 0.032 (9e-04) | 0.051 (8e-04) | 0.025 (8e-04) | 0.022 (8e-04) |
| ***MOTIV*** | 0.02 | 0.01 | 0.04 | 0.03 | 0.04 | 0.01 | 0 | 0.08 | 0.11 | 0 | -0.01 | -0.02 | -0.04 | 0.03 | 0.03 | NA | 0.051 (9e-04) | 0.004 (8e-04) | -0.025 (9e-04) |
| ***SUIC*** | 0.04 | 0.04 | 0.01 | -0.02 | 0.01 | 0.01 | -0.01 | 0.11 | -0.02 | 0.02 | 0 | 0.03 | 0.05 | 0.02 | 0.06 | 0.05 | NA | 0.073 (0.001) | 0.021 (9e-04) |
| ***TEAR*** | -0.02 | 0.07 | 0.1 | -0.01 | 0.06 | 0.05 | 0.01 | 0.06 | 0.09 | 0.02 | 0.04 | 0.09 | 0.06 | 0.07 | 0.03 | 0 | 0.07 | NA | 0.027 (8e-04) |
| ***TOB*** | 0 | 0.04 | 0.04 | 0.14 | 0.02 | 0.04 | 0.04 | 0 | -0.02 | 0.01 | 0.04 | 0.03 | 0.02 | 0.02 | 0.02 | -0.03 | 0.02 | 0.03 | NA |

#### ii. Between-subject

| | AGGR | AGIT | ANX | CANN | COGN | SLEEP | ELAT | HOPE | GUILT | HALL | IRR | MOOD | PAR | CONC | INS | MOTIV | SUIC | TEAR | TOB |
|---|---|---|---|---|---|---|---|---|---|---|---|---|---|---|---|---|---|---|---|
| ***AGGR*** | NA | 0.396 (0.0214) | -0.209 (0.0233) | -0.441 (0.0183) | 0.714 (0.0145) | 0 (0) | 0.248 (0.0194) | 0.175 (0.0203) | -0.174 (0.0228) | -0.339 (0.0228) | 0.358 (0.0165) | -0.339 (0.0183) | 0.176 (0.0199) | -0.344 (0.0247) | 0.486 (0.0158) | -0.466 (0.0181) | 0.464 (0.0168) | 0.074 (0.026) | 0.165 (0.0208) |
| ***AGIT*** | 0.42 | NA | -0.205 (0.0253) | 0.465 (0.0184) | -0.39 (0.0183) | -0.19 (0.02) | -0.427 (0.0194) | 0.39 (0.0195) | -0.104 (0.0229) | -0.237 (0.0222) | 0.165 (0.018) | 0.112 (0.0213) | -0.225 (0.0196) | 0.485 (0.0231) | -0.447 (0.0169) | 0 (0) | -0.315 (0.0181) | -0.217 (0.0271) | 0 (0) |
| ***ANX*** | -0.09 | -0.72 | NA | 0.391 (0.0205) | 0.257 (0.024) | 0.345 (0.0208) | -0.324 (0.0234) | 0.568 (0.0193) | 0 (0) | -0.356 (0.0244) | 0.105 (0.0217) | 0.28 (0.0203) | -0.333 (0.0197) | -0.128 (0.0282) | -0.235 (0.0216) | -0.288 (0.0247) | -0.359 (0.0192) | 0 (0) | 0.082 (0.0229) |
| ***CANN*** | -0.69 | 0.91 | 0.55 | NA | 0.493 (0.0164) | -0.234 (0.0172) | 0.805 (0.007) | -0.498 (0.0132) | 0 (0) | 0.193 (0.0198) | 0 (0) | -0.576 (0.0121) | 0.45 (0.0172) | 0 (0) | 0.41 (0.0153) | -0.286 (0.0128) | 0.725 (0.01) | 0 (0) | 0.251 (0.016) |
| ***COGN*** | 0.9 | -0.45 | 0.19 | 0.69 | NA | -0.294 (0.0195) | -0.466 (0.0152) | -0.056 (0.019) | -0.043 (0.0203) | 0.214 (0.0226) | 0 (0) | 0.153 (0.0166) | 0.076 (0.022) | 0.384 (0.0229) | -0.295 (0.0162) | 0.613 (0.0151) | -0.321 (0.016) | -0.182 (0.0255) | -0.444 (0.0172) |

| | | | | | | | | | | | | | | | | | | | |
|---|---|---|---|---|---|---|---|---|---|---|---|---|---|---|---|---|---|---|---|
| ***SLEEP*** | 0.01 | 0.09 | 0.52 | -0.08 | -0.38 | NA | 0 (0) | -0.137 (0.017) | -0.344 (0.0194) | 0.095 (0.0191) | 0.672 (0.0128) | -0.738 (0.0101) | 0 (0) | 0.314 (0.0197) | 0.485 (0.0172) | 0.41 (0.0171) | 0 (0) | -0.378 (0.023) | 0 (0) |
| ***ELAT*** | 0.45 | -0.87 | -0.44 | 0.87 | -0.64 | -0.34 | NA | 0.634 (0.0115) | -0.357 (0.0197) | -0.361 (0.0201) | 0.469 (0.0097) | 0.205 (0.0124) | 0 (0) | 0 (0) | 0.038 (0.0142) | 0.373 (0.0117) | -0.377 (0.0108) | -0.19 (0.0237) | -0.575 (0.0103) |
| ***HOPE*** | 0.19 | 0.78 | 0.87 | -0.56 | -0.1 | -0.12 | 0.66 | NA | 0.545 (0.0168) | 0.755 (0.0133) | -0.409 (0.0117) | -0.237 (0.0138) | 0.247 (0.0167) | -0.088 (0.0221) | 0 (0.0166) | 0.396 (0.0141) | 0.329 (0.0129) | 0.18 (0.0237) | 0 (0.0166) |
| ***GUILT*** | -0.54 | -0.2 | -0.19 | -0.04 | 0.2 | -0.58 | -0.42 | 0.59 | NA | -0.586 (0.0188) | 0.628 (0.0164) | 0 (0) | 0.389 (0.0171) | 0.355 (0.0217) | 0.438 (0.0162) | -0.194 (0.0209) | 0 (0) | -0.399 (0.0223) | -0.327 (0.0195) |
| ***HALL*** | -0.62 | -0.4 | -0.7 | 0.09 | 0.52 | 0.11 | -0.29 | 0.86 | -0.72 | NA | 0.412 (0.0165) | 0 (0.0182) | -0.042 (0.0204) | 0.108 (0.0276) | 0 (0) | -0.496 (0.0199) | 0 (0.0187) | -0.206 (0.0237) | 0 (0) |
| ***IRR*** | 0.33 | 0.18 | -0.03 | -0.02 | 0.07 | 0.82 | 0.51 | -0.41 | 0.93 | 0.47 | NA | 0.66 (0.01) | -0.594 (0.013) | -0.042 (0.0203) | -0.663 (0.012) | -0.14 (0.0124) | -0.532 (0.0107) | 0.344 (0.0231) | 0.643 (0.0119) |
| ***MOOD*** | -0.51 | 0.56 | 0.62 | -0.64 | 0.2 | -0.79 | 0.21 | -0.29 | -0.58 | -0.02 | 0.69 | NA | 0.726 (0.0129) | 0.106 (0.0196) | 0.657 (0.0152) | 0.158 (0.0123) | 0.893 (0.006) | -0.22 (0.0243) | -0.428 (0.0143) |
| ***PAR*** | 0.27 | -0.43 | -0.66 | 0.45 | 0.07 | 0.91 | -0.02 | 0.31 | 0.55 | -0.11 | -0.72 | 0.95 | NA | 0 (0.0221) | -0.656 (0.0144) | -0.378 (0.0163) | -0.685 (0.0128) | 0.236 (0.024) | 0.584 (0.0162) |
| ***CONC*** | 0.12 | 0.15 | 0.13 | 0.02 | -0.1 | -0.02 | 0 | -0.18 | 0.18 | 0.22 | -0.06 | 0.01 | 0.04 | NA | 0.269 (0.0206) | 0.168 (0.0252) | 0 (0) | 0 (0) | 0 (0.0208) |
| ***INS*** | 0.51 | -0.43 | -0.51 | 0.53 | -0.17 | 0.83 | -0.06 | 0.14 | 0.7 | 0.14 | -0.8 | 0.98 | -0.94 | 0.02 | NA | -0.137 (0.0154) | -0.726 (0.0113) | 0.126 (0.0251) | 0.477 (0.0152) |
| ***MOTIV*** | -0.74 | 0.12 | -0.51 | -0.37 | 0.9 | 0.65 | 0.45 | 0.36 | -0.1 | -0.64 | -0.23 | 0.18 | -0.43 | 0.21 | -0.18 | NA | 0 (0.0139) | 0 (0.0275) | 0.73 (0.0112) |
| ***SUIC*** | 0.62 | -0.7 | -0.63 | 0.79 | -0.37 | 0.64 | -0.41 | 0.37 | 0.49 | 0.01 | -0.55 | 0.97 | -0.87 | 0 | -0.93 | -0.01 | NA | 0.182 (0.024) | 0.251 (0.0151) |
| ***TEAR*** | -0.35 | -0.22 | -0.12 | 0.04 | 0.03 | -0.61 | -0.45 | 0.48 | -0.85 | -0.54 | 0.85 | -0.52 | 0.54 | 0.21 | 0.63 | 0.06 | 0.42 | NA | -0.229 (0.0249) |
| ***TOB*** | 0.34 | -0.21 | 0.35 | 0.3 | -0.68 | -0.91 | -0.63 | 0 | -0.46 | 0.18 | 0.72 | -0.49 | 0.69 | -0.1 | 0.56 | 0.83 | 0.29 | -0.52 | NA |

### (C) PSY

#### i. Contemporaneous

| | AGGR | AGIT | ANX | AROUS | CANN | COC | COG | DEL | SLEEP | ELAT | EMOT | HOPE | HALL | HOST | IRR | MOOD | PAR | CONC | INS | MOTIV | SUIC | TEAR | TOB | WGHT |
|---|---|---|---|---|---|---|---|---|---|---|---|---|---|---|---|---|---|---|---|---|---|---|---|---|
| ***AGGR*** | NA | 0.413 (0.0138) | 0.246 (0.0099) | 0.294 (0.0114) | 0.255 (0.0111) | 0.201 (0.0092) | 0.313 (0.0117) | 0.352 (0.0128) | 0.244 (0.0109) | 0.268 (0.0103) | 0.193 (0.0085) | 0.126 (0.0077) | 0.306 (0.0119) | 0.378 (0.0118) | 0.381 (0.0128) | 0.254 (0.0097) | 0.361 (0.0132) | 0.187 (0.0089) | 0.231 (0.0094) | 0.112 (0.0051) | 0.188 (0.0075) | 0.178 (0.0082) | 0.289 (0.0101) | 0.136 (0.0056) |
| ***AGIT*** | 0.1 | NA | 0.321 (0.0109) | 0.364 (0.0118) | 0.257 (0.0106) | 0.212 (0.0091) | 0.305 (0.012) | 0.359 (0.0134) | 0.322 (0.0112) | 0.266 (0.0111) | 0.178 (0.008) | 0.137 (0.0074) | 0.337 (0.0124) | 0.322 (0.0114) | 0.387 (0.0131) | 0.262 (0.0102) | 0.378 (0.0144) | 0.245 (0.0093) | 0.262 (0.0099) | 0.101 (0.0059) | 0.235 (0.0093) | 0.265 (0.0097) | 0.293 (0.0102) | 0.14 (0.0059) |
| ***ANX*** | 0.03 | 0.08 | NA | 0.161 (0.0074) | 0.212 (0.0081) | 0.126 (0.0059) | 0.401 (0.0113) | 0.246 (0.0101) | 0.305 (0.0096) | 0.146 (0.0063) | 0.136 (0.0052) | 0.12 (0.0048) | 0.263 (0.0096) | 0.159 (0.007) | 0.221 (0.0087) | 0.179 (0.0074) | 0.336 (0.0114) | 0.206 (0.0079) | 0.22 (0.0081) | 0.107 (0.005) | 0.146 (0.0064) | 0.209 (0.0069) | 0.197 (0.0076) | 0.129 (0.0047) |
| ***AROUS*** | 0.05 | 0.11 | 0.01 | NA | 0.187 (0.0074) | 0.142 (0.006) | 0.186 (0.0074) | 0.271 (0.01) | 0.167 (0.0069) | 0.28 (0.0096) | 0.108 (0.005) | 0.051 (0.0037) | 0.188 (0.0081) | 0.329 (0.0105) | 0.31 (0.0105) | 0.206 (0.0075) | 0.226 (0.0099) | 0.143 (0.0063) | 0.169 (0.0064) | 0.049 (0.0049) | 0.128 (0.0058) | 0.189 (0.0077) | 0.188 (0.0074) | 0.091 (0.0038) |
| ***CANN*** | 0.02 | 0.02 | 0.04 | 0.02 | NA | 0.395 (0.0107) | 0.241 (0.0093) | 0.282 (0.0108) | 0.194 (0.0082) | 0.191 (0.0074) | 0.118 (0.0055) | 0.124 (0.0048) | 0.251 (0.0099) | 0.193 (0.0076) | 0.236 (0.0092) | 0.19 (0.0074) | 0.323 (0.0114) | 0.167 (0.0074) | 0.229 (0.0084) | 0.113 (0.0043) | 0.16 (0.0065) | 0.131 (0.0066) | 0.308 (0.0094) | 0.127 (0.0048) |
| ***COC*** | 0.03 | 0.03 | -0.01 | 0.01 | 0.26 | NA | 0.164 (0.007) | 0.205 (0.0082) | 0.133 (0.0061) | 0.152 (0.0059) | 0.098 (0.004) | 0.077 (0.0036) | 0.204 (0.0079) | 0.128 (0.0058) | 0.173 (0.007) | 0.138 (0.0057) | 0.196 (0.0084) | 0.128 (0.0056) | 0.16 (0.0062) | 0.076 (0.0033) | 0.114 (0.0051) | 0.133 (0.0052) | 0.17 (0.0067) | 0.093 (0.0035) |
| ***COG*** | 0.06 | 0.03 | 0.17 | 0.01 | 0.03 | 0.02 | NA | 0.318 (0.0113) | 0.308 (0.0106) | 0.161 (0.0071) | 0.162 (0.0067) | 0.137 (0.0057) | 0.313 (0.0113) | 0.201 (0.0079) | 0.274 (0.0101) | 0.211 (0.0082) | 0.368 (0.0126) | 0.324 (0.0098) | 0.236 (0.0092) | 0.173 (0.0055) | 0.165 (0.0068) | 0.214 (0.0078) | 0.228 (0.0086) | 0.15 (0.0056) |
| ***DEL*** | 0.06 | 0.05 | 0.01 | 0.05 | 0.05 | 0.03 | 0.06 | NA | 0.256 (0.0104) | 0.251 (0.0089) | 0.155 (0.0065) | 0.133 (0.0056) | 0.444 (0.0132) | 0.266 (0.0097) | 0.288 (0.0113) | 0.233 (0.0089) | 0.444 (0.0141) | 0.226 (0.0094) | 0.298 (0.01) | 0.121 (0.005) | 0.212 (0.0077) | 0.192 (0.0081) | 0.239 (0.0093) | 0.149 (0.0054) |
| ***SLEEP*** | 0.02 | 0.06 | 0.09 | 0 | 0.01 | 0 | 0.06 | 0.01 | NA | 0.196 (0.0069) | 0.169 (0.0058) | 0.156 (0.0055) | 0.285 (0.0102) | 0.173 (0.0073) | 0.269 (0.0095) | 0.216 (0.0078) | 0.308 (0.011) | 0.273 (0.0089) | 0.268 (0.0089) | 0.145 (0.0058) | 0.198 (0.007) | 0.247 (0.008) | 0.236 (0.0081) | 0.148 (0.0054) |
| ***ELAT*** | 0.06 | 0.04 | 0 | 0.09 | 0.03 | 0.03 | -0.01 | 0.05 | 0.04 | NA | 0.084 (0.0042) | 0.031 (0.0033) | 0.184 (0.0075) | 0.246 (0.0086) | 0.278 (0.0095) | 0.239 (0.0074) | 0.201 (0.0087) | 0.171 (0.0059) | 0.197 (0.0066) | 0.058 (0.0033) | 0.116 (0.0051) | 0.158 (0.0066) | 0.178 (0.0071) | 0.068 (0.0034) |
| ***EMOT*** | 0.04 | 0.02 | 0.01 | 0 | 0 | 0.02 | 0.01 | 0.01 | 0.03 | -0.02 | NA | 0.106 (0.0035) | 0.184 (0.0067) | 0.143 (0.0052) | 0.178 (0.0062) | 0.142 (0.005) | 0.194 (0.0069) | 0.158 (0.0052) | 0.144 (0.0053) | 0.106 (0.0034) | 0.128 (0.0044) | 0.144 (0.0049) | 0.139 (0.0051) | 0.111 (0.0035) |
| ***HOPE*** | 0.01 | 0.02 | 0.01 | -0.02 | 0.02 | 0 | 0.01 | 0.01 | 0.03 | -0.03 | 0.03 | NA | 0.15 (0.0056) | 0.059 (0.0037) | 0.1 (0.0045) | 0.143 (0.0046) | 0.126 (0.0059) | 0.166 (0.0055) | 0.144 (0.0052) | 0.133 (0.0047) | 0.209 (0.0054) | 0.181 (0.0049) | 0.104 (0.0041) | 0.106 (0.0034) |
| ***HALL)*** | 0.04 | 0.06 | 0.04 | 0 | 0.03 | 0.04 | 0.05 | 0.17 | 0.05 | 0.01 | 0.03 | 0.02 | NA | 0.223 (0.0086) | 0.249 (0.0102) | 0.202 (0.0085) | 0.417 (0.0134) | 0.248 (0.0089) | 0.304 (0.0101) | 0.157 (0.0051) | 0.224 (0.0076) | 0.191 (0.0077) | 0.234 (0.009) | 0.117 (0.0054) |
| ***HOST*** | 0.13 | 0.06 | -0.01 | 0.12 | 0.02 | 0 | 0.01 | 0.04 | 0 | 0.06 | 0.03 | -0.02 | 0.03 | NA | 0.351 (0.0109) | 0.201 (0.0072) | 0.249 (0.0097) | 0.131 (0.006) | 0.155 (0.0066) | 0.043 (0.0032) | 0.125 (0.0054) | 0.176 (0.0067) | 0.2 (0.0075) | 0.093 (0.0041) |
| ***IRR*** | 0.1 | 0.09 | 0.01 | 0.08 | 0.03 | 0.02 | 0.04 | 0.02 | 0.05 | 0.07 | 0.04 | 0 | 0.01 | 0.11 | NA | 0.273 (0.009) | 0.3 (0.0115) | 0.227 (0.008) | 0.213 (0.0084) | 0.069 (0.0039) | 0.152 (0.0066) | 0.216 (0.0081) | 0.244 (0.009) | 0.144 (0.005) |
| ***MOOD*** | 0.04 | 0.03 | 0.01 | 0.04 | 0.03 | 0.01 | 0.02 | 0.02 | 0.03 | 0.08 | 0.03 | 0.04 | 0 | 0.03 | 0.07 | NA | 0.251 (0.0093) | 0.229 (0.0073) | 0.195 (0.0072) | 0.113 (0.0038) | 0.157 (0.0058) | 0.235 (0.0072) | 0.184 (0.0073) | 0.14 (0.0047) |

| | | | | | | | | | | | | | | | | | | | | | | | | |
|---|---|---|---|---|---|---|---|---|---|---|---|---|---|---|---|---|---|---|---|---|---|---|---|---|
| ***PAR*** | 0.07 | 0.06 | 0.09 | 0.01 | 0.09 | 0.01 | 0.09 | 0.14 | 0.05 | 0 | 0.04 | -0.01 | 0.12 | 0.03 | 0.03 | 0.04 | NA | 0.224 (0.0093) | 0.308 (0.0106) | 0.14 (0.0055) | 0.195 (0.0078) | 0.214 (0.0086) | 0.26 (0.0101) | 0.153 (0.0057) |
| ***CONC*** | -0.01 | 0.03 | 0.02 | 0.01 | 0 | 0.01 | 0.11 | 0.02 | 0.07 | 0.03 | 0.04 | 0.05 | 0.04 | -0.01 | 0.04 | 0.06 | 0.01 | NA | 0.231 (0.0078) | 0.173 (0.0054) | 0.192 (0.0062) | 0.217 (0.0068) | 0.18 (0.0067) | 0.132 (0.0046) |
| ***INS*** | 0.02 | 0.03 | 0.03 | 0.02 | 0.04 | 0.02 | 0.02 | 0.06 | 0.06 | 0.05 | 0.02 | 0.03 | 0.07 | 0 | 0.01 | 0.02 | 0.06 | 0.05 | NA | 0.125 (0.0048) | 0.191 (0.0064) | 0.185 (0.0066) | 0.209 (0.0074) | 0.112 (0.0044) |
| ***MOTIV*** | 0.01 | -0.01 | 0.01 | 0 | 0.02 | 0.01 | 0.05 | 0.01 | 0.03 | 0 | 0.04 | 0.06 | 0.04 | -0.02 | -0.02 | 0.03 | 0.02 | 0.06 | 0.02 | NA | 0.118 (0.0038) | 0.078 (0.0033) | 0.09 (0.0038) | 0.073 (0.0027) |
| ***SUIC*** | 0.02 | 0.04 | 0 | 0 | 0.02 | 0.01 | 0 | 0.04 | 0.04 | 0.01 | 0.03 | 0.11 | 0.05 | 0 | 0 | 0.02 | 0.01 | 0.05 | 0.04 | 0.03 | NA | 0.209 (0.0063) | 0.166 (0.0059) | 0.104 (0.0037) |
| ***TEAR*** | -0.01 | 0.05 | 0.05 | 0.04 | -0.02 | 0.03 | 0.03 | 0.01 | 0.07 | 0.02 | 0.03 | 0.08 | 0.01 | 0.03 | 0.03 | 0.08 | 0.02 | 0.06 | 0.03 | 0 | 0.07 | NA | 0.162 (0.0065) | 0.133 (0.0043) |
| ***TOB*** | 0.07 | 0.06 | 0.02 | 0.02 | 0.13 | 0.02 | 0.03 | 0.02 | 0.05 | 0.02 | 0.02 | 0.01 | 0.02 | 0.03 | 0.04 | 0.02 | 0.03 | 0.02 | 0.04 | 0 | 0.03 | 0.01 | NA | 0.134 (0.0047) |
| ***WGHT*** | 0.02 | 0.01 | 0.02 | 0 | 0.02 | 0.02 | 0.02 | 0.03 | 0.03 | -0.01 | 0.04 | 0.03 | -0.01 | 0 | 0.03 | 0.04 | 0.02 | 0.03 | 0.01 | 0.02 | 0.02 | 0.03 | 0.03 | NA |

### ii. Between-subject

| | AGGR | AGIT | ANX | AROUS | CANN | COC | COG | DEL | SLEEP | ELAT | EMOT | HOPE | HALL | HOST | IRR | MOOD | PAR | CONC | INS | MOTIV | SUIC | TEAR | TOB | WGHT |
|---|---|---|---|---|---|---|---|---|---|---|---|---|---|---|---|---|---|---|---|---|---|---|---|---|
| ***AGGR*** | NA | 0.073 (0.005) | 0 (0) | 0.059 (0.0045) | 0.045 (0.0025) | 0.016 (0.0012) | 0.08 (0.0065) | 0 (0) | -0.03 (0.0058) | -0.013 (0.0044) | 0 (0) | -0.009 (0.0025) | 0.02 (0.0015) | 0.049 (0.0026) | 0.079 (0.0057) | 0.062 (0.0053) | 0.032 (0.0011) | 0.007 (0.0019) | 0.014 (0.0015) | -0.007 (0.0022) | 0.026 (0.0026) | 0 (0) | 0.039 (0.0026) | 0 (0) |
| ***AGIT*** | 0.23 | NA | 0.063 (0.005) | 0.046 (0.0032) | 0.024 (0.0013) | 0.022 (0.0014) | 0.02 (0.0022) | 0.054 (0.0033) | 0.045 (0.0032) | 0.058 (0.0048) | 0.051 (0.0054) | 0 (0) | 0.045 (0.0032) | 0.044 (0.0026) | 0.057 (0.0035) | -0.01 (0.0044) | 0.029 (0.0013) | 0.006 (0.0023) | -0.028 (0.0057) | 0 (0) | 0.033 (0.0033) | 0.014 (0.0017) | -0.021 (0.0049) | 0.018 (0.002) |
| ***ANX*** | -0.11 | 0.2 | NA | 0.014 (0.0023) | 0 (0) | 0.014 (0.0012) | 0.133 (0.0116) | 0 (0) | 0.054 (0.0034) | -0.027 (0.0049) | 0 (0) | 0.019 (0.0017) | -0.016 (0.0048) | -0.012 (0.0033) | 0 (0) | 0.068 (0.0065) | 0.075 (0.0057) | 0.059 (0.0048) | 0.061 (0.0052) | -0.016 (0.0038) | 0 (0) | 0.028 (0.0023) | 0.018 (0.0015) | 0.014 (0.0019) |
| ***AROUS*** | 0.22 | 0.12 | 0.02 | NA | 0.016 (0.0013) | 0.026 (0.002) | -0.009 (0.0035) | 0.027 (0.0017) | 0.016 (0.002) | 0.051 (0.0041) | 0 (0) | 0.02 (0.003) | -0.018 (0.0035) | 0.055 (0.004) | 0.024 (0.0022) | 0 (0) | 0.025 (0.0017) | -0.011 (0.0028) | -0.008 (0.0029) | -0.024 (0.0036) | -0.006 (0.0024) | 0 (0) | -0.018 (0.004) | 0.004 (0.0021) |
| ***CANN*** | 0.11 | 0.04 | -0.06 | -0.01 | NA | 0.124 (0.0093) | 0.019 (0.0011) | 0.024 (0.001) | -0.012 (0.003) | 0.014 (0.0011) | -0.006 (0.002) | 0 (0) | 0.01 (0.0013) | 0.016 (0.0014) | 0.015 (0.0016) | 0.009 (0.0017) | 0.045 (0.0022) | 0.01 (0.0013) | 0.06 (0.0053) | 0.017 (0.0015) | -0.008 (0.0025) | 0.024 (0.0025) | 0.105 (0.0088) | 0 (0) |
| ***COC*** | 0.01 | 0.05 | 0.04 | 0.1 | 0.41 | NA | -0.009 (0.0024) | 0.006 (0.0013) | 0 (0) | 0 (0) | -0.017 (0.0025) | 0 (0) | 0.024 (0.0016) | 0.007 (0.0014) | 0.004 (0.0016) | 0.026 (0.0024) | 0.026 (0.0013) | -0.011 (0.0019) | -0.009 (0.0025) | 0.012 (0.0014) | 0.031 (0.0027) | -0.011 (0.0021) | 0 (0) | 0.027 (0.0027) |
| ***COG*** | 0.26 | -0.01 | 0.49 | -0.12 | 0.03 | -0.08 | NA | 0.046 (0.0027) | 0.022 (0.0015) | 0.019 (0.0017) | 0.06 (0.006) | 0.023 (0.0019) | 0.045 (0.0029) | 0.019 (0.0018) | 0.04 (0.0024) | -0.016 | 0.013 (0.0025) | 0.027 (0.0018) | 0.013 (0.002) | 0.037 (0.003) | 0.018 (0.0022) | 0.031 (0.0024) | 0.017 (0.0013) | 0.023 (0.0022) |

| | | | | | | | | | | | | | | | | | | | | | | | | |
|---|---|---|---|---|---|---|---|---|---|---|---|---|---|---|---|---|---|---|---|---|---|---|---|---|
| | | | | | | | | | | | | | | | | (0.0047) | | | | | | | | |
| ***DEL*** | -0.07 | 0.16 | -0.08 | 0.09 | 0.02 | -0.03 | 0.11 | NA | 0 (0.002) | 0.009 (0.0015) | -0.007 (0.0025) | 0 (0) | 0.041 (0.0018) | 0.035 (0.0021) | 0.016 (0.0017) | 0.01 (0.0015) | 0 (0) | 0.022 (0.0016) | 0.043 (0.0029) | -0.011 (0.0024) | 0.01 (0.0014) | -0.013 (0.0029) | 0.014 (0.001) | -0.004 (0.002) |
| ***SLEEP*** | -0.2 | 0.13 | 0.15 | 0.05 | -0.11 | -0.02 | -0.01 | -0.05 | NA | 0.013 (0.0013) | -0.012 (0.0035) | 0.042 (0.0034) | 0.052 (0.0037) | -0.02 (0.0041) | 0.076 (0.0071) | 0.022 (0.0018) | 0.032 (0.0015) | 0.046 (0.0032) | 0.04 (0.0028) | 0.023 (0.0021) | 0.036 (0.0028) | 0.025 (0.0017) | 0.033 (0.0026) | 0 (0.0017) |
| ***ELAT*** | -0.13 | 0.17 | -0.16 | 0.18 | 0.01 | -0.05 | 0.05 | -0.02 | 0.01 | NA | -0.018 (0.0034) | -0.008 (0.0023) | -0.023 (0.004) | 0 (0) | 0.047 (0.0032) | 0.096 (0.0094) | 0.007 (0.0012) | 0.026 (0.0025) | 0.05 (0.0046) | 0 (0) | 0 (0) | 0 (0) | 0.01 (0.0015) | 0.006 (0.0016) |
| ***EMOT*** | -0.03 | 0.22 | -0.1 | 0.05 | -0.08 | -0.08 | 0.22 | -0.06 | -0.08 | -0.11 | NA | 0.007 (0.0024) | 0 (0) | -0.018 (0.0039) | -0.009 (0.0034) | 0.057 (0.0065) | 0.013 (0.0016) | 0.044 (0.0045) | -0.009 (0.0035) | 0.039 (0.0043) | -0.026 (0.0052) | -0.039 (0.0059) | 0.016 (0.0025) | 0.014 (0.0025) |
| ***HOPE*** | -0.08 | -0.04 | 0.02 | 0.09 | -0.01 | -0.02 | 0.06 | 0.08 | 0.14 | -0.06 | -0.01 | NA | 0.019 (0.0014) | 0 (0) | -0.019 (0.0034) | 0.01 (0.0024) | 0.01 (0.0012) | 0 (0.0031) | 0.023 (0.0021) | 0.048 (0.0048) | 0.098 (0.0093) | 0.054 (0.0044) | 0 (0.0017) | 0 (0) |
| ***HALL)*** | 0.03 | 0.13 | -0.15 | -0.09 | -0.02 | 0.07 | 0.13 | 0.1 | 0.16 | -0.14 | -0.01 | 0.03 | NA | 0 (0.0016) | -0.016 (0.0037) | 0.02 (0.002) | 0.033 (0.0013) | 0 (0) | 0.055 (0.0044) | -0.021 (0.0039) | 0.006 (0.002) | 0.008 (0.0014) | 0 (0) | 0 (0.002) |
| ***HOST*** | 0.1 | 0.11 | -0.11 | 0.18 | 0 | -0.02 | 0.03 | 0.08 | -0.13 | -0.06 | -0.09 | 0.09 | -0.03 | NA | 0.118 (0.0105) | 0.045 (0.004) | 0.043 (0.0029) | 0.004 (0.0018) | 0 (0) | -0.006 (0.0022) | -0.012 (0.0026) | -0.017 (0.0036) | 0.02 (0.0016) | 0.034 (0.0038) |
| ***IRR*** | 0.25 | 0.14 | -0.06 | 0.02 | 0 | -0.04 | 0.09 | -0.02 | 0.29 | 0.13 | -0.09 | -0.11 | -0.12 | 0.43 | NA | 0.019 (0.0023) | 0.023 (0.0017) | 0.015 (0.0014) | 0.024 (0.0028) | 0.015 (0.0021) | -0.024 (0.0044) | 0.038 (0.0034) | 0.024 (0.0017) | 0.005 (0.0019) |
| ***MOOD*** | 0.22 | -0.1 | 0.25 | -0.05 | -0.02 | 0.08 | -0.16 | -0.01 | 0.03 | 0.37 | 0.24 | 0.03 | 0.04 | 0.13 | 0.03 | NA | 0 (0.0031) | -0.015 (0.0042) | 0.033 (0.0028) | 0.023 (0.0027) | 0.042 (0.0045) | 0.082 (0.0078) | 0.021 (0.002) | -0.029 (0.0049) |
| ***PAR*** | 0.04 | 0.02 | 0.24 | 0.04 | 0.11 | 0.05 | -0.04 | 0.31 | 0.07 | -0.02 | 0.02 | 0.01 | 0.04 | 0.12 | 0.02 | -0.1 | NA | 0 (0.0025) | 0.03 (0.0016) | 0.027 (0.0025) | 0.02 (0.0016) | -0.008 (0.0026) | 0.02 (0.0011) | -0.009 (0.0025) |
| ***CONC*** | 0.01 | -0.04 | 0.18 | -0.07 | 0 | -0.05 | 0.05 | 0.07 | 0.14 | 0.09 | 0.18 | -0.06 | 0.11 | -0.01 | 0.02 | -0.11 | -0.06 | NA | 0.015 (0.002) | 0.057 (0.0055) | 0 (0) | 0.028 (0.0023) | -0.019 (0.0038) | 0.033 (0.0035) |
| ***INS*** | 0.03 | -0.19 | 0.2 | -0.04 | 0.21 | -0.07 | -0.03 | 0.14 | 0.14 | 0.16 | 0.01 | 0.04 | 0.2 | -0.03 | 0.03 | 0.12 | 0.05 | 0.01 | NA | 0.014 (0.0018) | 0.031 (0.0028) | -0.025 (0.0051) | -0.013 (0.0038) | 0.023 (0.0026) |
| ***MOTIV*** | -0.05 | -0.02 | -0.11 | -0.12 | 0.05 | 0.04 | 0.12 | -0.08 | 0.06 | -0.02 | 0.15 | 0.18 | -0.14 | -0.04 | 0.05 | 0.07 | 0.08 | 0.21 | 0.05 | NA | 0 (0.0028) | 0.005 (0.0017) | 0.022 (0.0023) | 0 (0) |
| ***SUIC*** | 0.1 | 0.1 | -0.05 | -0.06 | -0.07 | 0.11 | 0.02 | 0 | 0.11 | -0.03 | -0.13 | 0.37 | -0.02 | -0.08 | -0.14 | 0.13 | 0.04 | 0.18 | 0.09 | -0.06 | NA | 0.03 (0.0026) | 0.036 (0.0033) | 0 (0) |

| | | | | | | | | | | | | | | | | | | | | | | | | |
|---|---|---|---|---|---|---|---|---|---|---|---|---|---|---|---|---|---|---|---|---|---|---|---|---|
| ***TEAR*** | -0.07 | 0.01 | 0.05 | 0 | 0.08 | -0.06 | 0.09 | -0.1 | 0.06 | -0.07 | -0.18 | 0.17 | 0 | -0.09 | 0.11 | 0.32 | -0.08 | 0.07 | -0.14 | 0 | 0.08 | NA | -0.02 (0.0037) | 0.015 (0.0021) |
| ***TOB*** | 0.12 | -0.17 | 0.02 | -0.07 | 0.37 | -0.04 | 0 | 0.03 | 0.12 | 0 | 0.12 | -0.03 | 0.06 | 0.05 | 0.04 | 0.05 | 0.01 | -0.11 | -0.09 | 0.08 | 0.12 | -0.1 | NA | 0.027 (0.0028) |
| ***WGHT*** | -0.04 | 0.07 | 0.02 | 0 | -0.04 | 0.1 | 0.06 | -0.05 | -0.02 | 0 | 0.07 | 0.17 | -0.03 | 0.14 | -0.01 | -0.13 | -0.07 | 0.14 | 0.1 | -0.02 | -0.03 | 0.04 | 0.11 | NA |

eFigure 5 Un-thresholded sub-network graphs: (i) UMD, (ii) BMD and (iii) PSY for (A) Temporal, (B) Contemporaneous and (C) Between-subject matrices. Edge weights are not presented for visualisation purposes but can be found in eTable 9.

**i. UMD**

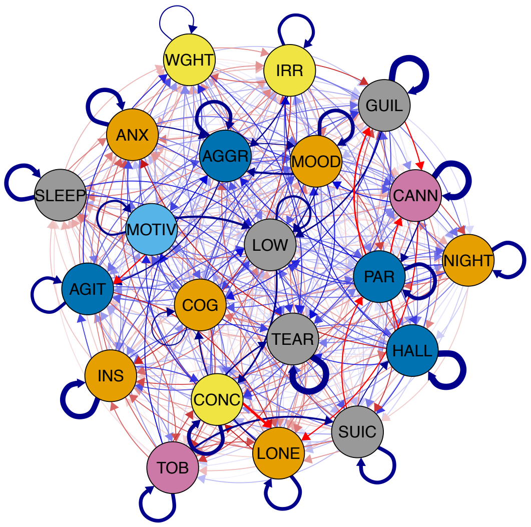

A.

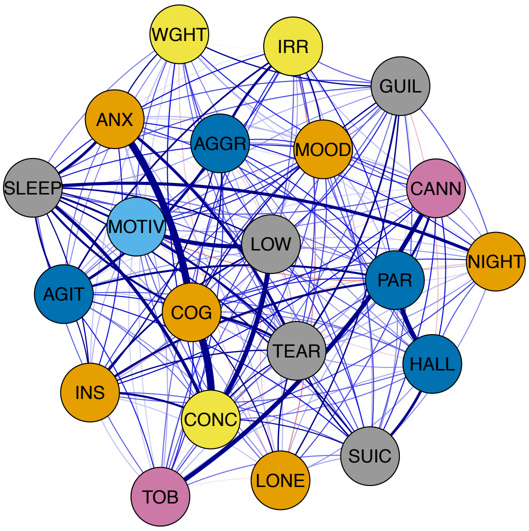

B.

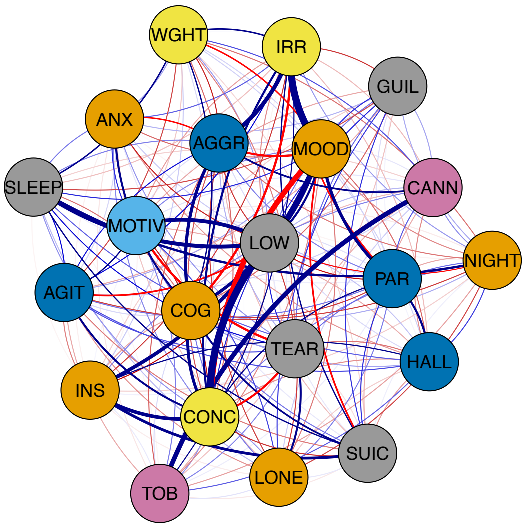

C.

### ii. BMD

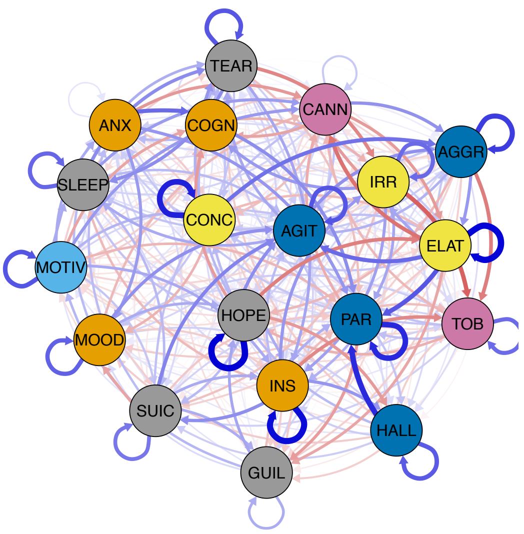
TEAR
CANN
ANX
COGN
AGGR
IRR
SLEEP
CONC
AGIT
ELAT
MOTIV
HOPE
PAR
TOB
MOOD
INS
SUIC
HALL
GUIL

A.

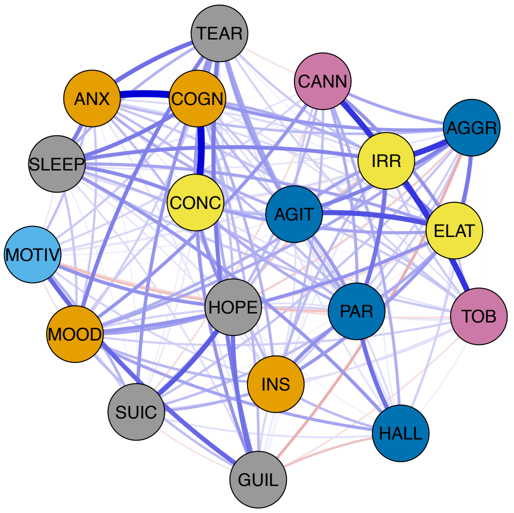
TEAR
CANN
ANX
COGN
AGGR
IRR
SLEEP
CONC
AGIT
ELAT
MOTIV
HOPE
PAR
TOB
MOOD
INS
SUIC
HALL
GUIL

B.

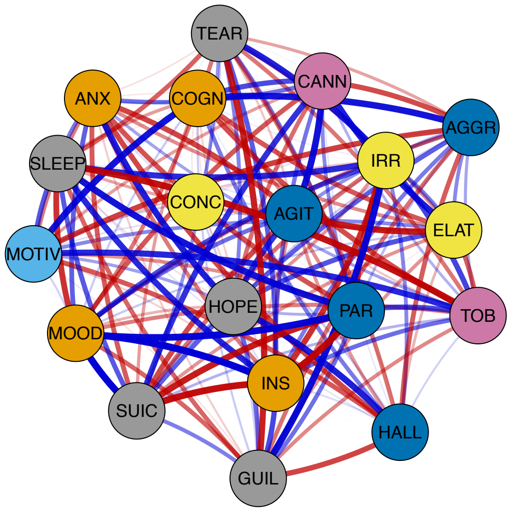
TEAR
CANN
ANX
COGN
AGGR
IRR
SLEEP
CONC
AGIT
ELAT
MOTIV
HOPE
PAR
TOB
MOOD
INS
SUIC
HALL
GUIL

C.

#### iii. PSY

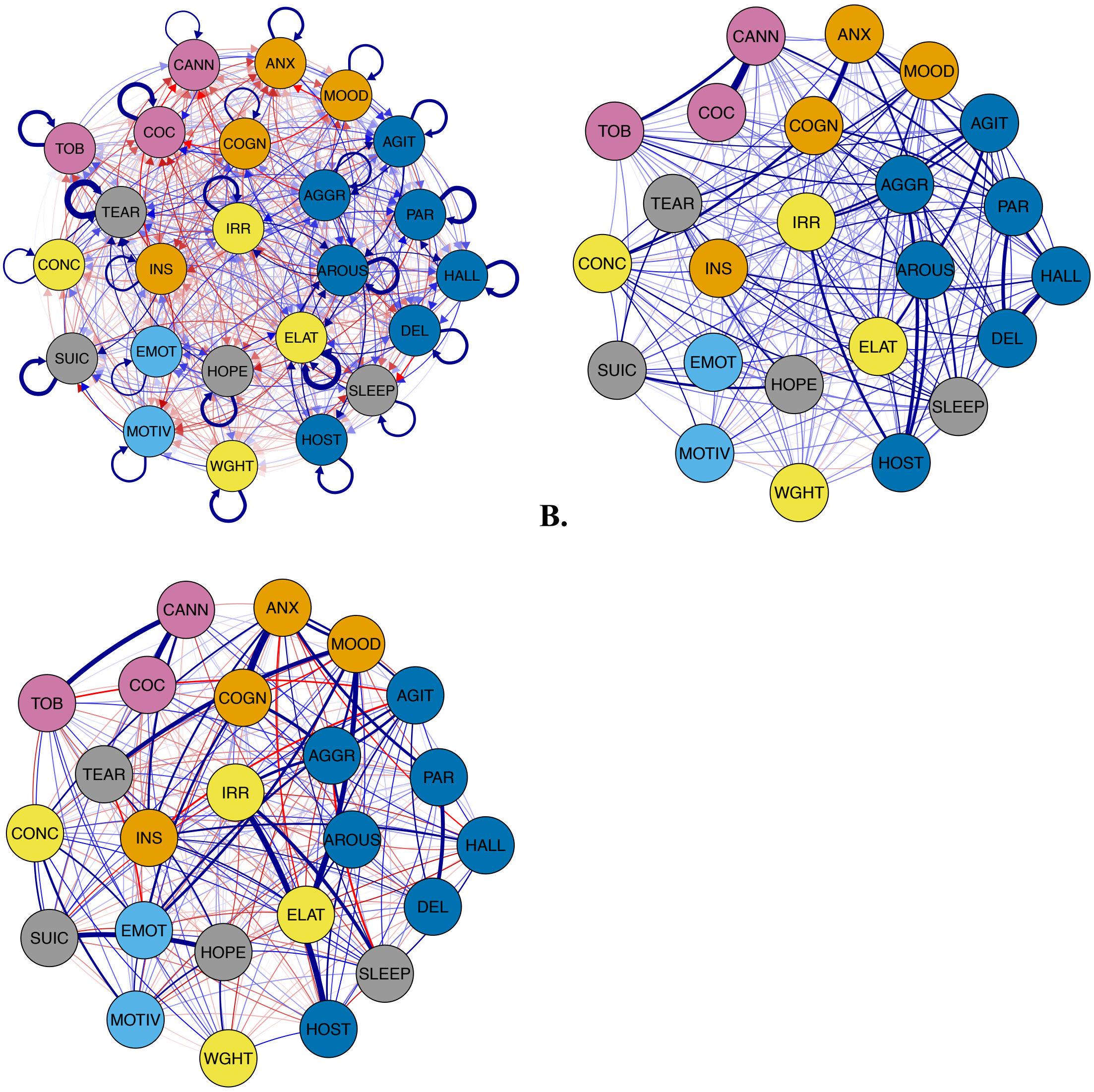
CANN
ANX
MOOD
TOB
COC
COGN
AGIT
TEAR
AGGR
IRR
PAR
CONC
INS
AROUS
HALL
DEL
ELAT
SUIC
EMOT
HOPE
SLEEP
MOTIV
HOST
WGHT

A.

B.

C.

eTable 13 Centrality measures for sub-networks: (A) UMD, (B) BMD and (C) PSY.
For temporal networks, centrality was defined as the sum of absolute (directed) edge weights in (in-centrality, $c_{in}$) and out (out-centrality, $c_{out}$) of a node (including autocorrelative edges). For contemporaneous and between-subject networks, centrality was defined as the sum of absolute (undirected) edge weights for a node (autocorrelative edges do not exist).

**(A) UMD**

| Feature | Temporal (out-centrality) | Temporal (in-centrality) | Contemporaneous | Between-subject |
|---|---|---|---|---|
| Aggression | 0.061 | 0.163 | 0.161 | 0.385 |
| Agitation | 0.026 | 0.065 | 0.083 | 0 |
| Anxiety | 0.032 | 0.027 | 0.359 | 0 |
| Cannabis use | 0.092 | 0.066 | 0.196 | 0.42 |
| Cognitive impairment | 0.027 | 0.06 | 0.456 | 0 |
| Disturbed sleep | 0 | 0 | 0.275 | 0.38 |
| Feeling lonely | 0.06 | 0.109 | 0 | 0 |
| Guilt | 0.094 | 0.037 | 0 | 0 |
| Hallucinations (all) | 0.061 | 0.029 | 0.209 | 0 |
| Irritability | 0.033 | 0.029 | 0.079 | 0.945 |
| Low energy | 0.04 | 0.141 | 0.246 | 1.314 |
| Mood instability | 0.102 | 0.057 | 0 | 1.018 |
| Nightmares | 0.026 | 0 | 0.093 | 0 |
| paranoia | 0.065 | 0.064 | 0.27 | 0 |
| Poor concentration | 0.087 | 0.067 | 0.418 | 1.023 |
| Poor insight | 0 | 0 | 0.071 | 0 |
| Poor motivation | 0.08 | 0.026 | 0.116 | 0.37 |
| Suicidality | 0.061 | 0.042 | 0.082 | 0 |
| Tearfulness | 0.026 | 0.064 | 0.163 | 0 |
| Tobacco use | 0.071 | 0 | 0.124 | 0.42 |
| Weight loss | 0 | 0 | 0 | 0 |

**(B) BMD**

| Feature | Temporal (out-centrality) | Temporal (in-centrality) | Contemporaneous | Between-subject |
|---|---|---|---|---|
| Aggression | 0.107 | 0.165 | 0.317 | 0 |
| Agitation | 0.051 | 0.226 | 0.206 | 0.913 |

| | | | | |
|---|---|---|---|---|
| Anxiety | 0.209 | 0 | 0.366 | 0 |
| Cannabis use | 0.096 | 0.248 | 0.14 | 0.913 |
| Cognitive impairment | 0.1 | 0.16 | 0.44 | 0.904 |
| Disturbed sleep | 0.098 | 0.052 | 0.266 | 1.812 |
| elation | 0.209 | 0.151 | 0.304 | 0 |
| Feeling hopeless | 0.202 | 0 | 0.21 | 0 |
| Guilt | 0 | 0 | 0.292 | 0.929 |
| Hallucinations (all) | 0.098 | 0.095 | 0.098 | 0 |
| irritability | 0.165 | 0.103 | 0.411 | 0.929 |
| Mood instability | 0.05 | 0.046 | 0.087 | 2.893 |
| paranoia | 0 | 0.274 | 0.194 | 2.792 |
| Poor concentration | 0.065 | 0 | 0.173 | 0 |
| Poor insight | 0.202 | 0 | 0 | 2.841 |
| Poor motivation | 0.046 | 0 | 0.105 | 0.904 |
| Suicidality | 0.056 | 0.052 | 0.113 | 1.897 |
| Tearfulness | 0.108 | 0.101 | 0.277 | 0 |
| Tobacco use | 0 | 0.186 | 0.14 | 0.906 |

### (C) PSY

| Feature | Temporal (out-centrality) | Temporal (in-centrality) | Contemporaneous | Between-subject |
|---|---|---|---|---|
| Aggression | 0.072 | 0.035 | 0.335 | 1.178 |
| Agitation | 0.073 | 0.038 | 0.31 | 0.451 |
| Anxiety | 0 | 0.033 | 0.343 | 0.988 |
| Arousal | 0.034 | 0.145 | 0.409 | 0.217 |
| Cannabis use | 0 | 0.031 | 0.47 | 0.984 |
| Cocaine use | 0 | 0.031 | 0.257 | 0.407 |
| Cognitive impairment | 0 | 0 | 0.364 | 0.977 |
| Delusional ideation | 0.033 | 0 | 0.112 | 0.214 |
| Disturbed sleep | 0.063 | 0 | 0.308 | 0.305 |
| Elation | 0.044 | 0.098 | 0.095 | 0.373 |
| Emotional withdrawal | 0 | 0 | 0 | 0.681 |
| Feeling hopeless | 0 | 0 | 0.291 | 0 |
| Hallucinations (all) | 0.072 | 0.035 | 0.335 | 1.178 |
| Hostility | 0.065 | 0.034 | 0.368 | 0.425 |
| Irritability | 0.032 | 0 | 0.383 | 0.96 |

| | | | | |
|---|---|---|---|---|
| Mood instability | 0.064 | 0 | 0 | 1.404 |
| Paranoia | 0 | 0 | 0.524 | 0.55 |
| Poor concentration | 0.033 | 0 | 0.112 | 0.214 |
| Poor insight | 0.031 | 0 | 0 | 0.21 |
| Poor motivation | 0.031 | 0 | 0 | 0.21 |
| Suicidality | 0 | 0 | 0.106 | 0.369 |
| Tearfulness | 0 | 0.033 | 0 | 0.32 |
| Tobacco use | 0 | 0 | 0.126 | 0.367 |
| Weight loss | 0 | 0 | 0 | 0 |

eResults 5 Secondary Analysis (sub-networks)

**a. UMD**

The strongest autocorrelations were observed for hallucinations ($z$=.11), guilt ($z$=.11), tearfulness (z=.10) and cannabis use ($z$=.10) with all the other autocorrelations z<0.10 (Figure 4A).

The most prominent unidirectional relationships were mixed with some positive: poor motivation-low energy ($z_{12}$=.05), guilt-low energy ($z_{12}$=.04), low energy-poor concentration ($z_{12}$=.04), tobacco use-suicidality ($z_{12}$=.04), poor concentration-tearfulness ($z_{12}$=.04); and others negative: poor concentration-loneliness ($z_{12}$=-.05), paranoia-guilt ($z_{12}$=-.04). All other unidirectional and bidirectional relationships were $|z_{12}|$<.04.

**b. BMD**

The strongest autocorrelation was observed for feeling hopeless (z=.12), elation (z=.12), poor insight (z=.11), paranoia (z=.10) and poor concentration (z=.10), with all the other autocorrelations z<0.10 (Figure 4B).

The most prominent unidirectional relationships were mixed, with some positive: hallucinations-paranoia ($z_{12}$=.10), elation-paranoia ($z_{12}$=.08), elation-agitation ($z_{12}$=.07), anxiety-cognitive impairment ($z_{12}$=.07), suicidality-agitation ($z_{12}$=.06), poor concentration-aggression ($z_{12}$=.06); and others negative: irritability-tobacco use ($z_{12}$=-.07). feeling hopeless-elation ($z_{12}$=-.06), aggression-tobacco use ($z_{12}$=-.06), tearfulness-irritability ($z_{12}$=-.06), elation-cannabis use ($z_{12}$=-.06). All other unidirectional and bidirectional relationships were $|z_{12}|$<.06.

**c. PSY**

The strongest autocorrelation was observed for tearfulness ($z$=.13), elation (z=.12) and paranoia (z=0.10) with all the other autocorrelations z<0.10 (Figure 4C).

The most prominent unidirectional relationships were all positive: agitation-arousal ($z_{12}$=.04) and elation-arousal ($z_{12}$=.04). With respect to bidirectional relationships, positively recurring pairs were observed between aggression-agitation ($z_{12}$=.04, $z_{21}$=.04). All other unidirectional and bidirectional relationships were $|z_{12}|$<.04.

eResults 6 Contemporaneous and between-subject results for sub-networks: (A) UMD, (B) BMD and (C) PSY.

(A) UMD

Inspection of associations between residuals (Figure, left), anxiety-cognitive impairment and cognitive impairment-poor concentration had the strongest positive relationship ($z$ = .19), followed by hallucinations (all)-paranoia and low energy-poor concentration ($z$ = .13). None of the negative associations survived thresholding. Within this network, the most central items were cognitive impairment ($c$ = .456), poor concentration ($c$ = .418) and anxiety ($c$ = .359) (eTable 13A).

Between-individuals estimates (Figure, right), suggested that low energy-poor concentration and irritability-mood instability shared the strongest positive relationship ($z$ =.56). The only negative association to survive thresholding was mood instability-poor concentration ($z$ =-.46). The most central items were low energy ($c$ = 1.314), poor concentration ($c$ = 1.023) and mood instability ($c$ = 1.018) (eTable 13A).

See eTable 12A for actual model and bootstrapped estimates.

Graphs represent positive (blue) and negative (red) relationships between nodes from actual model estimates in sub-networks. Edges are displayed as lines, with the thickness representing the strength of the edge weight estimate (partial correlation coefficient, $z$). **(left)** Contemporaneous network (edges thresholded at $|z| > .07$ and labels at $|z| > .08$). **(right)** Between-subject network (edges thresholded at $|z| > .37$ and labels $|z| > .44$).

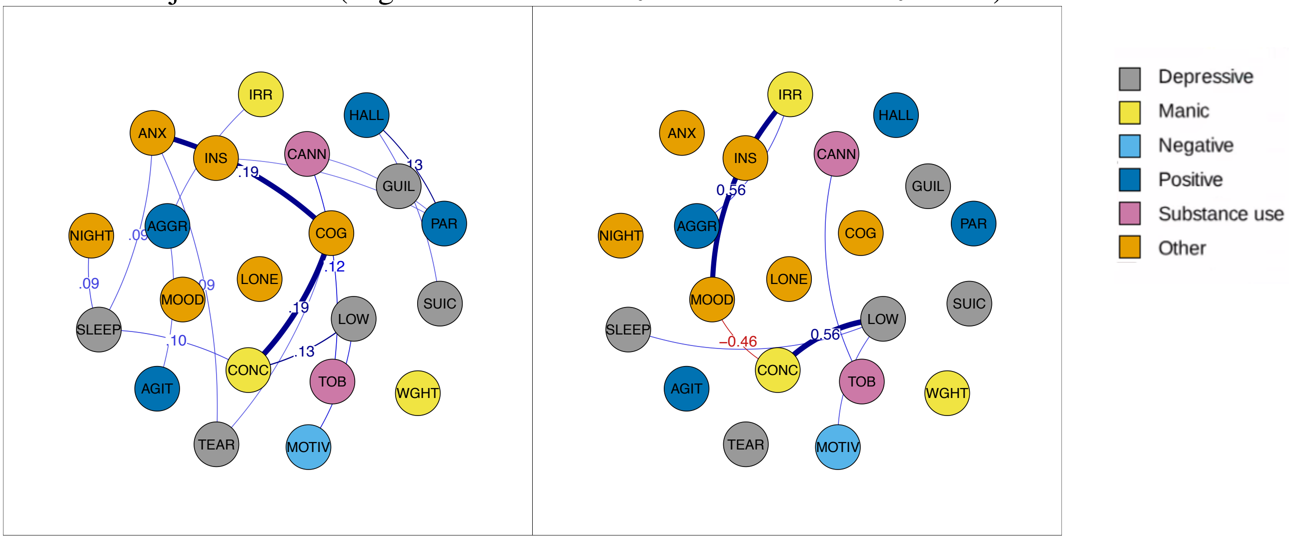

(B) BMD

Inspection of associations between residuals (Figure, left), anxiety-cognitive impairment had the strongest positive relationship ($z$ = .18), followed by cognitive impairment-poor concentration ($z$ = .17), and aggression-irritability ($z$ = .14). None of the negative associations survived thresholding. Within this network, the most central items were cognitive impairment ($c$ = .44), irritability ($c$ = .411), and anxiety ($c$ = .366) (eTable 13B).

Between-individuals estimates (Figure, right) suggested that mood instability-poor insight shared the strongest positive relationship ($z$ = .98), followed by mood instability-suicidality ($z$ =.97), and mood instability-paranoia ($z$ =.95). The strongest negative association was observed between paraonia-poor insight ($z$ =-.94). The most central items were mood instability ($c$ = 2.893), poor insight ($c$ = 2.841) and paranoia ($c$ = 2.792) (eTable 13B).

See eTable 12B for actual model and bootstrapped estimates.

Graphs represent positive (blue) and negative (red) relationships between nodes from actual model estimates in sub-networks. Edges are displayed as lines, with the thickness representing the strength of the edge weight estimate (partial correlation coefficient, $z$). **(left)** Contemporaneous network (edges thresholded at $|z| > .08$ and labels at $|z| > .10$). **(right)** Between-subject network (edges thresholded at $|z| > .9$ and labels $|z| > .93$).

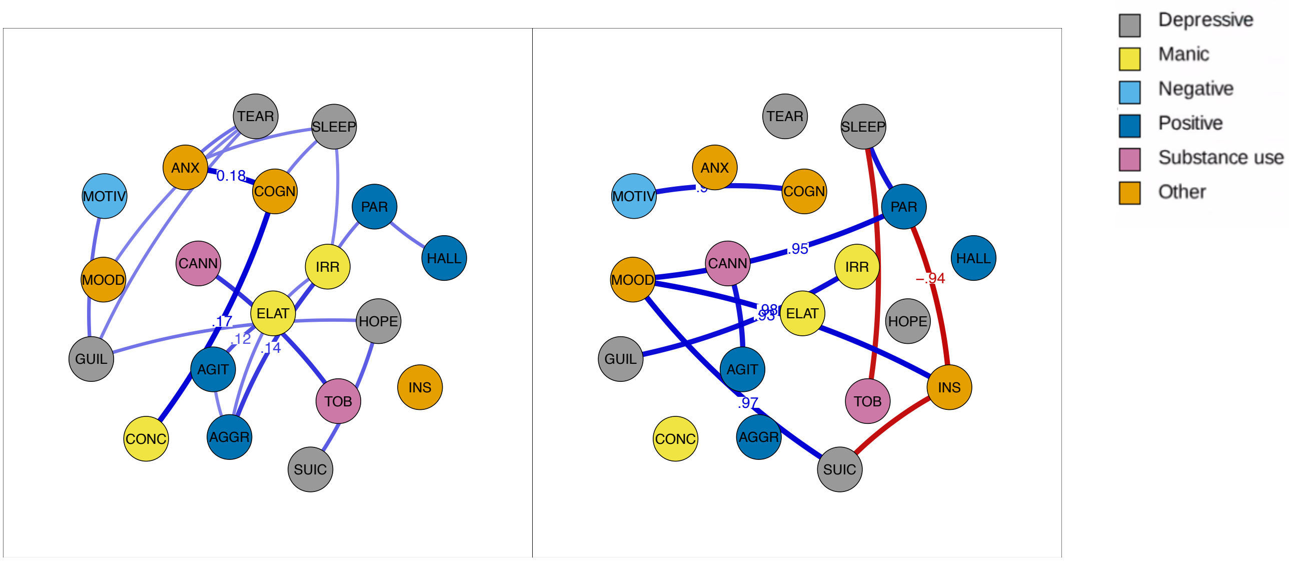

(C) PSY

Inspection of associations between residuals (Figure, left), cannabis use-cocaine use had the strongest positive relationship ($z$ = .26), followed by cognitive impairment-anxiety ($z$ = .17), and delusional thinking-hallucinations (all) ($z$ =.17). None of the negative associations survived thresholding. Within this network, the most central items were paranoia ($c$ = .524), cannabis use ($c$ = .470) and sexual arousal ($c$ = .409) (eTable 13C).

Between-individuals estimates (Figure, right) suggested that anxiety-cognitive impairment shared the strongest positive relationship ($z$ = .49), followed by hostility-irritability ($z$ = .43), and cannabis use-cocaine use ($z$ =.41). None of the negative associations survived thresholding. The most central items were mood instability ($c$ = 1.404), aggression ($c$ = 1.178) and hallucinations (all) ($c$ = .1.178) (eTable 13C).

See eTable 12C for actual model and bootstrapped estimates.

Graphs represent positive (blue) and negative (red) relationships between nodes from actual model estimates in sub-networks. Edges are displayed as lines, with the thickness representing the strength of the edge weight estimate (partial correlation coefficient, $z$). **(left)** Contemporaneous network (edges thresholded at | $z$ | > .08 and labels at | $z$ | > .11). **(right)** Between-subject network (edges thresholded at | $z$ | > .2 and labels | $z$ | > .22).

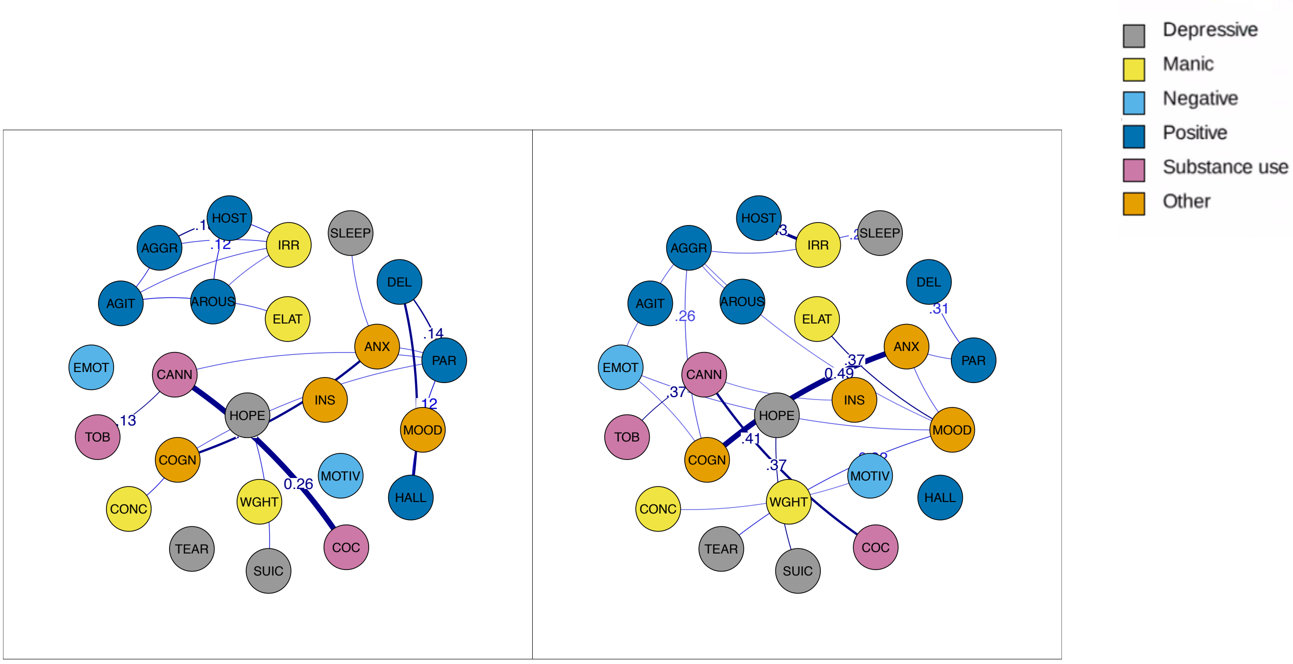

eMethods 7 Permutation analysis node selection

| **UMD (n=21)** | **BMD (n=19)** | **PSY (n=24)** | **Permutation analysis (n=16)** |
|---|---|---|---|
| Aggression | Aggression | Aggression | Aggression |
| Agitation | Agitation | Agitation | Agitation |
| Anxiety | Anxiety | Anxiety | Anxiety |
| Cannabis use | Cannabis use | Arousal | Cannabis use |
| Cognitive impairment | Cognitive impairment | Cannabis use | Cognitive impairment |
| Disturbed sleep | Disturbed sleep | Cocaine use | Disturbed sleep |
| Feeling lonely | Elation | Cognitive impairment | Hallucinations (all) |
| Guilt | Feeling hopeless | Delusional thinking | Irritability |
| Hallucinations (all) | Guilt | Disturbed sleep | Mood instability |
| Irritability | Hallucinations (all) | Elation | Paranoia |
| Low energy | Irritability | Emotional withdrawal | Poor concentration |
| Mood instability | Mood instability | Feeling hopeless | Poor insight |
| Nightmares | Paranoia | Hallucinations (all) | Poor motivation |
| Paranoia | Poor concentration | Hostility | Suicidality |
| Poor concentration | Poor insight | Irritability | Tearfulness |
| Poor insight | Poor motivation | Mood instability | Tobacco use |
| Poor motivation | Suicidality | Paranoia | |
| Suicidality | Tearfulness | Poor concentration | |
| Tearfulness | Tobacco use | Poor insight | |
| Tobacco use | | Poor motivation | |
| Weight loss | | Suicidality | |
| | | Tearfulness | |
| | | Tobacco use | |
| | | Weight loss | |

eTable 14 Permutation edge estimates for sub-networks: (A) UMD, (B) BMD and (C) PSY. For each sub-network, edge estimates are shown for the (i) temporal, (ii, lower triangle) contemporaneous, and (iii, upper triangle) between-subject relationships.

### (A) UMD

#### i. Temporal

| | AGGR | AGIT | ANX | CANN | COGN | CONC | HALL | INS | IRR | MOOD | MOTIV | PAR | SLEEP | SUIC | TEAR | TOB |
|---|---|---|---|---|---|---|---|---|---|---|---|---|---|---|---|---|
| AGGR | 0.066 | 0.032 | -0.001 | 0.008 | -0.006 | -0.004 | -0.003 | 0.018 | -0.012 | 0.03 | 0.002 | 0.016 | -0.005 | 0.004 | -0.011 | -0.008 |
| AGIT | 0.02 | 0.062 | -0.013 | 0.005 | 0.014 | 0.011 | 0.002 | -0.007 | 0.002 | 0.021 | -0.01 | 0.006 | -0.005 | 0.021 | 0.004 | -0.019 |
| ANX | 0.032 | 0.017 | 0.077 | -0.007 | 0.008 | 0.006 | 0.001 | 0.014 | -0.012 | 0.018 | -0.006 | 0.003 | 0.004 | -0.004 | 0.024 | -0.005 |
| CANN | -0.002 | 0.012 | -0.015 | 0.101 | 0.016 | -0.016 | -0.015 | -0.007 | 0.012 | 0.027 | 0 | 0.032 | -0.003 | 0.02 | -0.006 | 0.003 |
| COGN | 0.013 | 0.01 | 0.02 | 0.005 | 0.033 | 0.023 | 0.003 | -0.008 | 0.006 | 0.014 | 0.024 | 0.008 | 0.007 | 0.008 | 0.002 | -0.022 |
| CONC | 0.012 | 0.011 | -0.021 | 0.004 | -0.006 | 0.073 | 0.009 | -0.002 | -0.013 | 0.021 | 0.026 | 0.017 | -0.003 | 0.002 | 0.033 | -0.013 |
| HALL | 0.02 | 0.009 | -0.028 | -0.035 | 0 | 0.015 | 0.107 | -0.012 | 0.019 | 0.026 | 0.004 | 0.003 | -0.004 | 0.005 | 0.024 | -0.009 |
| INS | 0.004 | 0.021 | -0.008 | -0.01 | -0.007 | 0.025 | 0.002 | 0.093 | 0.006 | 0.018 | 0.004 | -0.006 | -0.007 | 0.009 | 0.021 | 0.013 |
| IRR | 0.033 | 0.021 | -0.016 | 0.012 | 0.021 | 0.004 | 0.016 | -0.024 | 0.059 | -0.015 | 0.002 | -0.004 | -0.014 | 0.015 | 0.005 | -0.003 |
| MOOD | 0.045 | 0.014 | -0.009 | -0.006 | 0.019 | 0.022 | 0.008 | 0.008 | -0.02 | 0.071 | -0.001 | -0.02 | -0.006 | 0.009 | 0.03 | 0.006 |
| MOTIV | 0.001 | -0.033 | -0.008 | 0.019 | 0.02 | 0.017 | 0.009 | 0.007 | 0.005 | 0.017 | 0.051 | -0.004 | -0.012 | -0.002 | 0.016 | -0.002 |
| PAR | 0.001 | 0.003 | -0.004 | 0.015 | -0.012 | 0.022 | 0.024 | -0.02 | 0.028 | 0.008 | 0.009 | 0.07 | -0.011 | -0.022 | 0 | 0.005 |
| SLEEP | -0.003 | 0.015 | -0.015 | 0.011 | -0.005 | 0.006 | 0.005 | 0.016 | -0.008 | -0.005 | 0.02 | 0.006 | 0.079 | -0.003 | 0.024 | 0 |
| SUIC | -0.018 | 0.012 | -0.001 | -0.008 | -0.009 | 0.006 | 0.029 | -0.018 | -0.005 | -0.015 | 0.017 | -0.032 | -0.01 | 0.068 | 0.016 | 0.007 |
| TEAR | 0.006 | -0.001 | 0.009 | 0.001 | -0.001 | 0.026 | 0.019 | 0.02 | 0.009 | -0.009 | 0.027 | -0.006 | -0.008 | 0.009 | 0.104 | -0.022 |
| TOB | 0.028 | 0.001 | -0.019 | 0.004 | 0.012 | -0.021 | 0.009 | -0.017 | 0.003 | 0.009 | 0.012 | -0.002 | -0.003 | 0.042 | 0.003 | 0.068 |

### ii. Contemporaneous (lower triangle) / Between-subject (upper triangle)

| | AGGR | AGIT | ANX | CANN | COGN | CONC | HALL | INS | IRR | MOOD | MOTIV | PAR | SLEEP | SUIC | TEAR | TOB |
|---|---|---|---|---|---|---|---|---|---|---|---|---|---|---|---|---|
| **AGGR** | NA | 0.331 | -0.236 | 0.211 | 0.323 | -0.329 | -0.006 | 0.064 | 0.407 | -0.316 | -0.059 | 0.141 | 0.014 | -0.086 | 0 | -0.129 |
| **AGIT** | 0.084 | NA | 0.072 | -0.025 | 0.007 | 0.16 | 0.13 | -0.058 | 0.205 | 0.067 | 0.172 | 0.162 | 0.067 | -0.136 | 0.097 | -0.043 |
| **ANX** | 0.029 | 0.028 | NA | -0.001 | 0.317 | -0.032 | -0.1 | 0.041 | 0.185 | -0.22 | 0.104 | 0.091 | 0.089 | 0.006 | 0.045 | -0.007 |
| **CANN** | 0.021 | 0.009 | 0.012 | NA | -0.033 | -0.069 | -0.079 | 0.098 | -0.03 | -0.001 | 0.048 | 0.146 | 0.029 | -0.056 | -0.038 | 0.42 |
| **COGN** | 0.051 | 0.012 | 0.193 | 0.045 | NA | 0.35 | 0.103 | -0.072 | -0.251 | 0.294 | -0.083 | -0.156 | -0.093 | 0.254 | 0.075 | 0.049 |
| **CONC** | 0.004 | 0.03 | 0.013 | 0.005 | 0.201 | NA | -0.108 | 0.31 | 0.255 | -0.383 | 0.014 | 0.165 | 0.403 | -0.169 | -0.159 | 0.042 |
| **HALL** | 0.035 | 0.028 | 0.002 | 0.045 | 0.026 | 0.031 | NA | 0.227 | -0.017 | -0.173 | -0.08 | 0.279 | 0.242 | -0.095 | -0.125 | 0.119 |
| **INS** | 0.051 | 0.046 | 0.043 | 0.045 | 0.019 | 0.079 | 0.03 | NA | -0.164 | 0.344 | -0.098 | 0.032 | -0.111 | 0.324 | -0.034 | 0.038 |
| **IRR** | 0.079 | 0.059 | 0.005 | 0.011 | 0.04 | 0.032 | 0.029 | 0.001 | NA | 0.531 | -0.129 | -0.256 | -0.033 | 0.219 | -0.059 | 0.236 |
| **MOOD** | 0.039 | 0.029 | 0.017 | 0.038 | 0.04 | 0.038 | 0.025 | 0.037 | 0.051 | NA | 0.228 | 0.308 | 0.281 | -0.263 | 0.067 | -0.136 |
| **MOTIV** | 0.001 | 0.017 | 0.015 | 0.024 | 0.047 | 0.045 | -0.005 | 0.024 | 0.043 | 0.047 | NA | -0.025 | 0.154 | 0.13 | -0.23 | 0.098 |
| **PAR** | 0.044 | 0.068 | 0.039 | 0.072 | 0.041 | 0.039 | 0.129 | 0.073 | 0.044 | 0.015 | -0.017 | NA | -0.102 | 0.19 | -0.009 | 0.033 |
| **SLEEP** | 0.021 | 0.063 | 0.093 | 0.034 | 0.079 | 0.112 | 0.036 | 0.052 | 0.047 | 0.046 | 0.06 | 0.028 | NA | 0.264 | 0.309 | -0.037 |
| **SUIC** | 0.037 | 0.047 | 0.034 | 0.044 | 0.024 | 0.054 | 0.084 | 0.032 | 0.014 | 0.038 | 0.05 | 0.022 | 0.06 | NA | 0.014 | -0.031 |
| **TEAR** | -0.004 | 0.046 | 0.092 | 0.021 | 0.086 | 0.056 | 0.02 | 0.053 | 0.036 | 0.037 | 0.039 | 0.021 | 0.075 | 0.061 | NA | 0.078 |
| **TOB** | 0.039 | 0.042 | 0.017 | 0.126 | 0.041 | -0.001 | 0.015 | 0.012 | 0.027 | 0.036 | 0.012 | 0.034 | 0.031 | 0.027 | 0.023 | NA |

### (B) BMD

### i. Temporal

| | AGGR | AGIT | ANX | CANN | COGN | CONC | HALL | INS | IRR | MOOD | MOTIV | PAR | SLEEP | SUIC | TEAR | TOB |
|---|---|---|---|---|---|---|---|---|---|---|---|---|---|---|---|---|
| **AGGR** | 0.096 | 0.039 | 0.003 | -0.011 | 0.008 | 0.023 | 0.018 | -0.005 | -0.004 | 0.008 | -0.008 | 0.01 | 0.017 | -0.025 | -0.016 | -0.056 |
| **AGIT** | 0.052 | 0.084 | -0.009 | -0.015 | -0.006 | -0.008 | 0.006 | 0.02 | -0.016 | -0.002 | -0.007 | 0.025 | 0.043 | -0.008 | 0.043 | 0.002 |
| **ANX** | 0.007 | 0 | 0.012 | -0.049 | 0.067 | -0.027 | 0.009 | 0.004 | -0.004 | 0.045 | -0.01 | 0.048 | -0.017 | 0.016 | 0.034 | 0.007 |
| **CANN** | 0.046 | 0.023 | 0.016 | 0.023 | -0.039 | -0.031 | -0.009 | -0.008 | -0.006 | 0.016 | -0.011 | 0.013 | 0.017 | -0.009 | 0.006 | -0.021 |
| **COGN** | 0.01 | 0.026 | 0.033 | -0.022 | 0.019 | 0.027 | 0.002 | 0.007 | -0.034 | 0.015 | -0.023 | 0.053 | 0.012 | 0.014 | 0.049 | -0.006 |
| **CONC** | 0.062 | 0.024 | 0.022 | -0.003 | -0.034 | 0.099 | 0.017 | 0.011 | -0.018 | -0.039 | -0.013 | 0.005 | 0.005 | -0.012 | 0.032 | 0.015 |
| **HALL** | 0.02 | -0.006 | -0.009 | -0.011 | -0.003 | 0.031 | 0.078 | 0.024 | -0.008 | -0.022 | 0.009 | 0.098 | -0.012 | 0.025 | -0.025 | -0.021 |
| **INS** | 0.019 | 0.031 | 0.018 | 0.006 | -0.007 | 0.014 | 0.048 | 0.111 | 0.046 | -0.004 | -0.007 | 0.037 | -0.033 | 0.053 | 0.011 | -0.054 |
| **IRR** | 0.03 | -0.001 | 0.009 | -0.044 | 0.046 | -0.013 | 0.02 | 0.046 | 0.07 | 0.01 | -0.021 | 0.042 | 0.028 | -0.028 | 0.007 | -0.07 |
| **MOOD** | 0.011 | 0.049 | 0.031 | -0.031 | 0.002 | 0.01 | 0.022 | -0.023 | -0.005 | 0.074 | -0.014 | 0.021 | -0.001 | -0.018 | -0.005 | 0.022 |
| **MOTIV** | 0.012 | 0.017 | 0.04 | 0.046 | -0.044 | 0.035 | -0.012 | 0.038 | -0.012 | -0.006 | 0.081 | -0.031 | 0.037 | 0.024 | -0.024 | -0.004 |
| **PAR** | 0.013 | 0.028 | 0.017 | -0.039 | 0.003 | -0.009 | 0.03 | 0.037 | -0.011 | 0.005 | -0.012 | 0.097 | 0.016 | 0 | 0.018 | -0.027 |
| **SLEEP** | 0.008 | 0.035 | 0.022 | 0.046 | 0 | 0.019 | 0.008 | 0.001 | -0.007 | 0.004 | 0.019 | 0.014 | 0.072 | 0.034 | 0.053 | -0.014 |
| **SUIC** | 0.006 | 0.059 | 0.032 | 0.028 | -0.008 | -0.009 | 0.007 | 0.042 | -0.002 | -0.041 | 0.016 | -0.024 | 0.003 | 0.065 | 0.009 | 0.018 |
| **TEAR** | -0.002 | 0.024 | 0.02 | 0.008 | 0.001 | -0.007 | -0.007 | 0.042 | -0.057 | 0.027 | -0.021 | -0.029 | 0.053 | 0.009 | 0.081 | 0.004 |
| **TOB** | 0.018 | 0.032 | 0.027 | -0.036 | -0.015 | -0.018 | -0.004 | -0.004 | -0.007 | 0.01 | -0.023 | 0.03 | -0.018 | 0.017 | -0.002 | 0.071 |

### ii. Contemporaneous (lower triangle) / Between-subject (upper triangle)

| | AGGR | AGIT | ANX | CANN | COGN | CONC | HALL | INS | IRR | MOOD | MOTIV | PAR | SLEEP | SUIC | TEAR | TOB |
|---|---|---|---|---|---|---|---|---|---|---|---|---|---|---|---|---|
| AGGR | NA | 0.684 | -0.059 | -0.663 | 0.78 | -0.68 | -0.466 | 0.717 | 0.575 | -0.479 | -0.319 | 0.382 | -0.263 | 0.687 | -0.331 | -0.157 |
| AGIT | 0.098 | NA | 0.188 | 0.33 | -0.663 | 0.732 | 0.427 | -0.401 | -0.147 | 0.087 | 0.444 | -0.032 | -0.12 | -0.298 | 0.124 | -0.111 |
| ANX | 0.051 | 0.059 | NA | 0.201 | 0.334 | -0.115 | -0.174 | -0.274 | -0.305 | 0.349 | -0.412 | -0.33 | 0.438 | -0.256 | 0.134 | 0.361 |
| CANN | 0.063 | 0.044 | 0.001 | NA | 0.429 | -0.387 | -0.102 | 0.884 | 0.862 | -0.831 | 0.234 | 0.784 | -0.711 | 0.88 | -0.499 | -0.592 |
| COGN | 0.069 | 0.052 | 0.177 | -0.001 | NA | 0.74 | 0.567 | -0.451 | -0.233 | 0.147 | 0.61 | -0.031 | -0.105 | -0.399 | 0.16 | -0.172 |
| CONC | 0.024 | 0.056 | 0.017 | 0.013 | 0.181 | NA | -0.44 | 0.451 | 0.23 | -0.149 | -0.369 | 0.095 | 0.075 | 0.371 | -0.007 | 0.058 |
| HALL | 0.037 | 0.027 | 0.002 | 0.056 | 0.021 | -0.001 | NA | 0.135 | -0.026 | 0.097 | -0.584 | -0.092 | 0.287 | 0.126 | -0.038 | 0.354 |
| INS | 0.038 | 0.032 | 0.046 | 0.052 | 0.058 | 0.04 | 0.047 | NA | -0.929 | 0.902 | -0.27 | -0.809 | 0.768 | -0.962 | 0.531 | 0.693 |
| IRR | 0.15 | 0.076 | 0.03 | 0.038 | 0.056 | -0.01 | 0.014 | 0.053 | NA | 0.98 | -0.488 | -0.906 | 0.903 | -0.96 | 0.564 | 0.84 |
| MOOD | 0.06 | 0.017 | 0.039 | 0.084 | 0.042 | 0.047 | 0.063 | 0.059 | 0.06 | NA | 0.578 | 0.92 | -0.929 | 0.937 | -0.536 | -0.878 |
| MOTIV | 0.013 | 0.011 | 0.046 | 0.037 | 0.045 | 0.045 | -0.002 | 0.037 | -0.01 | -0.011 | NA | -0.637 | 0.739 | -0.31 | 0.219 | 0.81 |
| PAR | 0.04 | 0.077 | 0.048 | 0.063 | 0.04 | 0.065 | 0.101 | 0.085 | 0.102 | 0.037 | -0.04 | NA | 0.9 | -0.837 | 0.528 | 0.866 |
| SLEEP | 0.023 | 0.033 | 0.093 | 0.024 | 0.096 | 0.087 | 0.072 | 0.023 | 0.09 | 0.048 | 0.018 | 0.054 | NA | 0.806 | -0.499 | -0.911 |
| SUIC | 0.036 | 0.045 | 0.008 | -0.021 | 0.017 | 0.022 | 0.023 | 0.06 | -0.005 | 0.036 | 0.059 | 0.047 | 0.013 | NA | 0.569 | 0.723 |
| TEAR | -0.027 | 0.073 | 0.1 | 0.001 | 0.069 | 0.082 | 0.017 | 0.031 | 0.037 | 0.099 | 0.019 | 0.062 | 0.058 | 0.08 | NA | -0.452 |
| TOB | 0.007 | 0.042 | 0.042 | 0.141 | 0.021 | 0.021 | 0.013 | 0.024 | 0.041 | 0.033 | -0.03 | 0.021 | 0.042 | 0.019 | 0.027 | NA |

### (C) PSY

### i. Temporal

| | AGGR | AGIT | ANX | CANN | COGN | CONC | HALL | INS | IRR | MOOD | MOTIV | PAR | SLEEP | SUIC | TEAR | TOB |
|---|---|---|---|---|---|---|---|---|---|---|---|---|---|---|---|---|
| **AGGR** | 0.065 | 0.045 | -0.023 | -0.028 | 0.009 | -0.008 | -0.006 | -0.006 | 0.008 | -0.014 | -0.01 | 0.03 | -0.011 | -0.007 | 0.023 | 0.016 |
| **AGIT** | 0.037 | 0.073 | -0.001 | -0.021 | 0 | -0.001 | 0.017 | 0.003 | 0.019 | -0.003 | -0.011 | 0 | -0.019 | 0 | 0.012 | 0.02 |
| **ANX** | 0.017 | 0.005 | 0.065 | -0.015 | 0.002 | -0.003 | 0.015 | -0.002 | -0.005 | -0.007 | -0.002 | 0.01 | 0.015 | -0.006 | 0.02 | 0.009 |
| **CANN** | 0.015 | 0.027 | -0.012 | 0.045 | -0.008 | -0.005 | 0.003 | -0.02 | -0.003 | 0.01 | 0.002 | 0.008 | 0.002 | -0.007 | 0.002 | 0.008 |
| **COGN** | 0.007 | 0.017 | 0 | -0.003 | 0.051 | -0.001 | 0.006 | 0 | 0.003 | 0.006 | -0.01 | 0.012 | 0.02 | 0.003 | 0.029 | -0.003 |
| **CONC** | 0.009 | 0.021 | -0.012 | -0.007 | 0.002 | 0.042 | -0.009 | 0.002 | 0.002 | 0.006 | -0.01 | 0.017 | 0.014 | 0.01 | 0.033 | 0 |
| **HALL** | 0.002 | 0.012 | 0.003 | -0.015 | -0.004 | 0.001 | 0.087 | -0.013 | -0.016 | -0.015 | 0 | 0.033 | 0.025 | 0.007 | 0.016 | -0.003 |
| **INS** | 0.02 | -0.01 | -0.023 | -0.03 | -0.007 | 0.004 | 0.004 | 0.056 | 0.009 | 0.006 | -0.006 | 0.018 | 0.002 | 0.03 | 0.029 | 0.014 |
| **IRR** | 0.013 | 0.032 | -0.019 | -0.009 | 0.021 | -0.012 | 0.001 | -0.02 | 0.08 | 0.003 | -0.006 | 0.006 | -0.002 | 0.007 | 0.008 | 0.018 |
| **MOOD** | -0.005 | 0.023 | -0.033 | -0.011 | 0.008 | 0.013 | -0.009 | -0.005 | 0.029 | 0.058 | 0 | 0.019 | 0.003 | 0.009 | 0.02 | -0.006 |
| **MOTIV** | -0.013 | -0.005 | 0.006 | -0.003 | 0.011 | -0.001 | 0.019 | 0.014 | 0.014 | 0.006 | 0.056 | 0 | -0.004 | -0.026 | 0.009 | -0.005 |
| **PAR** | 0.013 | 0.015 | -0.009 | -0.015 | 0.016 | 0.007 | 0.015 | -0.007 | 0.002 | 0.004 | -0.015 | 0.101 | 0.01 | 0 | 0.008 | -0.004 |
| **SLEEP** | 0.025 | -0.003 | 0 | 0.003 | -0.009 | 0.008 | 0.004 | 0.003 | -0.009 | 0.009 | -0.01 | 0.011 | 0.061 | -0.015 | 0.008 | 0.011 |
| **SUIC** | 0.003 | -0.004 | -0.007 | -0.013 | -0.014 | -0.007 | 0.005 | 0.005 | 0.018 | -0.008 | 0.008 | 0 | -0.02 | 0.093 | 0.021 | 0.001 |
| **TEAR** | 0.003 | 0.013 | 0.007 | -0.022 | -0.007 | 0.013 | -0.003 | 0.009 | 0.006 | 0.011 | -0.026 | 0.011 | -0.017 | -0.007 | 0.13 | -0.02 |
| **TOB** | 0.012 | 0.008 | 0.014 | 0.011 | -0.004 | -0.004 | 0.005 | -0.015 | -0.005 | -0.001 | -0.013 | -0.003 | 0 | 0.006 | 0.011 | 0.084 |

### ii. Contemporaneous (lower triangle) / Between (upper triangle)

| | AGGR | AGIT | ANX | CANN | COGN | CONC | HALL | INS | IRR | MOOD | MOTIV | PAR | SLEEP | SUIC | TEAR | TOB |
|---|---|---|---|---|---|---|---|---|---|---|---|---|---|---|---|---|
| **AGGR** | NA | 0.276 | -0.102 | 0.136 | 0.237 | -0.027 | 0.018 | -0.014 | 0.369 | 0.218 | -0.09 | 0.059 | -0.226 | 0.08 | -0.078 | 0.11 |
| **AGIT** | 0.137 | NA | 0.143 | 0.046 | 0.048 | 0.018 | 0.112 | -0.157 | 0.259 | 0.018 | -0.039 | 0.109 | 0.106 | 0.062 | -0.066 | -0.158 |
| **ANX** | 0.03 | 0.083 | NA | -0.042 | 0.488 | 0.159 | -0.137 | 0.176 | -0.144 | 0.191 | -0.105 | 0.235 | 0.177 | -0.024 | 0.097 | 0.005 |
| **CANN** | 0.044 | 0.041 | 0.036 | NA | -0.032 | -0.046 | 0.01 | 0.207 | 0.004 | -0.012 | 0.048 | 0.169 | -0.117 | -0.027 | 0.077 | 0.376 |
| **COGN** | 0.07 | 0.038 | 0.17 | 0.046 | NA | 0.122 | 0.152 | 0.007 | 0.088 | -0.133 | 0.178 | -0.025 | -0.029 | 0.008 | 0.06 | 0.055 |
| **CONC** | -0.006 | 0.038 | 0.021 | 0.011 | 0.115 | NA | 0.11 | 0.049 | 0.009 | -0.079 | 0.254 | -0.073 | 0.116 | 0.152 | 0.035 | -0.072 |
| **HALL** | 0.062 | 0.074 | 0.042 | 0.055 | 0.063 | 0.049 | NA | 0.191 | -0.19 | -0.013 | -0.126 | 0.08 | 0.161 | 0.006 | 0.008 | 0.064 |
| **INS** | 0.033 | 0.038 | 0.03 | 0.056 | 0.022 | 0.059 | 0.091 | NA | 0.055 | 0.186 | 0.04 | 0.087 | 0.149 | 0.114 | -0.158 | -0.083 |
| **IRR** | 0.137 | 0.125 | 0.016 | 0.05 | 0.048 | 0.046 | 0.025 | 0.024 | NA | 0.118 | -0.031 | 0.096 | 0.28 | -0.247 | 0.085 | 0.057 |
| **MOOD** | 0.056 | 0.045 | 0.01 | 0.039 | 0.022 | 0.071 | 0.013 | 0.032 | 0.089 | NA | 0.102 | -0.099 | 0.01 | 0.11 | 0.287 | 0.079 |
| **MOTIV** | 0.011 | -0.011 | 0.009 | 0.027 | 0.056 | 0.068 | 0.047 | 0.025 | -0.018 | 0.031 | NA | 0.049 | 0.074 | -0.006 | 0.017 | 0.109 |
| **PAR** | 0.088 | 0.074 | 0.089 | 0.104 | 0.097 | 0.01 | 0.156 | 0.076 | 0.046 | 0.048 | 0.02 | NA | 0.042 | 0.059 | -0.13 | 0.017 |
| **SLEEP** | 0.026 | 0.065 | 0.092 | 0.012 | 0.066 | 0.077 | 0.057 | 0.069 | 0.054 | 0.036 | 0.035 | 0.057 | NA | 0.193 | 0.109 | 0.094 |
| **SUIC** | 0.028 | 0.051 | 0.003 | 0.029 | 0.006 | 0.056 | 0.062 | 0.052 | 0.005 | 0.028 | 0.043 | 0.02 | 0.041 | NA | 0.19 | 0.11 |
| **TEAR** | 0.004 | 0.064 | 0.053 | -0.007 | 0.036 | 0.067 | 0.019 | 0.038 | 0.047 | 0.091 | 0.002 | 0.023 | 0.072 | 0.086 | NA | -0.124 |
| **TOB** | 0.088 | 0.071 | 0.023 | 0.141 | 0.029 | 0.027 | 0.033 | 0.041 | 0.048 | 0.026 | 0.005 | 0.034 | 0.054 | 0.035 | 0.018 | NA |

eFigure 6 Histograms of null distribution of permuted differences for significant pairwise edge comparisons (UMD-BMD, BMD-PSY, UMD-PSY) in temporal sub-networks. Histograms representing the distribution of differences calculated from 250 permutations under the null hypothesis. The vertical blue line marks the critical threshold for a 0.05 significance level. The red line indicates the observed difference from the actual model, highlighting the extent of deviation from the null expectation. Differences beyond the blue line are considered statistically significant.

(a) BMD-PSY

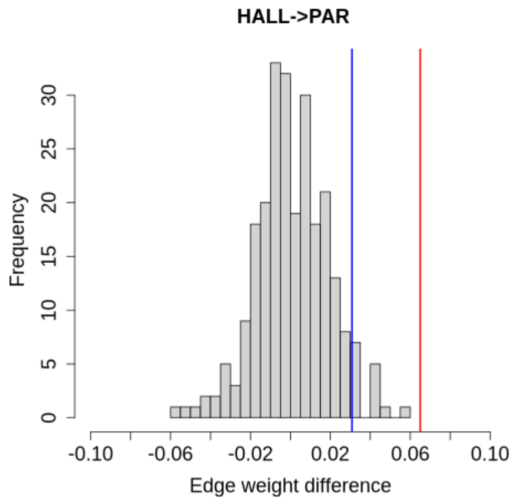

(b) UMD- BMD

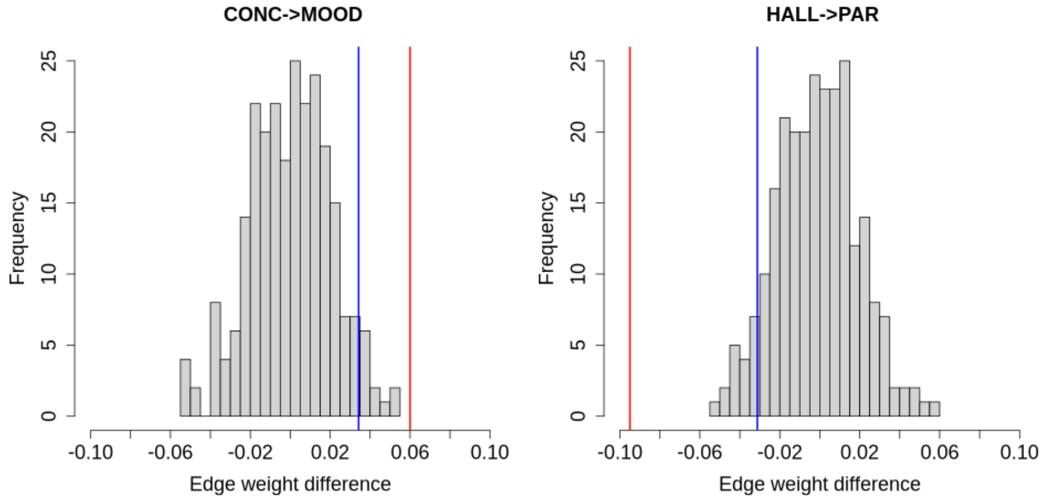

(c) UMD-PSY

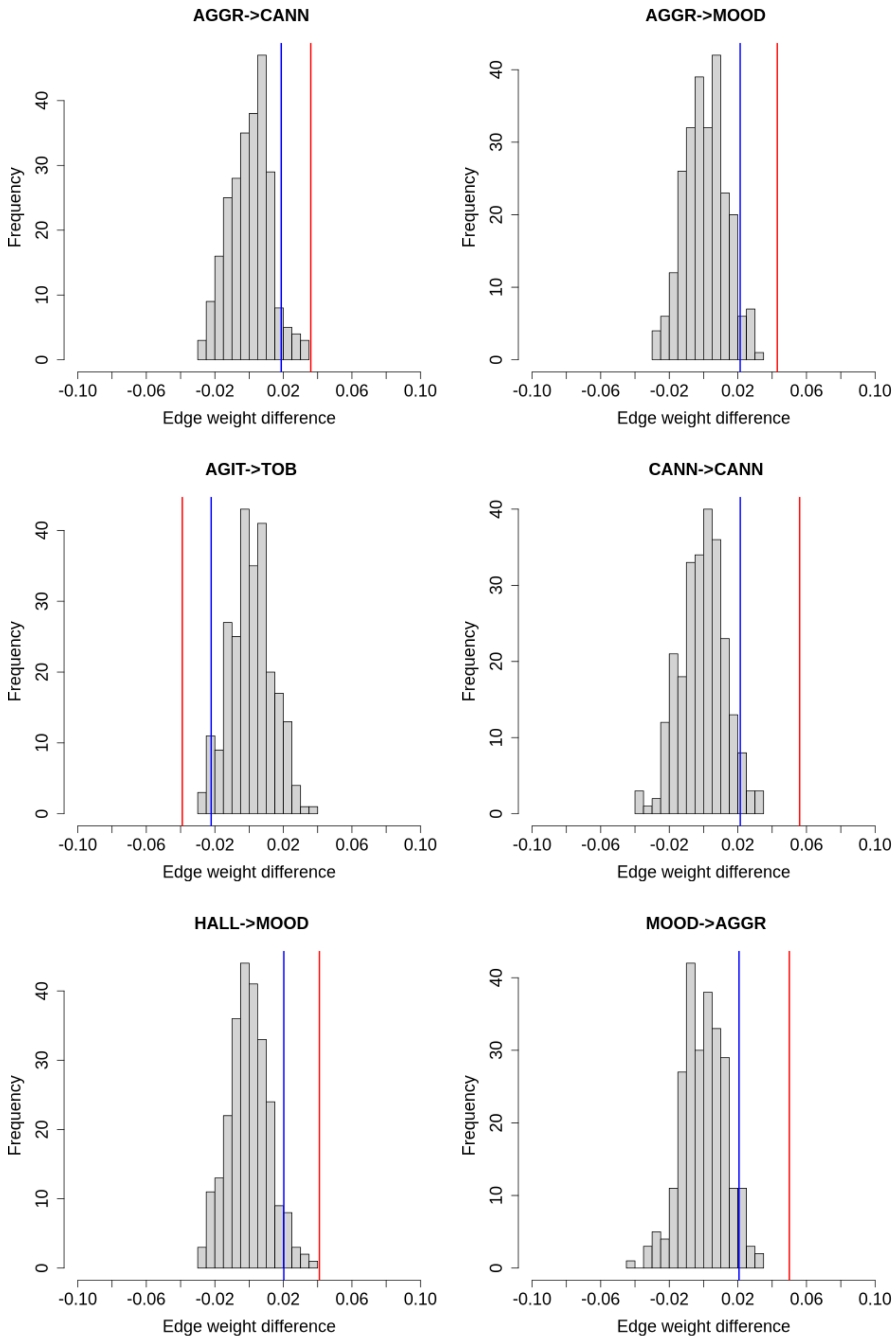
AGGR->CANN
AGGR->MOOD
AGIT->TOB
CANN->CANN
HALL->MOOD
MOOD->AGGR
Frequency
Edge weight difference
0
10
20
30
40
-0.10
-0.06
-0.02
0.02
0.06
0.10

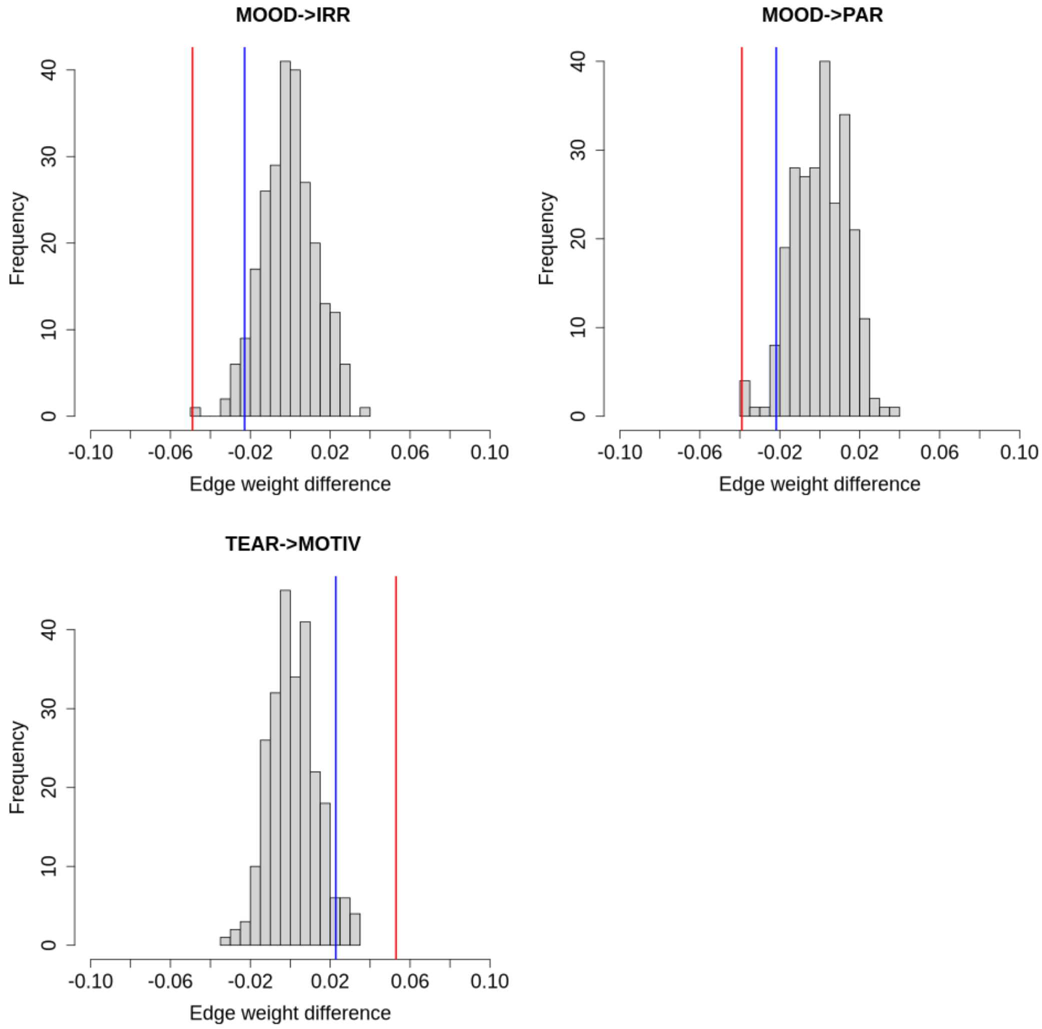
MOOD->IRR
MOOD->PAR
TEAR->MOTIV
Frequency
0
10
20
30
40
-0.10
-0.06
-0.02
0.02
0.06
0.10
Edge weight difference

eFigure 7 Heat-maps for pairwise edge comparisons (UMD-BMD, BMD-PSY, UMD-PSY) in contemporaneous sub-networks in permutation analysis. Magnitude and direction of effect size is colour-coded such that for the pairwise comparison Group1-Group2, yellow indicates the edge estimate is more positive in Group1>Group2 and blue indicates the opposite Group1<Group2. Significant pairwise comparisons (corrected p<0.05) are marked with an asterisk (*).

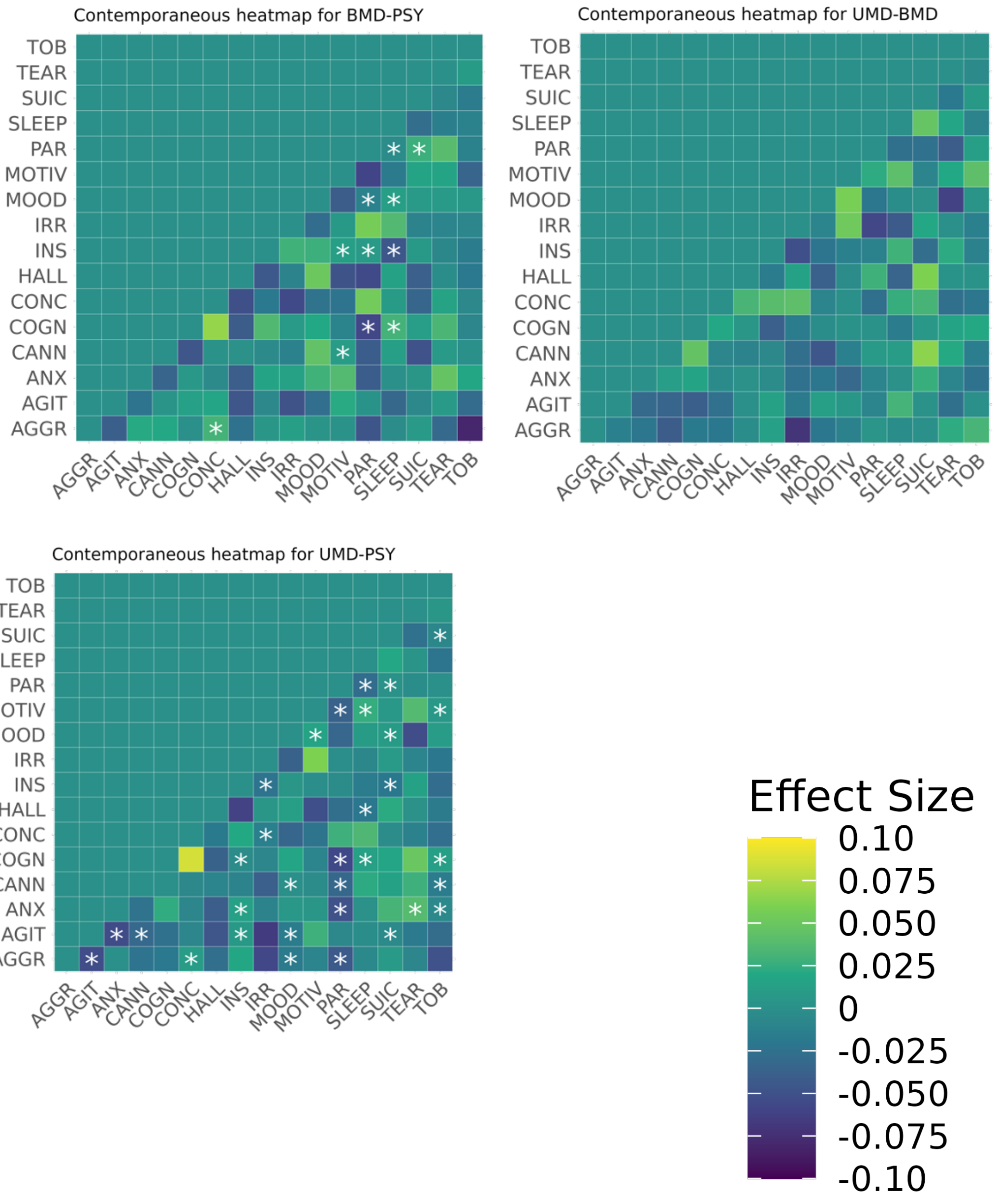

eFigure 8 Heat-maps for pairwise edge comparisons (UMD-BMD, BMD-PSY, UMD-PSY) in between-subject sub-networks in permutation analysis. Magnitude and direction of effect size is colour-coded such that for the pairwise comparison Group1-Group2, yellow indicates the edge estimate is more positive in Group1>Group2 and blue indicates the opposite Group1<Group2. Significant pairwise comparisons (corrected p<0.05) are marked with an asterisk (*).

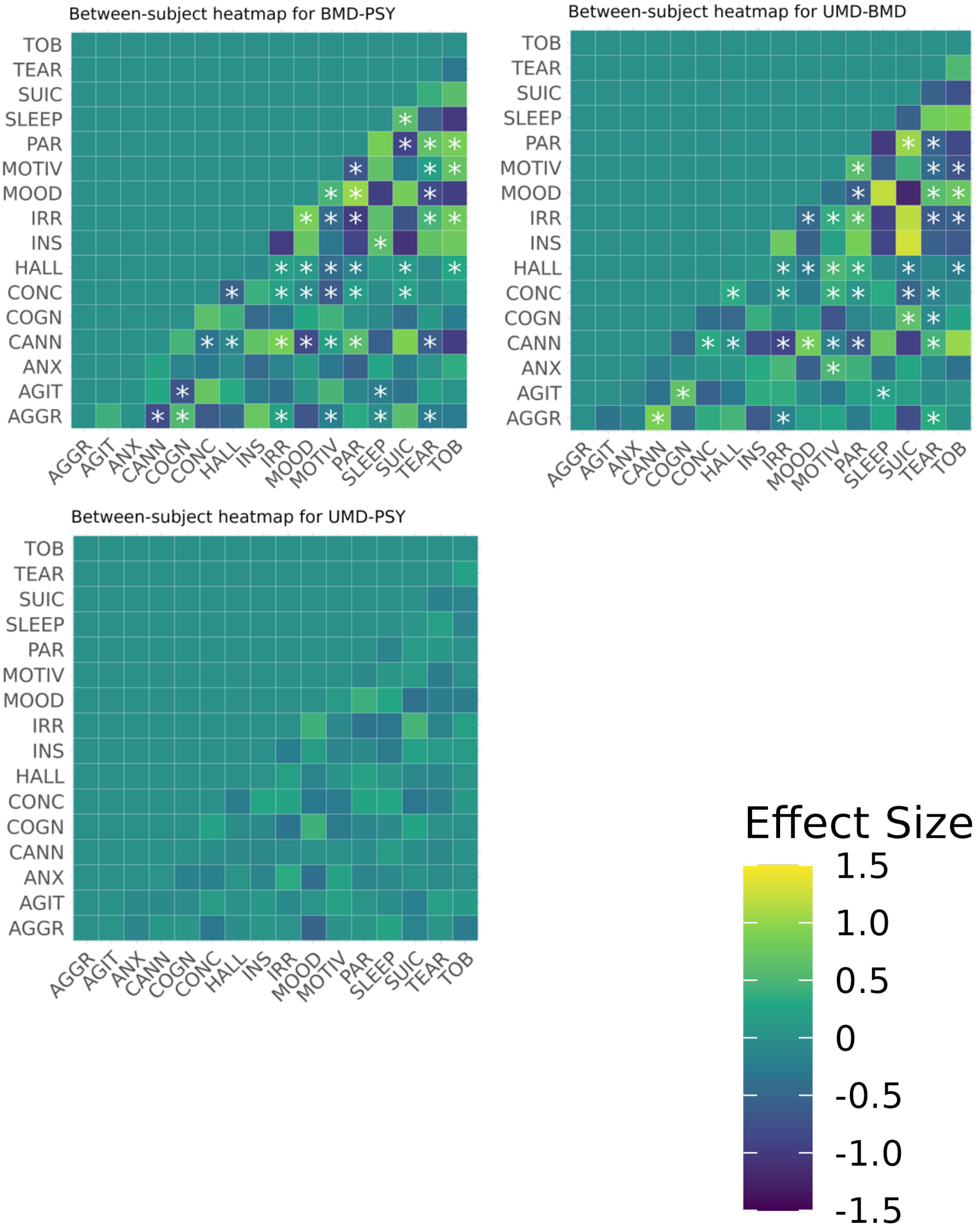

eLimitations 1

While the use of EHRs in this study has high ecological validity, the symptoms recorded in clinical notes are not psychometrically validated. However, the use of structured diagnostic interviews can itself lead to selection biases [28], and there is meta-analytical evidence indicating administrative data recorded in EHR are generally predictive of true validated diagnoses.[29] Furthermore, NLP tools generate some degree of noise as it is impossible to extract data from free text with 100% precision; clinician subjectivity, including structural or unconscious bias, can impact how symptoms are recorded for given individuals, thereby reducing standardisation of output [30]. We mitigated against this issue by pre-selecting NLP algorithms for an adequate level of precision (≥80%). Moreover, we could not externally validate these findings, and therefore their generalisability to other healthcare settings should be confirmed. Finally, the date of diagnosis may not be accurate regarding the timing of disorder onset but we mitigate against representing full-threshold symptoms by excluding the six months prior to diagnosis from our analyses.
